# Prioritizing Genes and Rare Protein-Coding Variants in Acute Myeloid Leukemia via Whole Genome Sequencing Data

**DOI:** 10.64898/2026.08.19.26360760

**Authors:** Stephen Vieno, Madhurbain Singh, Sydney Kramer, Chris Chatzinakos, Roseann Peterson, Brien Riley, Silviu-Alin Bacanu, Thang Dinh, Bon Q. Trinh, Tan-Hoang Nguyen

## Abstract

The extent to which rare and common genetic variants jointly contribute to the risk of acute myeloid leukemia (AML) still remains relatively unexplored in large-scale biobank whole-genome sequencing cohorts. Here, we leverage the latest sequencing and phenotypic data from the All of Us Research Program to identify variants, genes, and gene-sets associated with AML. We performed set-based association tests for rare protein-coding variants (N_cases_=265 and N_controls_=169,706) and single-variant association tests for common variants (N_cases_=265 and N_controls_=169,705) utilizing the large European-like ancestry sample. For the rare-variant set-based tests conducted using SAIGE-GENE+, four genes were statistically significant: *DNMT3A*, *TET2*, *SRSF2*, and *IDH2* (Bonferroni-corrected Cauchy p-value < 0.05). We also constructed multiple rare-variant burden risk scores using different gene-sets to identify those with a substantial rare-variant burden for AML. Gene-sets derived from Genomic Data Commons whole-genome sequencing data, comprising two distinct groups—genes observed to harbor somatic mutations in AML and genes observed to harbor somatic mutations across all cancer types—showed a statistically significant rare-variant burden (Bonferroni-corrected p-value < 0.05). Ultimately, these findings demonstrate that leveraging whole-genome sequencing in large-scale biobanks enables the identification of rare protein-coding variants, genes, and gene sets associated with AML.

**Graphical abstract:** 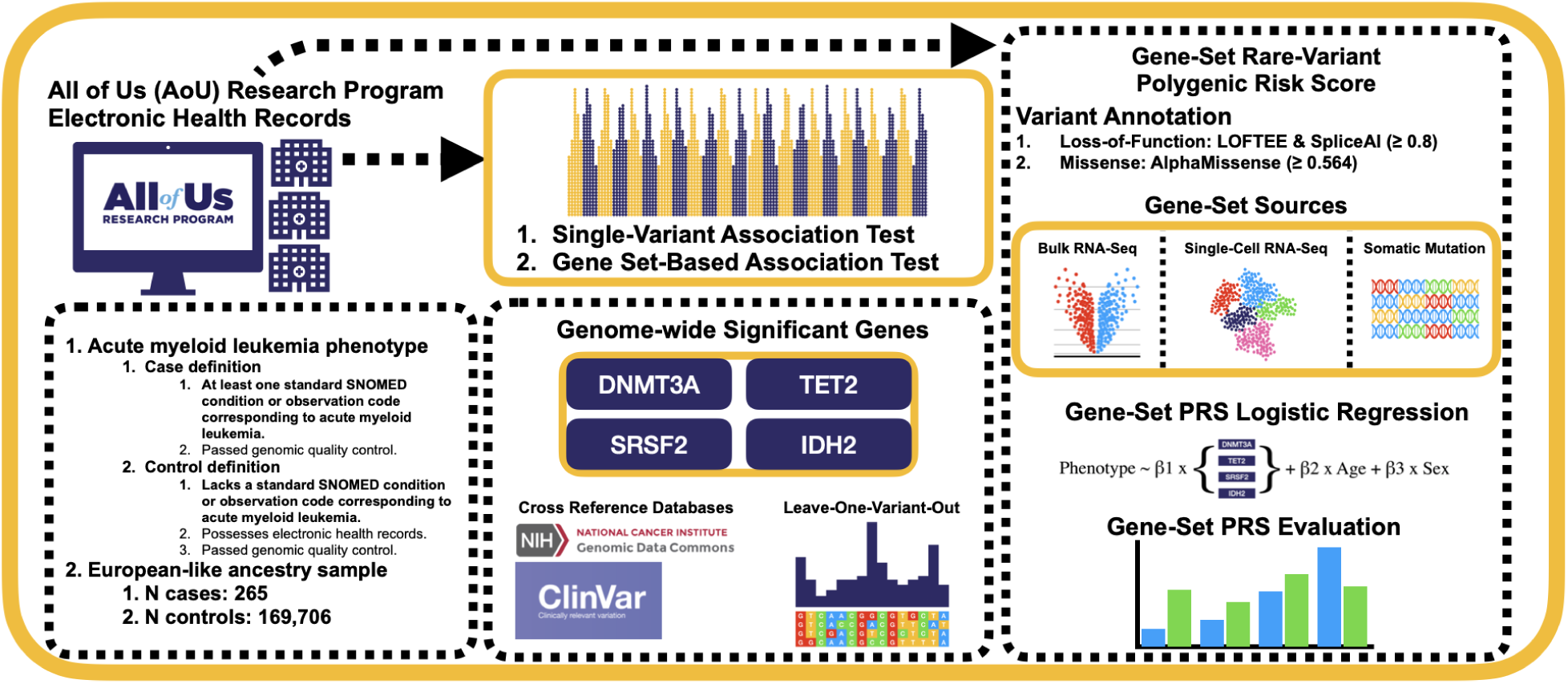

## Background

Acute myeloid leukemia (AML), a hematologic malignancy, continues to represent a major cause of cancer-related mortality in both pediatric and adult populations, despite advances in therapeutic strategies. Although overall survival in AML has improved in recent years with the introduction of several therapies, one-, three-, and five-year survival rates for patients diagnosed between 2011 and 2019 were relatively low at 48%, 31%, and 28%, respectively (1). Additionally, AML is a relatively common cancer, with prevalence estimates ranging from 0.6 to 11.0 per 100,000 individuals across all age categories, genders, and ethnicities, making it a considerable public health issue (2). Given its persistently poor outcomes and prevalence, further research into AML is warranted to inform therapeutic development. This begins with identifying genomic alterations that implicate genes and biological pathways as therapeutic targets.

To date, numerous methodologies have been employed to characterize the underlying heterogeneity of both somatic and germline alterations. For example, somatic profiling has identified a series of genes harboring variants and structural alterations associated with AML, including tumor protein p53 (*TP53*), isocitrate dehydrogenase 2 (*IDH2*), deoxyribonucleic acid methyltransferase 3 alpha (*DNMT3A*), and tet methylcytosine dioxygenase 2 (*TET2*) (3,4). Other studies have demonstrated significant associations with common germline variants, including a risk locus at 19q13 near BRD4 interacting chromatin remodeling complex associated protein (*BICRA*) and another at 11q13.2 near lysine methyltransferase 5B (*KMT5B*) (5,6). Additionally, rare pathogenic or likely pathogenic germline variants have also been reported in AML patients in genes associated with the deoxyribonucleic acid (DNA) damage response. A substantial proportion of adult AML patients (13.6%) harbor such variants, and 6.39% carry at least one clinically actionable pathogenic germline variant (7). These studies, however, only evaluated single variants independently and therefore did not account for the aggregate contribution of multiple variants, limiting their ability to detect variants with moderate effect sizes. Furthermore, a comprehensive study of both rare and common variants using population biobank data has not yet been conducted for AML.

Large-scale sequencing data from biobanks provide an opportunity to elucidate the contributions of individual germline variants and their aggregate effects on AML genetic risk. Hence, we conducted analyses in the large United States-based biobank, the All of Us Research Program (AoU). The AoU contains whole-genome sequencing data from over 400,000 participants, as well as survey and electronic health record (EHR) data that serve as resources for phenotyping (8). By leveraging the EHRs and large-scale sequencing data within the AoU, we performed genome-wide association analyses focusing on rare protein-coding variation. Our goal was to identify variants, genes, and biological pathways involved in leukemogenesis. Through these analyses, we identified genes and gene-sets in which cumulative rare protein-coding variation is associated with AML, providing additional insights into genetic predisposition to AML.

## Methods

The primary analysis steps are outlined below. To ensure full reproducibility within manuscript length constraints, the complete genomic quality control workflow; including variant filtering, sample-level quality control, and population structure controls; is detailed extensively in the Supplementary Methods.

### Phenotypes

EHRs were used to classify participants as either cases or controls. Within the All of Us (AoU) Research Program, multiple EHR *Source Vocabulary Systems* (e.g., ICD-9-CM and ICD-10-CM) are mapped to the *Standard Vocabulary System*, Systematized Nomenclature of Medicine (SNOMED) for condition codes, providing a unified terminology for defining cases and controls. Participants were classified as cases if they had at least one EHR from either *Condition* codes or *Observation* codes from the *Standard Vocabulary System* present within the AoU (8). The codes used to define cases were *Acute myeloid leukemia, disease* (Concept Identifier: 140352, Vocabulary System: SNOMED, Code: 91861009) and *Acute myeloid leukemia, minimal differentiation* (Concept Identifier: 4304199, Vocabulary System: SNOMED, Code: 103689001). Controls were participants who were not cases and had EHRs available within the AoU. Neither cases or controls were excluded on the basis of the presence of another malignancy in the EHR. Only excluding participants from either cases or controls based on non-AML-related EHRs introduces differences between cases and controls that are not related to our outcome of interest, thereby making our results less specific to AML and altering the heritability of the underlying trait (9).

### Rare-variant analysis

We analyzed variant and gene-level associations using SAIGE-GENE+, a linear mixed model able to handle case-control imbalance (10). Using these results, we then performed gene-set enrichment analysis. We also conducted burden analyses to further understand the contribution of rare variants.

### Covariate information

The covariates for analyses employing SAIGE-GENE+ included age, sex, and the first 10 within-ancestry principal components (10). Additionally, we conducted a secondary analysis that included the deprivation index as a covariate to evaluate for potential confounding (11). Age was defined as the duration in years from birth to the last documented EHR. To account for the difficulty in determining the age of diagnosis within the AoU cohort, we performed a secondary analysis focused on age at specimen collection. This enabled a more robust comparison of our results under two age definitions. All quantitative covariates were standardized.

### Genetic relationship matrix

After excluding ancestry group outliers and participants who failed the whole-genome sequencing (WGS) quality control procedures (Supplementary Methods), genotype microarray data were filtered and pruned within the European-like (EUR-like) ancestry participants. Variants were first filtered based on the following criteria: minor allele frequency (MAF) > 1%, missingness < 1%, Hardy-Weinberg equilibrium p-value > 10^-6^, removal of duplicated variants, strand-ambiguous variants, and multi-allelic variants (12). Then a single round of linkage disequilibrium pruning was performed using PLINK2 with the parameters --indep-pairwise 50 5 0.05, as recommended by SAIGE-GENE+ (10,13,14). A sparse genetic relationship matrix (GRM) was generated using 5,000 randomly selected markers, with a relatedness cutoff of 0.05 applied according to the SAIGE-GENE+ protocol (10,14). The resulting sparse GRM was used in both the rare set-based tests and common single-variant tests for SAIGE-GENE+.

### Gene set-based tests

Set-based tests, including Burden, sequence kernel association test (SKAT), and optimal sequence kernel association test (SKAT-O), were conducted using SAIGE-GENE+ version 1.5.0.2 on quality-controlled rare variants from the Exome WGS dataset in protein-coding genes (Supplementary Methods). For all set-based tests, the variance ratio file was generated by selecting 1,000 variants from each minor allele count (MAC) bin: 10 ≤ MAC < 20 and MAC ≥ 20. The sparse GRM was then used to fit the null model without genetic contribution for each phenotype, employing the options --useSparseGRMtoFitNULL=TRUE and --isCateVarianceRatio=TRUE (10,14).

Multiple variant masks were used, with maximum MAF cutoffs of 1%, 0.1%, and 0.01%, and annotation masks of loss-of-function (LoF), damaging missense, and LoF + damaging missense. LoF variants were defined as high-confidence variants from loss-of-function transcript effect estimator (LOFTEE) or as variants with a SpliceAI delta score greater than or equal to 0.8 (15,16). Missense variants were defined as those with an AlphaMissense predicted probability greater than 0.564 (17). Only consequences on the Matched Annotation from the NCBI and EMBL-EBI (MANE) Select protein-coding transcripts were used for assigning variants to annotation masks (18).

In a secondary analysis, we applied the same MAF thresholds while restricting the annotation mask to synonymous variants. Synonymous variants were defined as variants whose most severe predicted protein-coding consequence on the MANE Select transcript, as annotated by Variant Effect Predictor (VEP) version 115, was classified as ‘synonymous variant’ and had a SpliceAI score below 0.5 (16,18,19).

To combine p-values across masks, SAIGE-GENE+ used the Cauchy combination method (20). Exome-wide significance was defined as a Bonferroni-corrected Cauchy p-value < 0.05 (21).

### Independent validation of gene set-based results using Genebass

To demonstrate the reproducibility of our gene set-based results, we cross-referenced our findings with results from Genebass (22). Genebass is a web-based browser for exploring rare-variant association results and provides rare-variant association analyses for 4,529 phenotypes using single-variant and gene-based tests in 394,841 individuals from the UK Biobank. We searched Genebass for the closest matching phenotype, Myeloid leukemia (ICD 10 C92). Myeloid leukemia which comprises both Acute myeloid leukemia (CD92.A) and Chronic myeloid leukemia (C92.1), was used as the closest available proxy phenotype for acute myeloid leukemia. Exome-wide significance was defined as a Bonferroni-corrected SKAT-O p-value < 0.05, accounting for 37,761 gene-based tests (significance threshold = 1.32 x 10^-6^) (21).

### Leave-one-variant-out analysis

To determine the relative contribution of each rare variant to the observed gene-phenotype association, a leave-one-variant-out analysis was performed. Set-based tests were conducted iteratively while excluding one variant at a time from the variant set. If the exclusion of a single variant caused the Bonferroni-corrected Cauchy p-value to no longer be significant (adjusted p-value > 0.05), that variant was considered to predominantly account for the gene-phenotype association (20,21,23). In the main results, we reported the results from the gene set-based tests without conditioning on the deprivation index, as this provided the largest variant set; however, the results for the gene set-based tests conditioning on the deprivation index are reported in the Supplementary Materials.

### Gene-set association analysis using sparse signals

Gene-set Association Analysis using Sparse Signals (GAUSS) was used to conduct gene-set (pathway) association analyses (24). This method produces a p-value for each gene-set based on gene-level tests, including Burden, SKAT, and SKAT-O. Using the Cauchy-combined SKAT-O p-values from SAIGE-GENE+ for AML, gene-set analyses were performed with GAUSS. Analyses were conducted across both the Gene Ontology (GO) and Kyoto Encyclopedia of Genes and Genomes (KEGG) pathway collections. Additionally, gene-sets from the Molecular Signatures Database (MSigDB) were evaluated; however, due to overlap among gene-sets in MSigDB, they were filtered to include only AML-related gene-sets. In total, 64 AML-specific gene-sets were tested.

### Gene-set protein-coding variant burden

To better understand the impact of rare protein-coding variants in aggregate, we calculated burden scores by summing the rare predicted deleterious variants (MAF < 1%) identified in each gene-set. This effectively constructed a rare-variant protein-coding burden score for each gene-set, serving as a synthetic genetic marker for use in association testing. Predicted deleterious variants were defined using the same criteria as in the gene set-based analysis (either LoF or damaging missense) (15–18). Logistic regression models (‘stats’ R package) were fit separately for each gene-set, with the gene-set burden score as a predictor of the AML phenotype (25). Other covariates for the logistic regression models included the total count of rare variants across the exome (including both protein-coding and non-coding), sex, age, and the first ten ancestry-specific principal components (26). All quantitative predictors were standardized. Details on how the predefined gene-sets were derived are provided in the Supplementary Methods.

### Sensitivity analysis

To better understand our results, we conducted several additional analyses. These include adjustments for the deprivation index, refined age definitions, and biospecimen collection matching. Detailed descriptions of these analyses are provided in the Supplementary Materials.

### Single common variant analysis

To determine which class of genomic variation warrants further investigation, common or rare, a common-variant genome-wide association study (GWAS) (MAF > 1%) was performed alongside rare-variant analyses. Genome-wide single common variant association testing was conducted using SAIGE-GENE+ version 1.5.0.2 on the quality-controlled allele count / allele frequency (ACAF)-threshold WGS dataset (Supplementary Methods) (10). To generate the variance ratio file, 1,000 variants were randomly selected. The sparse GRM was then used to fit the null model without genetic contribution, employing the options --useSparseGRMtoFitNULL=TRUE and --skipVarianceRatioEstimation=FALSE. Single-variant association tests were then conducted using Firth logistic regression, employing the options --is_Firth_beta=TRUE and--pCutoffforFirth=0.1 (27). Firth logistic regression is used to address quasi- or complete separation, which can lead to inflated coefficient estimates and standard errors (28). It is commonly applied in genetic association studies to account for rare predictors (low-frequency variants) or low prevalence outcomes (case-control imbalance) (29). In this analysis, we employed Firth logistic regression to correct for the low prevalence of cases relative to controls and applied it only when the p-value fell below a pre-specified threshold. Genome-wide significance was defined as p-value < 5 × 10^-8^ (30).

### Gene based and pathway analysis for common variant results

Multi-marker Analysis of GenoMic Annotation (MAGMA) version 1.10 was used to map variant associations to genes and gene-sets (31). Before deploying MAGMA, variants were first mapped to rsIDs using GWASLab and Single Nucleotide Polymorphism Database (dbSNP) Build 157 (32,33). Subsequently, variants were then mapped to genes employing 35 kb upstream and 10 kb downstream windows. MAGMA gene tests were followed by gene-set tests using gene-sets from MSigDB (release 2025.1.Hs) (34). For all MAGMA analyses, European samples from Phase 3 of the 1000 Genomes Project were used as the reference panel (35).

Additional tests were performed using the same methodology described above, except that the window size was varied to assess its effect on MAGMA results. Window-size effects were evaluated across 11 iterations, beginning with 0 kb upstream and 0 kb downstream windows. At each iteration, the upstream window size was increased by 10 kb and the downstream window size by 5 kb, reaching a final window size of 100 kb upstream and 50 kb downstream. Gene and gene-set p-values were then combined using the Cauchy combination test, and significance was assessed after applying a correction for multiple hypothesis testing (20,21).

### Statistical analysis

We applied a Bonferroni correction to account for multiple testing across all analyses (21). For all tests, unless otherwise specified, an adjusted p-value < 0.05 was used to determine statistical significance. We followed the standard analytical protocols provided by the software packages used in this study.

## Results

### Phenotype distribution across ancestry assignments

Ancestry assignment was conducted using the *POP-MaD* (population grouping by Mahalanobis distance) pipeline (36,37). Using this pipeline, 82,675 samples were assigned to the African-like (AFR-like), 94,245 to the Admixed American-like (AMR-like), 7,972 to the Central/South Asian-like (CSA-like), 10,009 to the East Asian-like (EAS-like), 239,578 to the European-like (EUR-like), 2,687 to the Middle Eastern-like (MID-like), and 60 to the Oceanian-like (OCE-like) ancestry groups.

Following ancestry assignment, we identified 774 participants with at least one EHR record corresponding to an AML diagnosis in the AoU, who were considered cases. After intersecting these cases with the ancestry-assigned cohort, the number of cases decreased to 479. Of these, 286 were EUR-like, 106 were AMR-like, 65 were AFR-like, and fewer than 20 participants were classified as EAS-like, MID-like, CSA-like, or Unassigned. Given the substantial reduction in case counts among non-EUR-like ancestries, subsequent quality control analyses were restricted to participants assigned to the EUR-like ancestry group.

After sequencing data quality control and restricting analyses to participants with available EHR records, the primary analysis cohort consisted of 265 cases (N_females_=145; N_males_=120; median age [interquartile range (IQR)]: 68.30 [57.30-75.38]) and 169,706 controls (N_females_=102,694; N_males_=67,012; median age [IQR]: 62.29 [46.29-72.30]). For analyses incorporating the deprivation index, which was not available for all participants, the sample size was further reduced to 253 cases (N_females_=137; N_males_=116; median age [IQR]: 69.30 [58.29-76.00]) and 165,704 controls (N_females_=100,247; N_males_=65,457; median age [IQR]: 62.29 [46.29-72.30]). The sample size was insufficient to analyze AML subgroups; therefore, we analyzed AML as a single phenotype in all analyses.

### Discovery of genome-wide significant protein-coding genes via gene-level tests

To investigate the potential impact of rare protein-coding variants on an individual’s predisposition to AML, a rare-variant set-based gene analysis using SAIGE-GENE+ was conducted within the AoU for 18,064 protein-coding genes (N_cases_=265 and N_controls_=169,706) (Table S1). Examining the SKAT-O p-value results shows that, overall, the combination of LoF and missense variants produced the strongest genomic signal for most genes. This signal was evident when the maximum MAF thresholds were set at 1% and 0.1%, with only a few genes reaching genome-wide significance at a maximum MAF threshold of 0.001%. These findings indicate that, for the majority of genes, the association signal is primarily driven by variants with minor allele frequencies between 0.01% and 1% (Figure S1, Tables S2-S5).

In total, four genes—*DNMT3A* (adjusted p-value = 4.44 x 10^-10^), *TET2* (adjusted p-value = 2.92 x 10^-8^), serine and arginine rich splicing factor 2 (*SRSF2*) (adjusted p-value = 1.53 x 10^-7^), and *IDH2* (adjusted p-value = 1.34 x 10^-6^)—were identified as genome-wide significant for the LoF and damaging missense variant set (Table 1). As these identified genes are known to harbor somatic mutations that accumulate with age at biospecimen collection, we performed additional set-based tests matching cases and controls using age at biospecimen collection across multiple case-control ratios (Supplementary Methods). The results of these analyses suggest that, while the impact of age-related somatic burden cannot be fully excluded, the observed associations between rare deleterious variation and AML are primarily attributable to germline variation rather than somatic mutational burden.

Adjusting for the deprivation index generally increased the significance of the p-values, a trend observed across the majority of genes, including the most significant ones (Figure 1, Table 1). Of the significant genes, *TET2*, showed the greatest improvement in the p-value after including the deprivation index (adjusted p-value = 3.35 x 10^-9^).

For both SKAT-O and Cauchy combination p-values, none of the gene-based tests restricted to synonymous variants reached genome-wide significance, with the lowest adjusted p-value observed for TBC1 domain family member 21 (*TBC1D21*) (adjusted p-value = 3.10 x 10^-5^) (Figure S2, Tables S6-S9).

**Figure 1.**
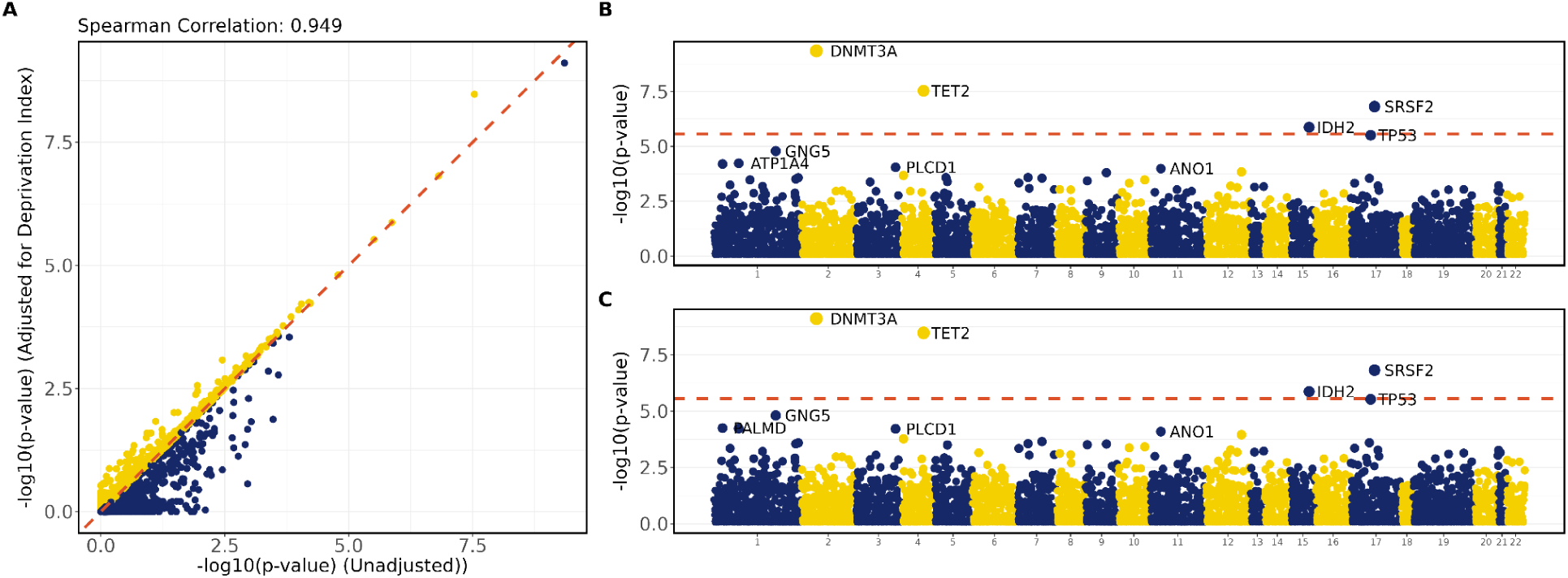
Cauchy p-values for AML from rare-variant set-based tests. (A) **Correlation of** −log10 p-values for Cauchy SKAT-O results, comparing models unadjusted and adjusted for the deprivation index. The Spearman correlation coefficient is shown in the top-left corner. **(B)** Manhattan plot of rare-variant set-based gene association results from SAIGE-GENE+, unadjusted for the deprivation index. The red dashed line indicates the genome-wide significance threshold after Bonferroni multiple testing correction. **(C)** Manhattan plot of rare-variant set-based gene association results from SAIGE-GENE+, adjusted for the deprivation index. The red dashed line indicates the genome-wide significance threshold after Bonferroni multiple testing correction.

**Table 1.** Significant rare-variant set-based associations for AML. Significant Cauchy SKAT-O p-values (Bonferroni-adjusted p-values < 0.05) from SAIGE-GENE+ rare-variant analysis of AML. Analysis was performed with and without the deprivation index as a covariate to assess environmental influence on genetic associations.

| Gene | Cauchy p-value | Cauchy p-value (deprivation index) | Bonferroni p-value | Bonferroni p-value (deprivation index) |
| --- | --- | --- | --- | --- |
| DNMT3A | $4.44 \times 10^{-10}$ | $7.78 \times 10^{-10}$ | $8.03 \times 10^{-6}$ | $1.41 \times 10^{-5}$ |
| TET2 | $2.92 \times 10^{-8}$ | $3.35 \times 10^{-9}$ | $5.27 \times 10^{-4}$ | $6.04 \times 10^{-5}$ |
| SRSF2 | $1.53 \times 10^{-7}$ | $1.51 \times 10^{-7}$ | $2.77 \times 10^{-3}$ | $2.73 \times 10^{-3}$ |
| IDH2 | $1.34 \times 10^{-6}$ | $1.34 \times 10^{-6}$ | $2.43 \times 10^{-2}$ | $2.42 \times 10^{-2}$ |

### Cross-referencing results with the UK Biobank demonstrate reproducibility of results

To determine if our results were reproducible with another out-of-sample genome sequencing biobank, we cross-referenced our results with results from the UK Biobank. We searched Genebass for the closest matching phenotype, Myeloid leukemia (ICD 10 C92). Myeloid leukemia (ICD 10 C92; N_cases_=424, N_controls_=394,417), which comprises both Acute myeloid leukemia (CD92.A) and Chronic myeloid leukemia (C92.1), was used as the closest available proxy phenotype for acute myeloid leukemia. Genebass results demonstrated significant associations for LoF variants in *TET2* (SKAT-O p-value = 1.53 x 10^-25^) and missense or LOFTEE low-confidence variants in *DNMT3A* (SKAT-O p-value = 2.58 x 10^-7^). Associations for missense or LOFTEE low-confidence variants in *IDH2* (SKAT-O p-value = 4.08 x 10^-6^) and *SRSF2* (SKAT-O p-value = 3.19 x 10^-5^) approached exome-wide significance.

### Quantifying the contribution of individual rare variants via leave-one-variant-out analysis

A leave-one-variant-out analysis was performed to identify which variants contributed the most to the significance of identified genes—*DNMT3A*, *TET2*, *SRSF2*, and *IDH2*. For *DNMT3A*, removing chr2:25234373:C:T, a variant previously identified in the Genomic Data Commons (GDC) and ClinVar, almost resulted in a reduction of the p-value to below genome-wide significance (AlphaMissense-0.564, Cauchy p-value = 2.3 x 10^-6^). *TET2* demonstrated that removing any single variant did not result in a reduction of the p-value to below genome-wide significance, thereby demonstrating that a group of variants are contributing to the significance observed for *TET2* (Figures S3-S4, Tables S10-S11).

On the other hand, for *SRSF2* (chr17:76736877:G:T, AlphaMissense-0.564, Cauchy p-value = 3.3 x 10^-3^; chr17:76736877:G:A, AlphaMissense-0.564, Cauchy p-value = 4.7 x 10^-6^) and *IDH2* (chr15:90088702:C:T, AlphaMissense-0.564, Cauchy p-value = 5.2 x 10^-1^), removing a single variant led to a reduction in the p-value below genome-wide significance. Each of these variants that resulted in the loss of genome-wide significance is already recorded in both the GDC and ClinVar (Figures S3-S4, Tables S10-S11).

### Gene-set association analysis using sparse signals to identify myeloid expression profile

Using the gene-level p-values from the set-based tests, gene-set enrichment analyses were performed with GAUSS (Tables S12). Across both the GO and KEGG gene-set collections, no gene-sets were significant (Figure S5, Tables S12). Given the absence of significant associations in these biological pathway gene-sets, additional AML-specific gene-sets from MSigDB were evaluated to determine whether any signals could be detected. In total, 64 additional gene-sets were tested, and only one reached significance: [VALK AML CLUSTER 9](https://www.gsea-msigdb.org/gsea/msigdb/human/geneset/VALK_AML_CLUSTER_9.html) (P = 1.24 × 10^−5^, adjusted p-value = 9.47 × 10^-3^) (Figure S5, Table S12). This gene-set consists of the top 40 genes from Cluster 9 of the AML expression profile. The expression profile for this cluster is derived from samples corresponding to the French-American-British classification M4 or M5 subtypes, all of which harbor the inv(16) chromosomal inversion that produces the core-binding factor subunit beta-myosin heavy chain 11 fusion (*CBFB*-*MYH11*) fusion, a genetic event typically associated with a favorable survival prognosis (34).

### Identifying significant gene-sets via global rare protein-coding variant burden analysis

In addition to the gene-set enrichment analysis using p-values from SAIGE-GENE+, we evaluated gene-set rare-variant burdens by performing association tests directly on exome-wide variant counts. Several AML-related gene-sets were curated from multiple sources, including both transcriptomic and genomic data (Tables S13-S16). We observed that gene-sets composed of genes frequently observed to harbor somatic mutations—first in AML specifically and subsequently across all cancer types within the GDC—performed best overall (Table 2, Tables S17-18). When the AML-specific somatic mutation gene-set was further subsetted by the deleteriousness of observed somatic mutations in the GDC, those containing more deleterious mutations tended to show stronger associations; however, this pattern was not consistent for gene-sets derived from observed somatic mutations across all cancer types in the GDC (Table 2, Tables S17-18). Among the transcriptomic-derived gene-sets, only the single-cell ribonucleic acid sequencing-derived set comparing dendritic cells to other cell types, including both malignant and non-malignant cells, remained statistically significant after correction for multiple testing (Table 2, Tables S17-18). Together, these findings suggest that genes enriched for rare deleterious variants also tend to be genes that harbor somatic mutations in AML.

**Table 2.** Significant gene-sets from rare-variant gene-set burden score analysis. These results are for the significant gene-sets after Bonferroni correction for multiple hypothesis testing (Bonferroni-adjusted p-values < 0.05). Gene-set p-values were obtained from logistic regression models regressing the gene-set rare-variant burden score, exome rare-variant burden, age, sex, and the first 10 principal components on the AML phenotype in both related and unrelated individuals. (Results restricted to unrelated individuals, excluding related individuals, are provided in the Supplementary Materials.)

| Gene-Set Description | Odds Ratio | 95% Confidence Interval | p-value | Bonferroni p-value |
| --- | --- | --- | --- | --- |
| AML frequently mutated census genes protein coding VEP moderate high logistic summary | 1.34 | (1.21,1.48) | $2.79 \times 10^{-8}$ | $4.16 \times 10^{-6}$ |
| AML frequently mutated census genes protein coding VEP high logistic summary | 1.29 | (1.17,1.42) | $1.14 \times 10^{-7}$ | $1.69 \times 10^{-5}$ |
| AML frequently mutated census genes protein coding logistic summary | 1.28 | (1.15,1.43) | $9.40 \times 10^{-6}$ | $1.40 \times 10^{-3}$ |
| cancer frequently mutated census genes protein coding logistic summary | 1.28 | (1.15,1.42) | $1.09 \times 10^{-5}$ | $1.63 \times 10^{-3}$ |
| cancer frequently mutated census genes protein coding VEP moderate high logistic summary | 1.28 | (1.15,1.42) | $1.09 \times 10^{-5}$ | $1.63 \times 10^{-3}$ |
| cancer frequently mutated census genes protein coding VEP high logistic summary | 1.28 | (1.14,1.42) | $1.14 \times 10^{-5}$ | $1.69 \times 10^{-3}$ |
| conventional dendritic cell vs other cells single cell downregulation significant results logistic summary | 1.21 | (1.1,1.34) | $1.85 \times 10^{-4}$ | $2.75 \times 10^{-2}$ |

### Lower signal in single common variants versus rare protein-coding variants

To examine the contribution of common variants to the genetic architecture of AML, a genome-wide association analysis was conducted using SAIGE-GENE+ (N_cases_=265 and N_controls_=169,705). No variants reached genome-wide significance (Figures S6-S7, Tables S19-S20). To identify genes and biological pathways potentially implicated in AML predisposition, we next performed gene and gene-set analyses using MAGMA. We first mapped variants to genes and then genes to gene-sets, applying different window thresholds to define the range of possible variant-to-gene mappings (Figures S8-S11). These results were meta-analyzed to identify significant genes using the Cauchy combination; however, similar to the variant-level results, no genes or biological pathways showed evidence of statistical association after multiple testing correction. The top gene and gene-set were zinc finger protein 282 (*ZNF282*), Cauchy p-value = 8.17 x 10^-6^, adjusted p-value = 0.16) (Tables S21-S22) and gene-sets (Top Gene-Set: Hesson Tumor Suppressor Cluster 3p21.3, Cauchy p-value = 2.53 x 10^-6^, adjusted p-value = 0.09) (Tables S23-S24).

## Discussion

Our analysis reveals that rare protein-coding variants from biobank data converge with somatic mutations at the variant, gene, and biological pathway levels. At the variant level, all identified genome-wide significant genes harbor rare protein-coding variants that resemble somatic mutations in the GDC. Through our leave-one-variant-out analysis, we were able to demonstrate that a proportion of these same variants contribute to the observed gene-phenotype associations.

In terms of genes, all identified genome-wide significant genes—*DNMT3A*, *TET2*, *SRSF2*, and *IDH2*—are well-established, clinically relevant genes for AML. These genes are among the most frequently mutated in AML, with *DNMT3A* and *TET2* being the most frequently mutated genes (38–41). Additionally, somatic mutations in these genes have been associated with poorer survival outcomes, highlighting their role as key determinants of disease progression (38,40,42,43).

Intriguingly, the top enriched rare variants we identified in AML are in key genes that regulate several critical molecular pathways related to DNA methylation. DNA methyltransferase 3A enzyme (encoded by *DNMT3A*) sets up new methylation marks, whereas the epigenetic enzyme ten-eleven translocation 2 (encoded by *TET2*) promotes DNA demethylation (41,42).

On the other hand, the splicing factor serine and arginine rich splicing factor 2 (encoded by *SRSF2*) promotes exon inclusion or affects alternative splicing decisions (44). Isocitrate dehydrogenase 2 (encoded by *IDH2*) is a key enzyme in the Krebs cycle, catalyzing the conversion of isocitrate to α-ketoglutarate (45). Of note, *SRSF2* and *IDH2* indirectly impact DNA methylation by interacting with enzymatic regulation of the methylome (e.g., TET enzymes, DNA methyltransferases, and associated epigenetic modifiers). Mutation of *SRSF2* leads to mis-splice genes encoding transcription factors and epigenetic regulators (including those affecting DNA methylation) (46). Pathogenic somatic mutations in *IDH2* lead to increased production of the oncometabolite 2-hydroxyglutarate, which in turn interferes with enzymes regulating DNA and histone demethylation, such as *TET2* (45). Further supporting the relevance of this shared biological pathway in AML, hypomethylating agents constitute a cornerstone of therapy. These agents, including 5-azacytidine (AZA) and 5-aza-2′-deoxycytidine (decitabine, DAC), inhibit DNA methyltransferases and are widely used in treatment (41). In addition, *IDH2* is therapeutically targetable with enasidenib, which is approved for AML with *IDH2* mutations (45,47). Our findings, therefore, provide a foundation for further investigation into the functional consequences of these rare variants in AML pathogenesis.

While our approach successfully identified rare variation associations in AML, several limitations should be considered. First, EHR data within the AoU do not represent participants’ complete medical histories due to the manner of data collection. Second, sample size was insufficient to analyze within-phenotype subtypes. Third, AoU quality-controlled variants may comprise a mixture of germline variants and somatic mutations arising from clonal hematopoiesis or chimerism; in particular, all the genes identified are genes known to harbor clonal hematopoiesis of indeterminate potential (CHIP) variants (8,48–50). The first two limitations could not be addressed in this study, as they are intrinsic limitations of EHRs in biobanks. While we attempted to address the third limitation through a careful age-at-biospecimen-matched analysis (Supplementary Materials), we acknowledge this does not fully correct for the underlying issue. It remains possible that AML cases contain a higher burden of CHIP variants compared with controls, given the increased prevalence of CHIP among individuals with malignancies (51,52). Furthermore, without longitudinal samples collected at multiple time points or samples from multiple tissues within the same individual, it is difficult to determine whether these variants are causal. Specifically, we cannot establish whether the acquisition of CHIP preceded and contributed to AML pathogenesis or whether AML development resulted in the emergence or expansion of CHIP-associated variants.

Another approach to address the influence of clonal hematopoiesis would have been to filter out putative CHIP variants before analysis. However, filtering methods are primarily designed to identify CHIP variants with higher variant allele frequencies (typically >2–5%) and rare CHIP variants lack sufficient information to correctly identify rare CHIP variants using these methods (53,54). Furthermore, while these methods have been shown to identify likely CHIP variants, they have not been demonstrated to identify all CHIP-derived variants such that only true germline variants remain after filtering. Consequently, there is currently no effective method to identify and remove rare CHIP variants while preserving genuine rare germline variants. This represents one of challenges for rare variant association studies of hematological phenotypes in large-scale biobanks, and will require new approaches to improve upon existing methods.

There are also potential improvements to be made in the modelling methods. In terms of modelling approach, we modelled genomic-phenotype associations within each gene, ignoring the broader genomic background. However, co-mutations in different genes that share biological pathways can have a synergistic effect (44,55,56). *SRSF2* has demonstrated a synergistic effect with enhancer of zeste 2 polycomb repressive complex 2 subunit (*EZH2*) and additional sex combs like 1, transcriptional regulator (*ASXL1*), with co-mutations in *SRSF2* and *ASXL1* associated with a significantly worse survival prognosis compared to either somatic mutation alone in AML (44). Second, although the age at diagnosis is an important factor, this information could not be accurately retrieved from the biobank data. To address this, we evaluated two age definitions—age at last EHR entry and age at specimen collection—and observed consistent significant associations for the same genes across both approaches, suggesting minimal impact of age definition on the results (Supplemental Materials).

Overall, it is of considerable interest to determine whether these relationships are reflected in the accumulation of deleterious variation underlying AML predisposition. Our findings provide preliminary support, as global protein-coding burden analyses indicate that the cumulative effect of rare protein-coding variants across multiple genes contributes to AML risk. However, further studies are required to confirm this and to assess whether incorporating the broader genomic background into the genome-phenotype association analyses improves characterization of the genetic architecture of AML.

## Conclusion

The findings from this work, together with the expanding literature on the contribution of germline variation to malignancy, demonstrate that rare variation warrants further investigation to clarify its role in the heritability of cancers such as AML. Although there are established genetic syndromes in which a single rare germline variant confers substantial risk, for most individuals, genetic susceptibility to AML is likely driven by the combined effects of multiple genomic alterations. By expanding upon this work and leveraging large-scale biobanks, it may be possible to better understand the effects of pathogenic variation on the distribution of normal cellular processes that eventually lead to leukemogenesis.

## Data Availability

The data used in this study are publicly available. Individual-level data from the AoU Curated Data Repository version 8 [https://www.researchallofus.org] were accessed following approval of the Data Use and Registration Agreement. VEP version 115, along with associated annotation plugins, was used to annotate variants present in the AoU dataset [https://github.com/Ensembl/ensembl-vep/archive/release/115.zip]. Clinical information, somatic mutation frequencies, and RNA-sequencing (RNA-seq) expression data were obtained from the Genomic Data Commons (September 18, 2025 - December 1, 2025) [https://portal.gdc.cancer.gov]. Single-cell RNA-sequencing data (GSE116256) were obtained from the Gene Expression Omnibus on December 10, 2025 [https://www.ncbi.nlm.nih.gov/geo/query/acc.cgi?acc=GSE116256]. Variants present in Clinvar were obtained on April 12, 2026 [https://www.ncbi.nlm.nih.gov/clinvar/]. The AoU does not permit the public sharing of AoU workspaces; however, to facilitate reproducibility, all code implemented within the AoU during this research is available in the accompanying GitHub repository, along with the corresponding summary statistics [https://github.com/StephenVieno/Rare-Variant-Analysis-of-EHR-Derived-AML-in-the-AoU.git]. In accordance with AoU publication policies, participant counts below 20 and allele counts below 40 are not reported in the publicly available summary statistics.

https://github.com/Ensembl/ensembl-vep/archive/release/115.zip

https://portal.gdc.cancer.gov

https://www.ncbi.nlm.nih.gov/geo/query/acc.cgi?acc=GSE116256

https://www.ncbi.nlm.nih.gov/clinvar/

https://github.com/StephenVieno/Rare-Variant-Analysis-of-EHR-Derived-AML-in-the-AoU.git

## Declarations

Not applicable

## Ethics approval and consent to participate

Not applicable

## Consent for publication

Not applicable

## Availability of data and materials

The data used in this study are publicly available. Individual-level data from the AoU Curated Data Repository version 8 [https://www.researchallofus.org] were accessed following approval of the Data Use and Registration Agreement. VEP version 115, along with associated annotation plugins, was used to annotate variants present in the AoU dataset [https://github.com/Ensembl/ensembl-vep/archive/release/115.zip]. Clinical information, somatic mutation frequencies, and RNA-sequencing (RNA-seq) expression data were obtained from the Genomic Data Commons (September 18, 2025 - December 1, 2025) [https://portal.gdc.cancer.gov]. Single-cell RNA-sequencing data (GSE116256) were obtained from the Gene Expression Omnibus on December 10, 2025 [https://www.ncbi.nlm.nih.gov/geo/query/acc.cgi?acc=GSE116256]. Variants present in Clinvar were obtained on April 12, 2026 [https://www.ncbi.nlm.nih.gov/clinvar/].

The AoU does not permit the public sharing of AoU workspaces; however, to facilitate reproducibility, all code implemented within the AoU during this research is available in the accompanying GitHub repository, along with the corresponding summary statistics [https://github.com/StephenVieno/Rare-Variant-Analysis-of-EHR-Derived-AML-in-the-AoU.git]. In accordance with AoU publication policies, participant counts below 20 and allele counts below 40 are not reported in the publicly available summary statistics.

## Competing interests

The authors declare that they have no competing interests.

## Funding

Funding for this project was generously provided by the Children’s Hospital of Richmond at VCU, Child Health Research Institute, which, in turn, is funded by the Children’s Hospital Foundation; by the National Institutes of Health [R21MH137508, K25AA030072, P50-AA022537]; and by the National Alliance for Research on Schizophrenia and Depression [Young Investigator Grant 28599]; National Cancer Institute (NCI) grants R21 CA270067 and K01 CA222707; grant 134088-IRG-19-143-33-IRG from the American Cancer Society; 4-VA grant from UVA; state funding within the UVA Comprehensive Cancer Center and UVA School of Medicine (B.Q.T.).

## Authors’ contributions

S.V. (Conceptualization, Formal analysis, Visualization, Writing), M.S. (Material contribution, Reviewing, Writing), S.K. (Reviewing, Writing), C.C. (Material contribution, Reviewing), R.P. (Material contribution, Reviewing), B.P.R. (Reviewing, Writing), S-A.B. (Reviewing, Writing), T.D. (Conceptualization, Material contribution), B.Q.T. (Conceptualization [equal], Supervision [equal], Writing), T.H.N. (Conceptualization [equal], Methodology, Supervision [equal], Writing).

## Acknowledgements

We gratefully acknowledge *All of Us* participants for their contributions, without whom this research would not have been possible. We also thank the National Institutes of Health’s *All of Us* Research Program for making available the participant data examined in this study.

## Authors’ information

Not applicable

## Footnotes

Not applicable

## Supplementary Methods

### Overview of electronic health records in AoU

Within the AoU, in order to harmonize EHRs from over 50 different health care organizations, the AoU implemented the Observational Medical Outcomes Partnership (OMOP) Common Data Model version 5 to translate the EHRs from the original EHR vocabulary systems, designated as the *Source Vocabulary System*, to a single Vocabulary System, designated as the *Standard Vocabulary System* (1). This analysis utilized the OMOP mappings through the AoU Cohort Builder on the AoU Workbench to map standard vocabulary codes to source vocabulary codes, and thereby phenotype AML within the AoU.

### Deprivation index information

The deprivation index was derived from the 3-digit ZIP code using data from the U.S. Census American Community Survey. The components of the deprivation index include the following: proportion of the population receiving assisted income benefits within the past 12 months, proportion of the population aged 25 years or older with educational attainment of at least a high school or General Educational Development (GED) equivalent, median household income in the past 12 months, proportion of the population with no health insurance coverage, proportion of the population with income below the federal poverty level within the past 12 months, and proportion of houses that are vacant (2).

### Microarray quality control

This analysis utilized 414,830 microarray samples from the AoU array dataset within the AoU Curated Data Repository release C2024Q3R3 ([Genomic Research Data Quality Report](https://support.researchallofus.org/hc/en-us/article_attachments/40491583732116)) (1). Microarrays were assessed for sex concordance, call rate, and cross-individual contamination rates by the AoU. Sex concordance and call rate were evaluated using GenCall tool v3.0.0 and Picard 2.26.0 (3,4). Samples that failed sex concordance were not released by the AoU, and thus, were excluded from analysis. Only microarrays with a call rate greater than 98% were released by the AoU. Cross-individual contamination rates were assessed using BAFRegress (5). Although this metric was not used to filter microarray samples, any sample with cross-individual contamination greater than 10% were not released by the AoU. As only samples that passed both microarray and WGS data quality control procedures were used in this analysis, this effectively excluded samples with cross-individual contamination exceeding 10% from the analysis (1).

### Whole genome sequencing quality control

This analysis also utilized 414,830 participants from the AoU WGS dataset, obtained from the AoU Curated Data Repository release C2024Q3R3. The sample quality control performed by AoU was used in this analysis ([Genomic Research Data Quality Report](https://support.researchallofus.org/hc/en-us/article_attachments/40491583732116)) (1). The AoU used the DRAGEN pipeline (version 3.7.8) for WGS quality control; the DRAGEN pipeline has been previously used in other analyses of large-scale biobanks and rare variant analyses of oncological phenotypes (6–8).

The DRAGEN pipeline was used to generate sex chromosome ploidy calls based on heterozygous chrX variant calls, which were subsequently compared to self-reported sex. Participants whose self-reported sex did not match sex chromosome ploidy calls from the DRAGEN pipeline were not released by the AoU. To check for swapped samples and large amounts of sample contamination, AoU assessed fingerprint concordance, defined as the concordance between WGS samples and their matching arrays. Samples were released by the AoU if the log-likelihood ratio of fingerprint concordance was greater than -3 (logarithm of the odds > -3). Cross-individual contamination rate, defined as the proportion of data originating from an individual other than the one being processed, was measured. Samples with a cross-individual contamination rate ≥ 3% were excluded. Additionally, WGS samples with corresponding arrays that had a contamination rate greater than 10% were also not released. The AoU intended to restrict samples to those with ≥ 30× mean coverage, ≥ 90% of bases at 20× coverage, and ≥ 8 × 10^10^ aligned Q30 bases; however, due to an error in the AoU quality control process, the released samples exhibited ≥ 23.77× mean coverage, ≥ 78% of bases at 20× coverage, and ≥ 8 × 10^10^ aligned Q30 bases instead for the EUR-like ancestry (1).

For this analysis, the variant set generated by AoU, which was processed using hard threshold filters and allele-specific variant quality score recalibration, was used. The variants were only included in the final set if they passed all the hard threshold filters: high-quality genotypes, such as a genotype quality (GQ) of at least 20, a read depth (DP) of at least 10, and an allele balance (AB) of at least 0.2 for heterozygotes (GQ ≥ 20, DP ≥ 10, and AB ≥ 0.2 for heterozygotes); ExcessHet (defined as Phred-scaled p-value for exact test of excess heterozygosity) value below 54.69 (corresponding to filtering out variants with a z-score ≤ -4.5 or a p-value ≤ 3.4e-06 for this test); QUAL greater than or equal to 60 for SNVs (QUAL ≥ 60), and sites with less than or equal to 100 alternate alleles. The AoU variant quality control procedures also implemented the Variant Extract-Train-Score method, filtering out variants based on specific features. These features included Variant Confidence/Quality by Depth (AS_QD), Z-score from the Wilcoxon rank sum test of alternative vs. reference read mapping qualities (AS_MQRankSum), Z-score from the Wilcoxon rank sum test of alternative vs. reference read position bias (AS_ReadPosRankSum), Phred-scaled p-value using Fisher’s exact test to detect strand bias (AS_FS), RMS Mapping Quality of reference vs. alt reads (AS_MQ), and Symmetric Odds Ratio of a 2x2 contingency table to detect strand bias (AS_SOR). All variants that failed the Variant Extract-Train-Score method were excluded from this analysis (1).

After sample and variant quality control, AoU flagged samples that failed one of the following quality control hard thresholds and were identified as outliers: number of SNVs < 2.4M or > 5.0M, number of variants not present in Genome Aggregation Database (gnomAD) 3.1 > 100K, and heterozygous-to-homozygous ratio > 3.3. However, no samples were flagged based on these hard thresholds by the AoU.

In addition to the hard thresholds, samples were flagged as population outliers in one of the WGS quality control metrics for an assigned ancestry. Instead of using the AoU ancestry-outlier–flagged samples based on AoU’s ancestry assignments, ancestry assignments described in the *Ancestry Assignment* section were used to identify population outliers. This approach was taken to ensure a consistent definition of ancestry across all analyses.

Accordingly, samples were flagged and subsequently removed from the analysis if one of the WGS metrics was greater than eight standard deviations from the ancestry-specific mean (9,10). These WGS metrics included number of deletions, number of insertions, number of SNVs, number of variants not present in gnomAD 3.1 (singletons), insertion-to-deletion ratio, transition-to-transversion ratio, SNV heterozygous-to-homozygous ratio, and indel heterozygous-to-homozygous ratio. This approach flagged 300 participants in total. This is far fewer than the 987 (0.2%) samples that were flagged as outliers by the AoU using the AoU ancestry assignments (1). However, after removing all samples for quality control metrics, sex discordance, or ancestry assignment outlier (greater than 3 standard deviations from the ancestry median), both approaches functionally excluded approximately the equivalent number of participants from each ancestry assignment.

The above quality control for WGS was primarily performed by AoU. At this stage, samples flagged as population outliers and individuals with discordant genetically and self-reported sex (participants who reported “Intersex,” “Prefer not to answer,” “None of these fully describe me,” or who skipped the question) were excluded from the analysis dataset (1,6,11). Samples that passed these quality control procedures were checked for sample missingness. The following procedures were applied to both the ACAF Threshold WGS and Exome WGS datasets separately. Variants were removed if they had a call rate of < 90%, and the remaining variants were then used to calculate sample missingness. Samples with a call rate below 90% were flagged in each dataset and subsequently removed from all analyses using that dataset (6,11).

### Ancestry assignment

After microarray quality control, as described above, ancestry assignment was conducted by implementing the *POP-MaD* (population grouping by Mahalanobis distance) pipeline (12,13). This was performed by estimating ancestry principal components from an external reference panel consisting of the merged 1000 Genomes Project (KGP) and Human Genome Diversity Project (HGDP) datasets (14). Microarray samples from AoU were projected onto the KGP and HGDP principal components using smartPCA (15). Using 10 ancestry principal components, AoU samples were then assigned to ancestry assignments based on the minimum Mahalanobis distance. Ancestry outlier samples were removed by restricting assignment to those within 3 standard deviations of the reference group’s median Mahalanobis distance (13). Consequently, 82,675 samples were assigned to the African-like (AFR-like), 94,245 to the Admixed American-like (AMR-like), 7,972 to the Central/South Asian-like (CSA-like), 10,009 to the East Asian-like (EAS-like), 239,578 to the European-like (EUR-like), 2,687 to the Middle Eastern-like (MID-like), and 60 to the Oceanian-like (OCE-like) (12).

After ancestry assignment, within-ancestry principal components were computed using FlashPCA 2.0 for samples with microarray data available in the AoU (16). Quality control and linkage disequilibrium pruning were performed separately within each ancestry assignment. Principal components were calculated using an unrelated subset of participants (defined as having less than third-degree relatedness, estimated using KING) within each ancestry assignment (17). Related individuals were then projected onto the computed principal components (18). In this study, we restricted our analyses to the largest ancestry group (EUR-like), including both related and unrelated individuals.

### Exome variant quality control

Variant quality control for the Exome WGS dataset was performed within the EUR-like ancestry assignment after excluding ancestry-outliers and participants who failed the WGS quality control procedures (9,11). After excluding these samples, variant quality control procedures previously used in several rare variant analyses were applied to the Exome WGS dataset; variants with a call rate less than 90% (variant call rate < 90%), Hardy–Weinberg equilibrium p-value less than 1 × 10^-15^ (p-value < 1 × 10^-15^), presence in low-complexity regions, and monomorphic variants (minimum MAC cutoff of 1) in the quality-controlled cohort were subsequently removed (9,11,19–22).

### ACAF variant quality control

Variant quality control for the ACAF Threshold WGS dataset was performed within the EUR-like ancestry assignment after excluding ancestry-outliers and participants who failed the WGS quality control procedures. After excluding these samples, variants were filtered based on the following criteria: call rate of less than 90% (variant call rate < 90%), Hardy–Weinberg equilibrium p-value less than 1 × 10^-10^ (p-value < 1 × 10^-10^), and MAF < 1% (10,11,23).

### Sensitivity analysis for deprivation index

After performing the initial gene set-based analysis with the standard set of covariates (age, sex, principal components), both survey and EHR data were examined for potential risk factors, confounders, and EHR accessibility indicators to include as additional covariates in set-based tests. These factors included smoking history, prior exposure to alkylating agents, and the deprivation index (24,25).

Examined covariates are described as follows:

- Smoking: Any endorsement of smoking history in either the EHRs or surveys (concept identifiers: 1332800, 1332802, 1585857, 1585858, 1586166, 1586167, 1586174, 1586175, 1586182, 1586183, 1586190, 1586191, 1762678, 4011455, 4021160, 4021161, 4034855, 4044698, 40482909, 4052779, 40766360, 40766642, 40766646, 4090847, 4132133, 4188539, 4191706, 4193014, 4206526, 4219336, 4224317, 4226233, 4227477, 42709996, 4276526, 4282779, 4298794, 4301349, 43054909, 4308357, 4310250, 44783000, 45765448, 45878118, 45881517, 45883458, 45884038, 45890719, 46272595, 46272634, 46273821, 765320, 763760, 762498, 762499, 903652, 903654, 903656, 903657, 903659, 903661, 915749, 36715277, 36716475, 37395605).
- Alkylating agent status: EHR documentation of alkylating agent exposure prior to AML diagnosis in the EHRs (concept identifier: 21601388).
- Observation period: Time from the first EHR to the last EHR for a participant.

As an initial test, the burden score for each gene of the top three significant genes–*DNMT3A*, *TET2*, *SRSF2*–from the gene set-based analysis was computed. We calculated burden scores by summing the rare predicted deleterious variants (MAF < 1%) identified in the gene. Predicted deleterious variants were defined using the same criteria as in the gene set–based analysis (either LoF or damaging missense). Logistic regression models (‘stats’ R package) were fitted separately for each gene burden score, with the gene burden score as a predictor of the AML phenotype (Table S25) (26). Other covariates for the logistic regression models included the total count of rare variants across the exome (including both protein-coding and non-coding), sex, age, and the first ten ancestry-specific principal components. All quantitative predictors were standardized (27). For the sensitivity analysis, an additional covariate was included in the model (smoking history, prior exposure to alkylating agents, and the deprivation index). Each potential covariate was tested separately and represents a separate model.

Additionally, using the same methodology, tests were conducted using a Cox regression model (‘survival’ R package) using the same covariates as specified above (Table S26) (28). Both the time from birth to the first EHR corresponding to AML, as well as the time from the start of the observation period (i.e., the time of the first EHR) to the first EHR corresponding to AML, were modeled.

For these tests, the p-value of the gene burden score as well as the effect sizes of the covariates were examined. The covariate was selected for inclusion in the gene set-based tests if the significance of the gene burden score consistently increased across all genes and did not change the direction of the effect size of the gene burden score.

Based on these criteria, we tested whether the deprivation index could improve the significance of the gene set-based results (Tables S4-S5, S8-S9). Other than this modification, the same configuration as that used in the initial set-based tests was retained, including the sparse GRM, null model parameters, variant annotations, and the variants included in the variant set. As part of evaluating the impact of including the deprivation index as a covariate, we examined genomic inflation (Figures S14-S15, Table S1) and the burden-test beta estimates (Figure S16, Tables S4-S5 and S8-S9), comparing these against the SAIGE-GENE+ set-based test without including the deprivation index. Additionally, we assessed -log10 p-value correlations to demonstrate that this inclusion increased the overall significance of our results (Figure S17, Tables S4-S5 and S8-S9). Finally, to ensure that the individual components of the deprivation index (vacant housing, poverty, lack of health insurance, median income, and high school education) did not contain additional information that could further improve gene identification beyond the composite deprivation index, an additional association test was performed adjusting for the components of the deprivation index (Figures S18-S21, Tables S27-S30). For the gene set-based tests, both the deprivation index and its component variables were standardized.

### Sensitivity analysis for age definition

In addition to our other sensitivity analyses, we performed a supplemental analysis in which the definition of age was modified. In the initial set-based tests, age was included as one of the standard covariates in the SAIGE-GENE+ and was defined as age at the last EHR encounter. This was done to account for the full duration of the observation period for each individual. We then assessed alternative definitions of age: age at biospecimen collection (defined as the age at which a whole-genome sequencing sample was collected) and age at diagnosis (defined as age at first EHR corresponding to AML for cases and age at the last EHR encounter for controls). For both analyses, the modified definition of age, along with the other standard covariates—including sex and the first 10 principal components—was included in the set-based tests. Other than this modification to the age definition, the same configuration as that used in the initial set-based tests was retained, including the sparse GRM, null model parameters, variant annotations, and the variants included in the variant set.

Although using age at diagnosis yielded stronger significance for the top genes compared with using age at the last EHR encounter, the overall differences across age definitions were negligible (Figures S22-S25, Tables S31-S38). The -log10 p-values and burden coefficient correlations remained high (r > 0.998) across different definitions of age (Figures S14-S15). Given this high concordance, age at the last EHR encounter was deemed an appropriate definition of age for our primary analysis. The results of these tests are included in the Supplementary Tables.

### Age at biospecimen collection

We also examined age at biospecimen collection in greater detail as a potential confounder. Specifically, somatic variation may accumulate with increasing age at biospecimen collection, potentially biasing our results (6,29). We therefore conducted a whole-exome association study using inverse-normalized age at biospecimen collection as the phenotype to assess whether age was associated with an increased burden of rare variants. Set-based tests were performed adjusting for sex and the first 10 within-ancestry principal components in the same cohort used for the AML set-based tests. The null model specifications, variant annotations, and variants set were identical to that used in the AML set-based tests. This analysis identified several genes as exome-wide significant (adjusted p-value < 0.05), including several genes also identified in the AML set-based tests (*DNMT3A*, *TET2*, and *SRSF2*) (Figures S26-S27, Tables S39-S42). Given these results, we further explored the findings at the single-variant level.

Next, rare single-variant tests were conducted to identify rare variants whose frequency is associated with age at biospecimen collection, potentially representing mutational hotspots. These tests were performed within genes that were significant in the AML set-based tests (*DNMT3A*, *TET2*, *SRSF2*, and *IDH2*), using age at inverse normalized biospecimen collection as the outcome and adjusting for sex and 10 principal components within SAIGE-GENE+. Variants were included if they passed exome WGS quality control and had a cohort-specific MAC > 0 and MAF < 0.01. When p-values from the initial test were < 0.05, Fifth regression was applied to calculate more accurate p-values. Additionally, for variants with MAC ≤ 10, efficient resampling was applied to compute p-values. The null model specifications were identical to those used in the rare set-based tests for age at biospecimen collection. Genome-wide significance was defined as P < 5 × 10^-8^ (Table S43). The results of these tests are included in the Supplementary Tables.

### Age at biospecimen collection matching

To address potential confounding that persisted after conditioning, cases and controls were matched based using their age at biospecimen collection. We used the *MatchIt* R package’s nearest neighbor matching method to match controls to cases (30). Using *MatchIt* to subsample the original cohort, we generated multiple matched cohorts with different case–control matching ratios, including 1:5 (N_cases_=265 and N_controls_=1325), 1:10 (N_cases_=265 and N_controls_=2650), 1:50 (N_cases_=265 and N_controls_=13,250), 1:100 (N_cases_=265 and N_controls_=26,500), and 1:300 (N_cases_=265 and N_controls_=79,500). As recommended in the *MatchIt* documentation, matching balance was initially assessed using standardized mean differences, variance ratios, and empirical cumulative distribution function (eCDF) statistics generated by MatchIt (30). Across all case-control matching ratios, standardized mean differences and eCDF statistics were close to zero, while variance ratios were close to one (all values ≤ 0.0003, 0.0094, and 1.0047, respectively), indicating good matching balance (Table S44).

Next, after matching, rare single-variant association tests were conducted to determine whether matching cases to controls affected the effect sizes of individual rare variants. (This approach was not feasible for set-based tests because matching alters cohort-specific MAF estimates used for weighting rare variants, consequently affecting effect sizes and statistical significance.) For the single-variant tests, these tests were performed within genes that were significant in the AML set-based tests (*DNMT3A*, *TET2*, *SRSF2*, and *IDH2*), using binary AML phenotype as the outcome and adjusting for sex, age at biospecimen collection, and 10 principal components within SAIGE-GENE+. Variants were included if they passed exome WGS quality control and had a cohort-specific MAC > 0 and MAF < 0.01. When p-values from the initial tests were < 0.05, Firth regression was applied to obtain more accurate p-values.

Additionally, for variants with MAC ≤ 10, efficient resampling was used to calculate p-values. The null model specifications were identical to those previously used for AML phenotypes. We then examined the correlation of effect sizes for rare variants (0.0001 < MAF < 0.01) to ensure that the matching procedure did not alter effect size estimates across different matching ratios (Figure S28, Table S45). (Below this threshold, restricting the control set frequently reduces the MAC below the level at which SAIGE-GENE+ can accurately estimate effect sizes.)

We then conducted set-based tests for genes significant in the primary AML analyses (*DNMT3A*, *TET2*, *SRSF2*, and *IDH2*) using both the binary AML phenotype and quantitative age at biospecimen collection as phenotypes in the biospecimen collection–matched cohorts. For the AML phenotype analyses, models were adjusted for age at biospecimen collection, sex, and the first 10 principal components. For the age at biospecimen collection analyses, inverse-normalized age at biospecimen collection was used as the phenotype, and set-based tests were performed adjusting for sex and the first 10 within-ancestry principal components. The same configuration as that used in the initial set-based tests was retained for both analyses, including the sparse GRM, null model parameters, variant annotations, and the variants included in each gene-based set.

These set-based tests aimed to show that increasingly restrictive case-control matching produced minimal attenuation in AML associations but substantial attenuation in associations with age at biospecimen collection as the phenotype, indicating that the matching procedure effectively controlled for age-related effects without materially affecting AML signals. This pattern was reflected in the results: as the control set became more restrictive, −log10 p-values for age at biospecimen collection declined approximately linearly, whereas AML associations showed minimal attenuation (Figure S29, Table S46-S47). A marked reduction in AML significance was only observed at a 1:5 case-control ratio, where cohort size was substantially reduced and cohort-specific MAF weights used in the set-based tests were likely altered. Together, these findings suggest that, while age-related somatic burden cannot be fully excluded, the observed associations between rare deleterious variation and AML are primarily attributable to germline variation rather than somatic mutational burden.

### Cross referencing

For the genes identified from the gene set-based tests, variants within the variant sets were cross-referenced with variants from the GDC and ClinVar databases (31,32). For this, AoU variant identifiers were translated into Human Genome Variation Society (HGVS) nomenclature to enable cross-referencing with GDC somatic alterations (33). The proportion of overlapping variants is reported for rare variants (MAF < 1% and MAC ≥ 10) and ultra-rare variants (MAC < 10), considering overlap with either the GDC or ClinVar databases, or both (Figure S30-S31, Table 48). Additionally, the proportion of overlapping variants is reported stratified by annotation tools used to designate variant function (LOFTEE, SpliceAI, or AlphaMissense) (Figure S32-S33, Table 48).

Finally, to assess whether a proportion of the identified variants have previously established associations with AML or AML-related phenotypes, the phenotypes corresponding to the cross-referenced AoU variants in ClinVar were examined. The counts of associations with variants present in the identified genes, for rare variants (MAF < 1% and MAC > 10) and ultra-rare variants (MAC ≤ 10), respectively, were reported (Figure S34-S35).

### Global burden gene-sets: Somatic mutation gene-sets

The frequency of somatic mutations in protein coding genes was obtained from the GDC (31). The first set of gene-sets was derived from the somatic mutation frequency across all cancer types in the GDC, encompassing 16,747 participants with available open-access somatic mutation frequency data. All genes included in the following gene-sets were identified by the Cancer Gene Census as high-confidence cancer-driving genes (34). The gene-sets included (Table S13):

- Protein-coding genes with at least one somatic mutation
- Protein-coding genes with at least one somatic mutation predicted to have moderate or high impact by VEP
- Protein-coding genes with at least one somatic mutation predicted to have high impact by VEP

The second set of gene-sets was derived from the somatic mutation frequency across AML in the GDC, encompassing 571 participants with available open-access somatic mutation frequency data. All genes included in the following gene-sets were identified by the Cancer

Gene Census as high-confidence cancer-driving genes (34). The gene-sets included (Table S13):

- Protein-coding genes with at least one somatic mutation
- Protein-coding genes with at least one somatic mutation predicted to have moderate or high impact by VEP
- Protein-coding genes with at least one somatic mutation predicted to have high impact by VEP

### Global burden gene-sets: Cox regression gene-sets

Bulk RNA-seq gene-level unstranded raw counts processed using the GDC mRNA Analysis Pipeline were obtained from the GDC for participants diagnosed with AML: 2,000 bone marrow samples of “primary tumor,” 599 peripheral blood samples of “primary tumors,” and 324 bone marrow samples from “normal tissue” (31). Within each biospecimen type, genes with raw counts less than 10 were removed, and the remaining genes were transformed using the variance-stabilizing transformation from DESeq2 (35). For each protein-coding gene, a separate Cox regression (‘survival’ version 3.8.3 R package) was performed to predict the time from diagnosis to vital status at last follow-up (28,36,37). Each Cox regression included the variance-stabilizing transformed counts for a given gene, age at diagnosis, and reported gender as predictors. Quantitative covariates, including both age at diagnosis and gene counts, were standardized to have a mean of 0 and a standard deviation of 1. Batch information was not available from the GDC and therefore could not be included as a covariate for batch correction in the statistical model. Instead, the GDC harmonizes RNA-seq data across projects using the GDC RNA-Seq Expression Pipeline, which standardizes data processing and reduces technical variability (31).

After performing the Cox regressions, the coefficient p-values corresponding to each gene were Bonferroni-corrected for multiple testing, and all significant protein-coding genes with an adjusted p-value < 0.05 were included in the gene-set (Table S14) (38). Additionally, genes upregulated in individuals with a higher risk of mortality (hazard ratio [HR] > 1) and genes downregulated in individuals with a lower mortality risk (HR < 1), using the same

Bonferroni-corrected p-value threshold (adjusted p-value < 0.05), were used to define two additional gene-sets: one consisting of genes whose upregulation was associated with increased mortality, and another consisting of genes whose upregulation was associated with decreased mortality.

### Global burden gene-sets: Differential gene expression gene-sets

Bulk RNA-seq gene-level unstranded raw counts processed using the GDC mRNA Analysis Pipeline were obtained from the GDC for participants diagnosed with AML: 2,000 bone marrow samples of “primary tumor” and 324 bone marrow samples from “normal tissue” (31). Within each biospecimen type, genes with raw counts less than 10 were removed, and the remaining genes were transformed using the variance-stabilizing transformation from DESeq2 (35). Differential gene expression was then conducted using DESeq2 comparing affected versus unaffected bone marrow adjusting for gender and age. Age was standardized to have a mean of 0 and a standard deviation of 1. An absolute log2 fold change greater than or equal to 0.58 and a Bonferroni-corrected p-value < 0.05 were used to identify protein-coding genes for inclusion in the gene-set (Table S15) (38). Additionally, we subdivided the differentially expressed gene-set into upregulated and downregulated gene-sets. The upregulated gene-set included genes with a log2 fold change > 0.58 and an adjusted p-value < 0.05, whereas the downregulated gene-set included genes with a log2 fold change < −0.58 and an adjusted p-value < 0.05.

### Global burden gene-sets: Single-cell differently expressed gene-sets

Single-cell differentially expressed gene-sets were generated by performing differential gene expression analysis for each annotated cell type in GSE116256 (39). To obtain cell type–specific gene-sets in adult AML at diagnosis (prior to treatment), a subset of data from GSE116256 was used, comprising 16 AML bone marrow samples collected at diagnosis and 5 healthy bone marrow samples. These samples had already undergone quality control and cell type annotation before being deposited in the Gene Expression Omnibus (40). The quality control steps required that cells have at least 1,000 UMIs, at least 500 unique genes, and less than 20% of total gene counts derived from mitochondrial or ribosomal RNA genes (39). In addition to these quality control measures, after normalizing each sample with SCTransform in Seurat (version 5.3.0), DoubletFinder (version 2.0.6) was applied to each sample independently to identify doublets. DoubleFinder was applied using an expected doublet rate of 0.5734%, which was calculated based on the Poisson distribution (41–43). After removing predicted doublets, the samples were integrated into a single dataset using Harmony with 30 principal components to examine the prior cell type annotations from GSE116256 (39,44). Differential gene expression analysis of single-cell data was performed using MAST within the Seurat FindMarkers function (42,45). Only cells classified as either ‘malignant’ or ‘normal’ were included in the differential gene expression analysis, while cells classified as ‘unclear’ were excluded. Multiple tests were conducted, varying which cell types were designated as cases or controls and whether cells from healthy bone marrow samples were included in the analysis. These included:

- Malignant cells vs unaffected cells in AML samples
- Malignant cells vs unaffected cells in both AML affected and unaffected healthy bone marrow
- Single cell type contrasted against all other cell types in AML samples
- Single cell type contrasted against all other cell types in AML samples and unaffected healthy bone marrow

Analyses that included only AML-affected bone marrow used sample ID, age, and sex as covariates. In analyses that included both AML-affected and unaffected bone marrow, only age and sex were used as covariates. Differential gene expression analyses in single-cell RNA-seq typically yield a larger number of significant genes than bulk RNA-seq, so a more stringent significance threshold was applied. A Bonferroni-corrected p-value < 0.01 and an absolute average log2 fold change of at least 0.58 were used to identify protein-coding genes for inclusion in their respective gene-sets (Table S16) (38). Additionally, we subdivided the single-cell differentially expressed gene-sets into upregulated and downregulated gene-sets. The upregulated gene-set included genes with an average log2 fold change > 0.58 and an adjusted p-value < 0.01, whereas the downregulated gene-sets included genes with an average log2 fold change < −0.58 and an adjusted p-value < 0.01.

### Limitations

Differences in quality control procedures can lead to different variants being included in the set-based tests, which ultimately affects the significance of the set-based tests. The quality control pipeline developed by the team from the AoU attempts to accurately call variants from technical artifacts, such as contamination; however, the results in the quality controlled variants can be composed of a mixture of germline variants and somatic mutations accumulated through clonal hematopoiesis or chimerism (1,10,46,47). Combined with our findings from the set-based tests using age at biospecimen collection as a phenotype, these results suggest that some of the variants identified in the AML set-based tests likely represent a mixture of germline variants and somatic mutations. This does not undermine our primary findings, which establish associations between AML and the accumulation of deleterious rare variants, as these variants are potential drivers of disease whether germline or somatic.

The AoU released 4,044 CDRv8 samples that did not meet the WGS coverage metric of ≥30× mean coverage and were erroneously included in the callset ([Genomic Research Data Quality Report](https://support.researchallofus.org/hc/en-us/article_attachments/40491583732116)) (1). These samples were included during AoU variant quality control, affecting which variants were present in the Exome and ACAF WGS datasets. Although there were options to exclude these samples, given that they were already included in the AoU variant quality control, the decision was made to retain them in the analysis. Excluding these samples could potentially alter the results.

In the current manuscript, it was assumed that synonymous variants do not significantly contribute to disease risk and, therefore, can be used as an empirical negative control for p-value thresholding. However, synonymous variants have previously been reported to be associated with (1) hematological malignancies, most notably synonymous variants in immunoglobulin lambda like polypeptide 5 (*IGLL5*) associated with chronic lymphocytic leukemia and (2) other complex traits (7,48–50). These associations are unlikely to reflect a functional effect of the synonymous variants themselves and therefore should not be interpreted as evidence of a causal relationship with hematological malignancies.

In our analysis, EHRs were used as the phenotyping instrument to define cases and controls; however, the EHRs within the AoU do not represent a complete medical history of participants. The AoU includes only EHRs from health systems that cooperate with the program, so it is possible that participants have additional records in other systems. Furthermore, no EHR data exist prior to 1981, meaning that for some individuals, a portion of their life is unobserved.

For the current manuscript, all AML cases were analyzed together as a single phenotype. This approach does not account for within-phenotype heterogeneity, such as differences in age of onset (pediatric vs. adult AML) or AML subtypes.

We focused on the contribution of rare and common variants to AML; however, these types of variants do not encompass the full range of genomic architectures that could predispose individuals to AML, such as chromosomal translocations and structural variants. Further analyses are needed both to explore the contributions of these additional genomic features and to integrate all aspects of genomic architecture into a single comprehensive analysis.

## Supplementary Figures

**Supplementary Figure 1.**
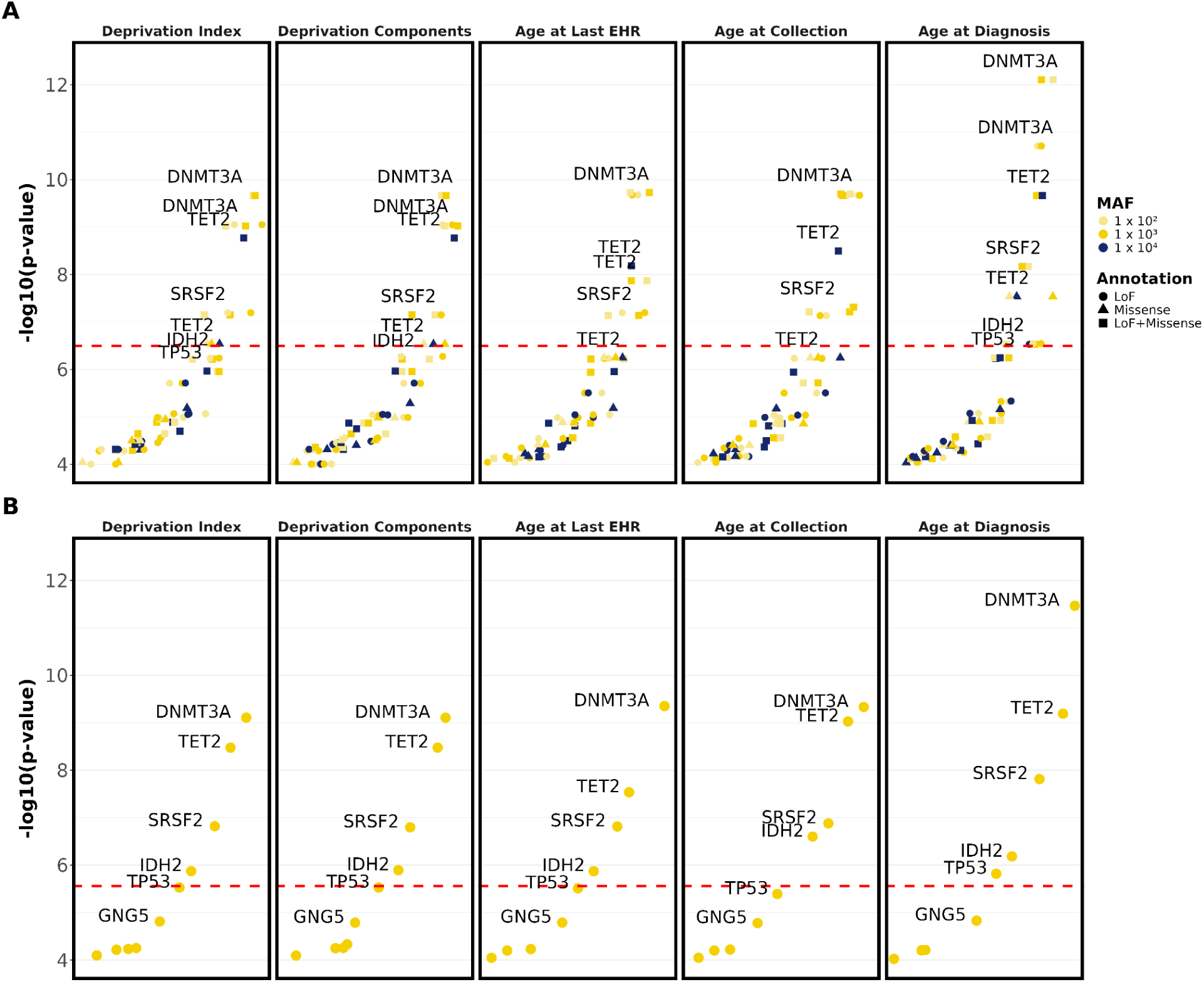
Top genes for AML set-based tests. P-values of top genes from gene set-based tests are shown for SKAT-O analyses using non-Cauchy and Cauchy methods, stratified by analysis type. All displayed genes have p-values ≤ 1 × 10^-4^. The top row (A) shows non-Cauchy SKAT-O p-values, and the bottom row (B) shows Cauchy SKAT-O p-values. The red dashed line represents genome-wide significance after multiple hypothesis correction. “Age at Last EHR” is the baseline model using standard covariates (age, sex, principal components). “Deprivation Index” and “Deprivation Components” include additional socioeconomic covariates beyond the standard covariates, where the Deprivation Index model includes the composite deprivation score and the Deprivation Components model includes its individual components. “Age at Collection” and “Age at Diagnosis” use the standard set of covariates; however, they differ from the baseline analysis in how age is defined. “Age at Collection” uses time from birth to biosample collection, and “Age at Diagnosis” uses age at diagnosis for cases and age at last EHR for controls.

**Supplementary Figure 2.**
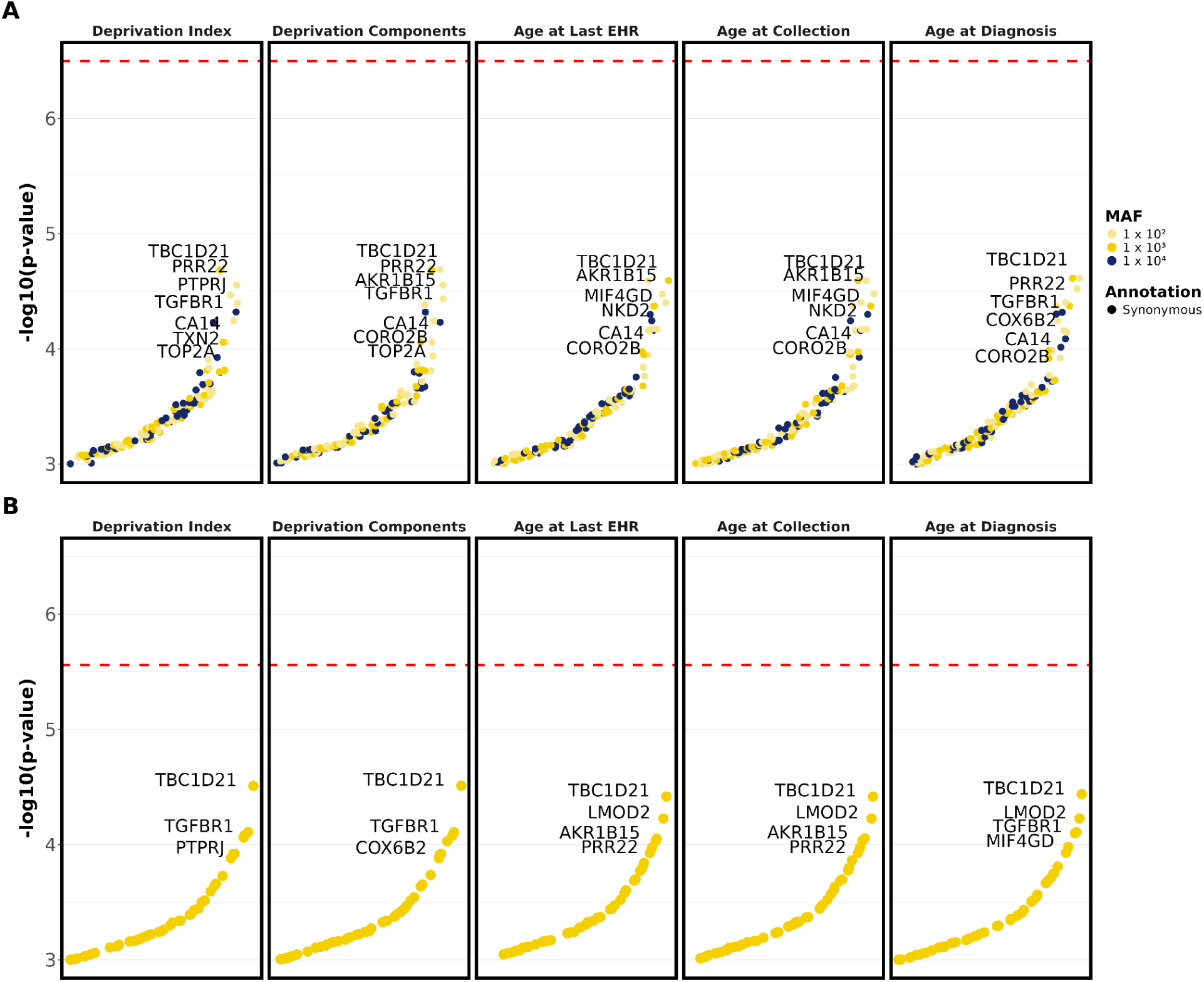
Top genes for AML synonymous set-based tests. P-values of top genes from synonymous gene set-based tests are shown for SKAT-O analyses using non-Cauchy and Cauchy methods, stratified by analysis type. All displayed genes have p-values ≤ 1 × 10^-3^. The top row (A) shows non-Cauchy SKAT-O p-values, and the bottom row (B) shows Cauchy SKAT-O p-values. The red dashed line represents genome-wide significance after multiple hypothesis correction. “Age at Last EHR” is the baseline model using standard covariates (age, sex, principal components). “Deprivation Index” and “Deprivation Components” include additional socioeconomic covariates beyond the standard covariates, where the Deprivation Index model includes the composite deprivation score and the Deprivation Components model includes its individual components. “Age at Collection” and “Age at Diagnosis” use the standard set of covariates; however, they differ from the baseline analysis in how age is defined. “Age at Collection” uses time from birth to biosample collection, and “Age at Diagnosis” uses age at diagnosis for cases and age at last EHR for controls.

**Supplementary Figure 3.**
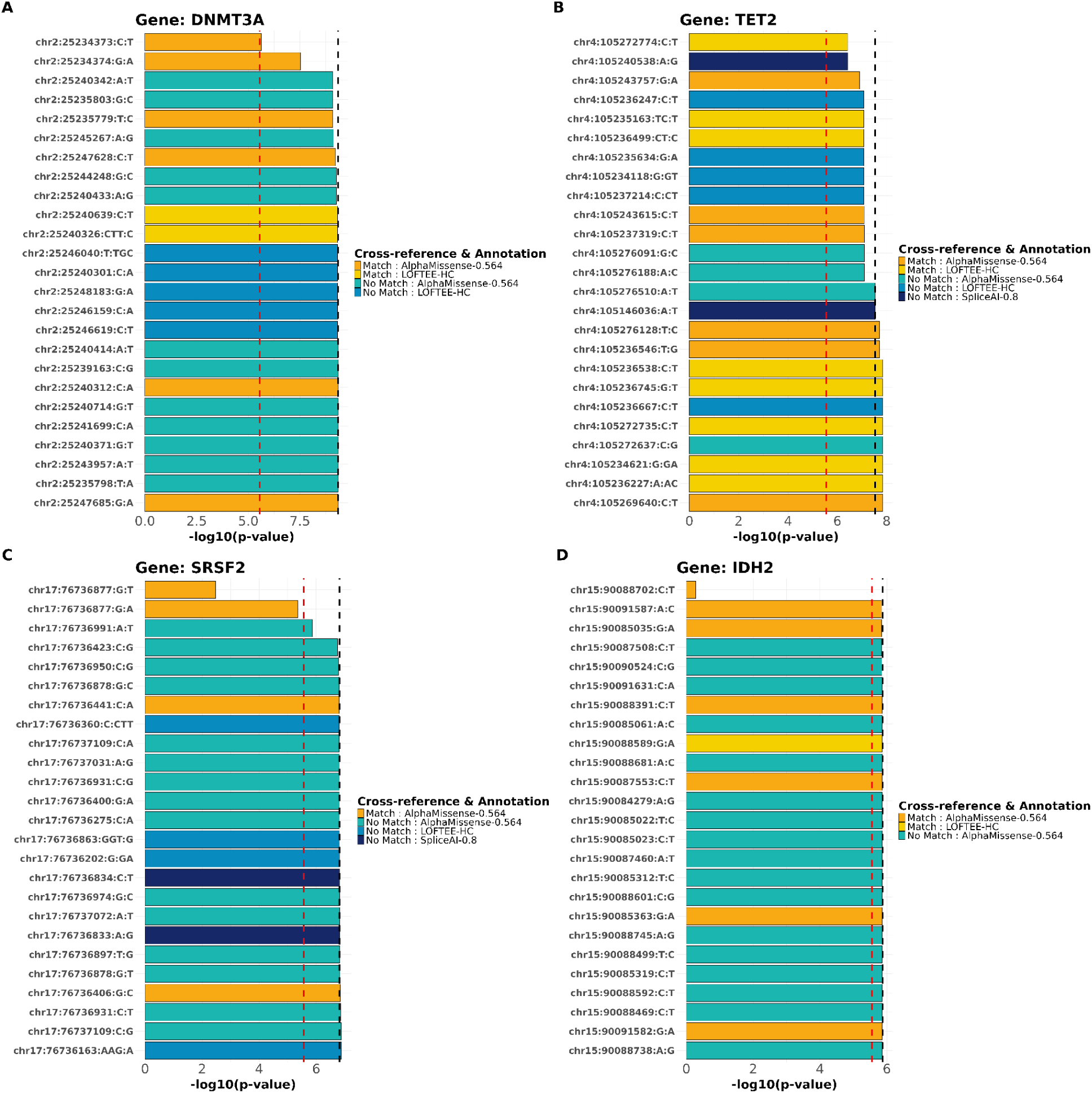
Leave-one-variant-out tests for AML. Bar plot of Cauchy SKAT-O p-values from the leave-one-variant-out analysis of AML using standard covariates, displayed as -log10 p-values. The black dashed line represents the p-value from the standard analysis, where no variants are removed from the gene-based test. The bars display the p-values obtained after removing one variant at a time in the leave-one-variant-out analysis. The red dashed line indicates the genome-wide significance threshold. A reduction in significance such that the bar lies between the black dashed line and the red dashed line suggests that the removed variant contributes to the gene-level signal but is not solely responsible for genome-wide significance. In contrast, if the bar falls below the red dashed line, this indicates that the removed variant is largely or solely driving the genome-wide significant association for the gene.

**Supplementary Figure 4.**
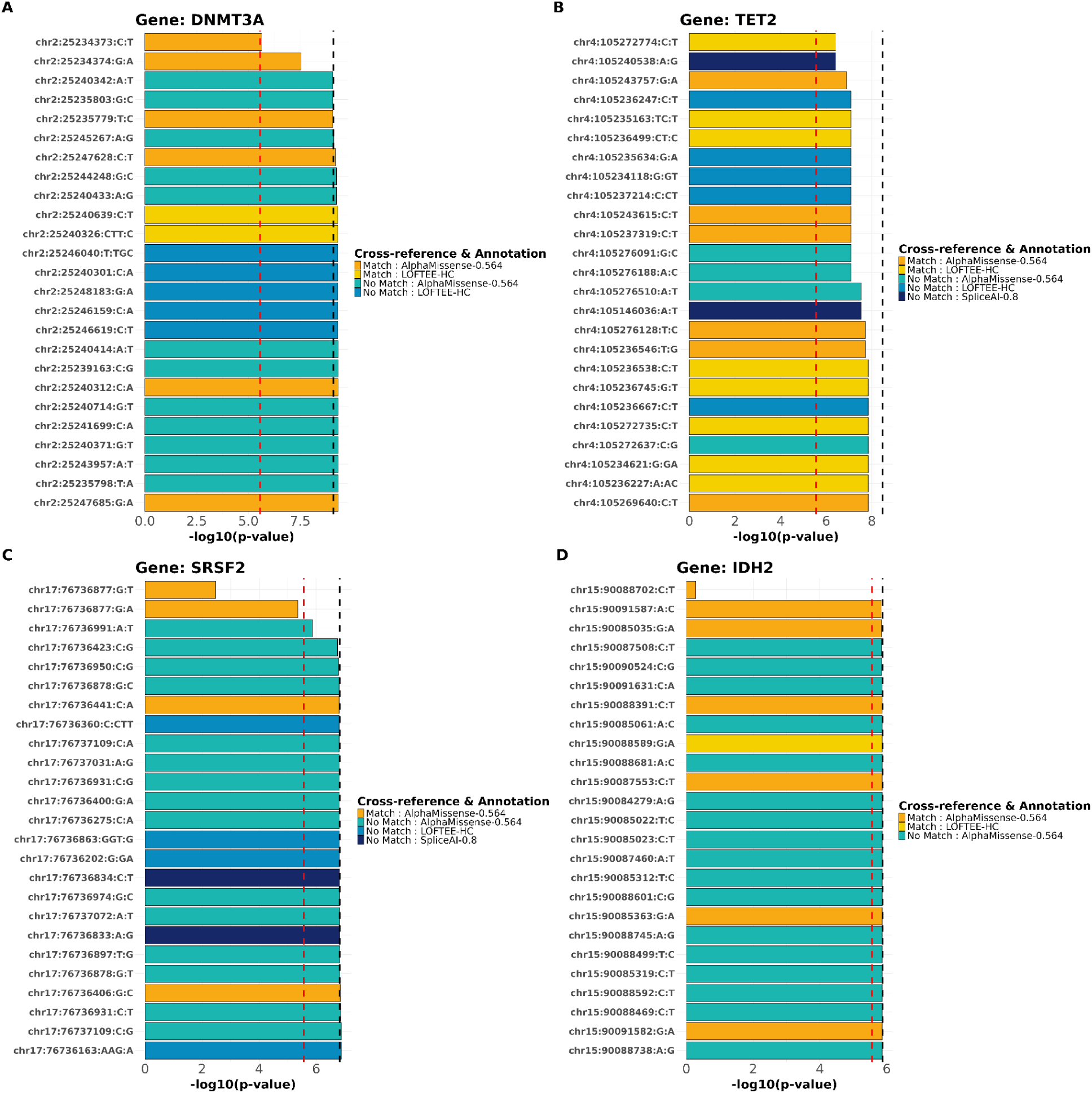
Leave-one-variant-out tests for AML adjusted for the deprivation index. Bar plot of Cauchy SKAT-O p-values from the leave-one-variant-out analysis of AML using standard covariates plus the deprivation index, displayed as -log10 p-values. The black dashed line represents the p-value from the standard analysis, where no variants are removed from the gene-based test. The bars display the p-values obtained after removing one variant at a time in the leave-one-variant-out analysis. The red dashed line indicates the genome-wide significance threshold. A reduction in significance such that the bar lies between the black dashed line and the red dashed line suggests that the removed variant contributes to the gene-level signal but is not solely responsible for genome-wide significance. In contrast, if the bar falls below the red dashed line, this indicates that the removed variant is largely or solely driving the genome-wide significant association for the gene.

**Supplementary Figure 5.**
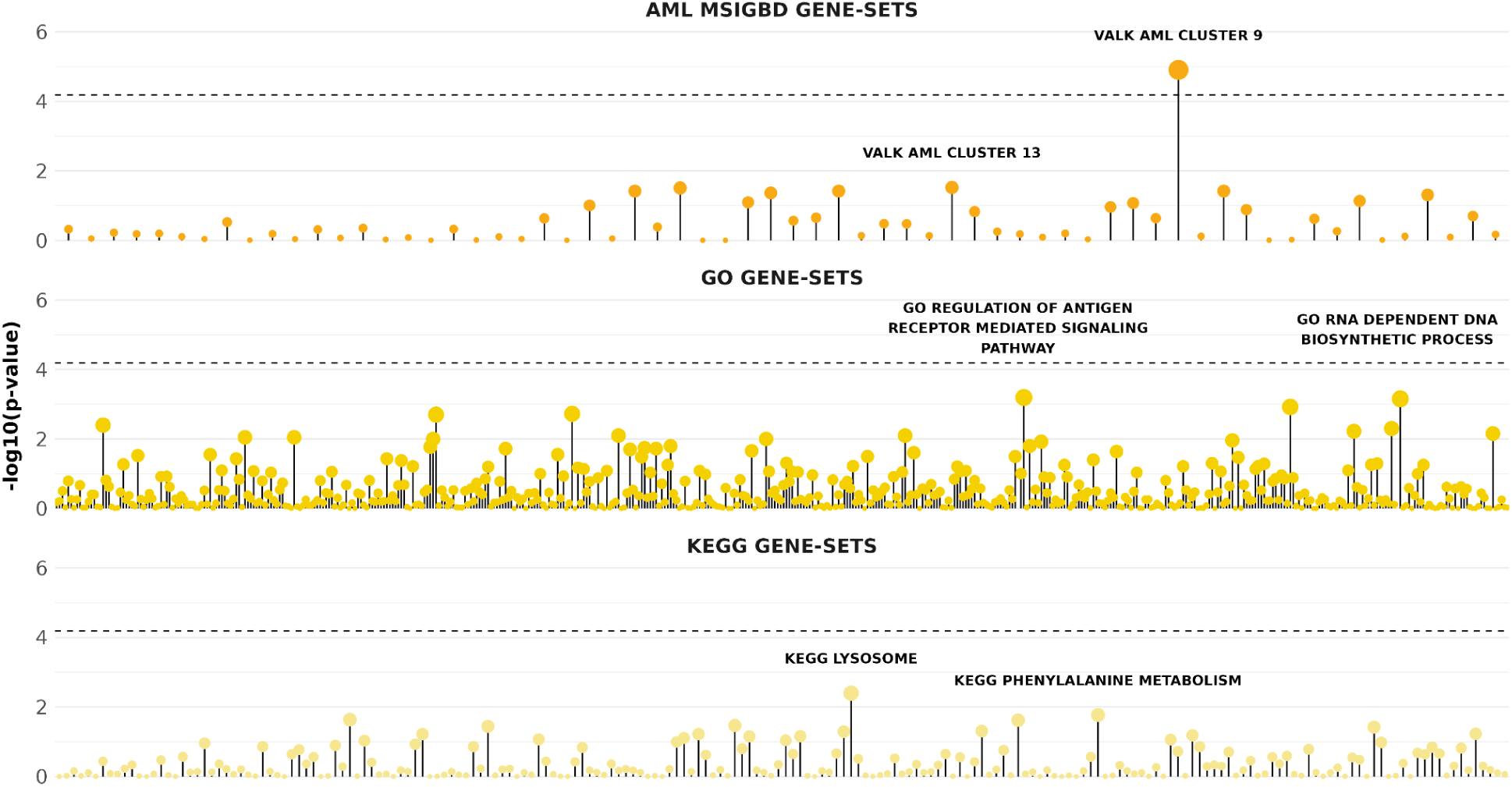
Gene-set results from GAUSS for MSigDB, GO, and KEGG. The plot displays -log10 p-values from gene-set association analyses conducted using GAUSS. The dashed black line represents the significance threshold after multiple hypothesis correction across all gene sets. Analyses were performed using both the GO (middle panel) and KEGG (bottom panel) pathway collections. Additionally, gene sets from the MSigDB were evaluated; however, due to overlap among MSigDB gene sets, the analysis was restricted to AML-related gene sets. In total, 64 AML-specific gene sets were tested (top panel).

**Supplementary Figure 6.**
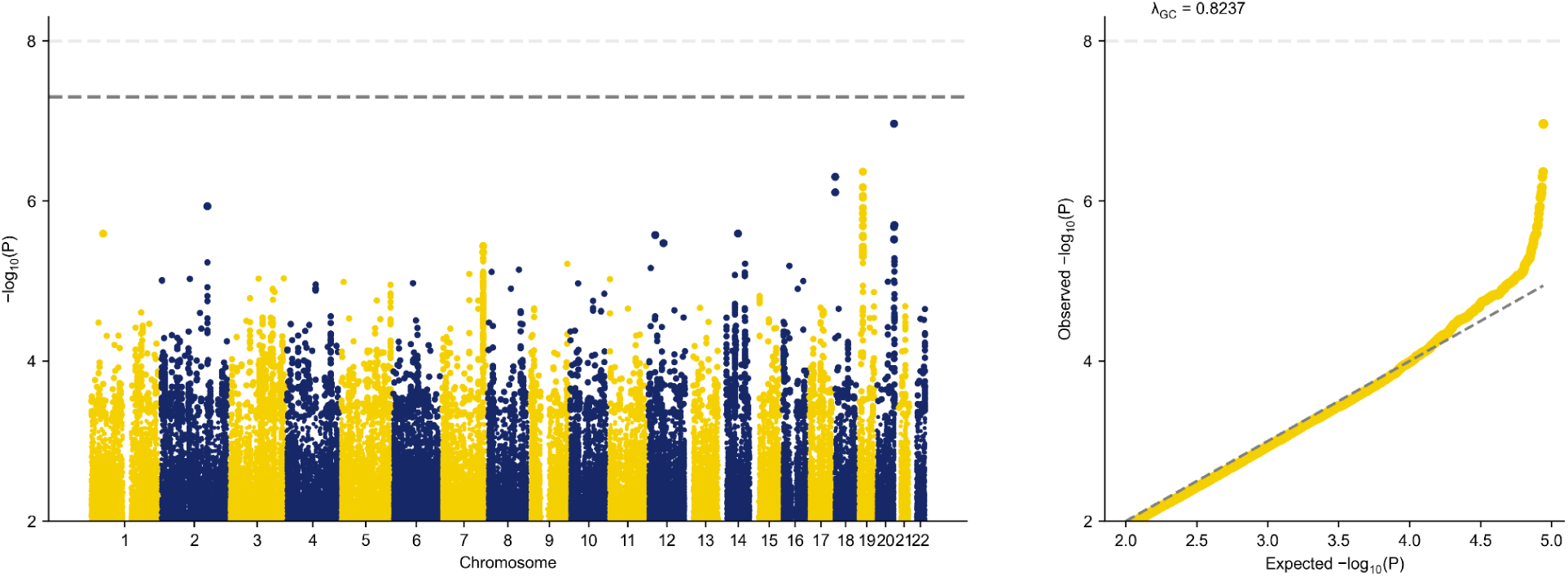
Single-variant tests for AML. Manhattan plot (left panel) and quantile-quantile (QQ) plot (right panel) for common variants (MAF > 1%) in AML, adjusted for the standard set of covariates. Genome-wide significance was defined as p < 5 × 10^-8^ and is indicated by the dashed light grey line. The QQ plot also illustrates the genomic inflation of the test statistics.

**Supplementary Figure 7.**
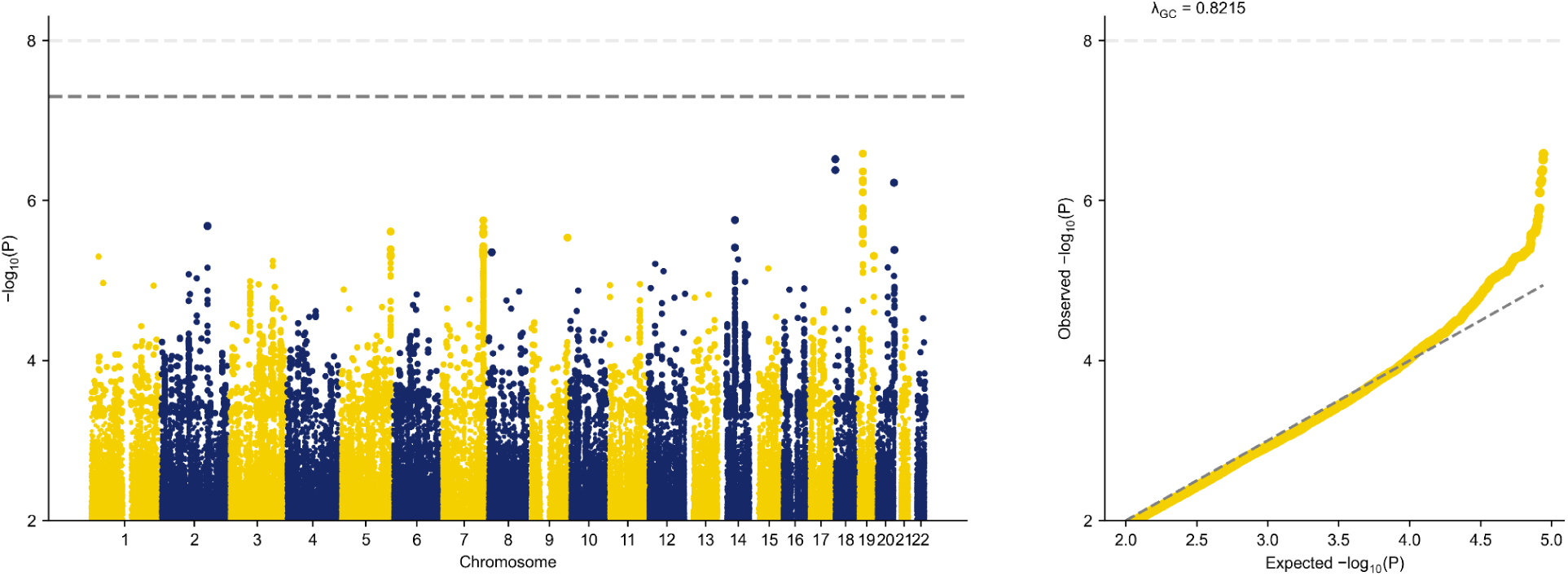
Single-variant tests for AML adjusted for the deprivation index. Manhattan plot (left panel) and quantile-quantile (QQ) plot (right panel) for common variants (MAF > 1%) in AML, adjusted for the standard set of covariates plus the deprivation index. Genome-wide significance was defined as p < 5 × 10^-8^ and is indicated by the dashed light grey line. The QQ plot also illustrates the genomic inflation of the test statistics.

**Supplementary Figure 8.**
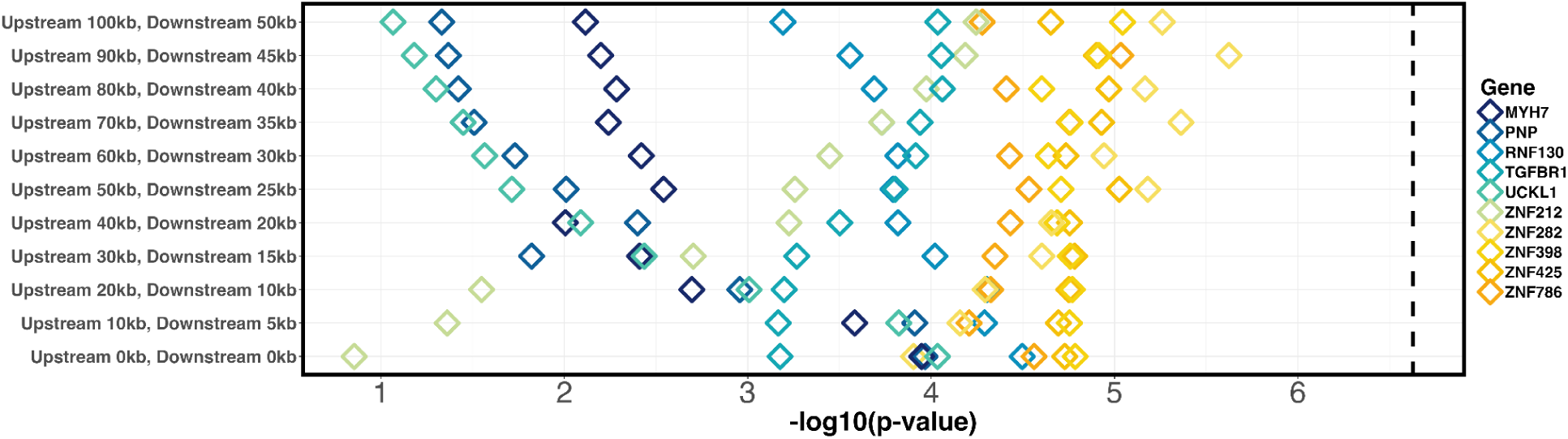
MAGMA gene tests for AML. Using genome-wide association summary statistics for AML, adjusted for the standard set of covariates, MAGMA analysis was conducted to evaluate window-size effects across 11 iterations. The analysis began with 0 kb upstream and 0 kb downstream windows. At each subsequent iteration, the upstream window size was increased by 10 kb and the downstream window size by 5 kb, reaching final windows of 100 kb upstream and 50 kb downstream. The -log10 p-values for the top 10 genes from MAGMA gene-based association tests are displayed for each window configuration. The dashed black line represents the significance threshold after multiple hypothesis correction.

**Supplementary Figure 9.**
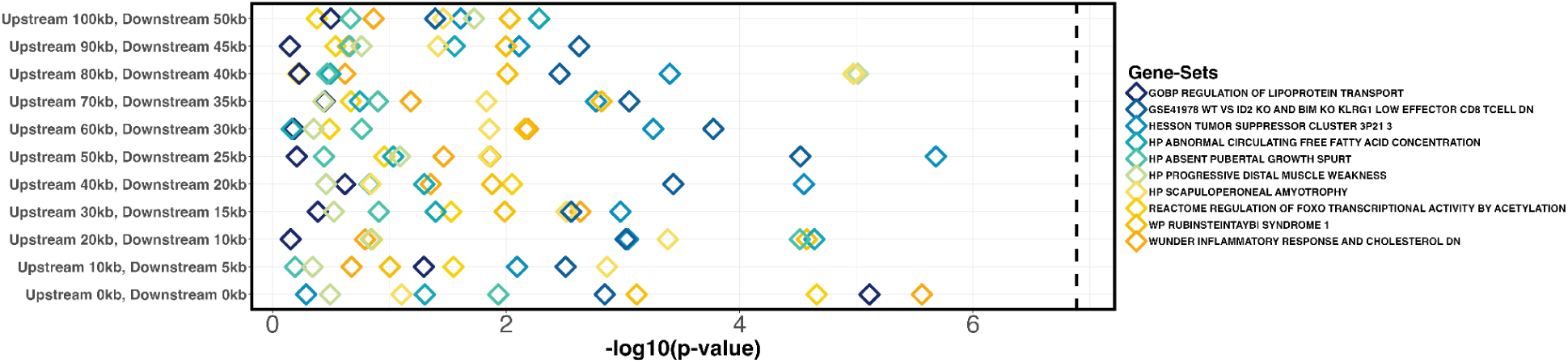
MAGMA gene-set tests for AML. Using genome-wide association summary statistics for AML, adjusted for the standard set of covariates, MAGMA analysis was conducted to evaluate window-size effects across 11 iterations. The analysis began with 0 kb upstream and 0 kb downstream windows. At each subsequent iteration, the upstream window size was increased by 10 kb and the downstream window size by 5 kb, reaching final windows of 100 kb upstream and 50 kb downstream. The -log10 p-values for the top 10 gene-sets from MAGMA gene-set association tests are displayed for each window configuration. The dashed black line represents the significance threshold after multiple hypothesis correction.

**Supplementary Figure 10.**
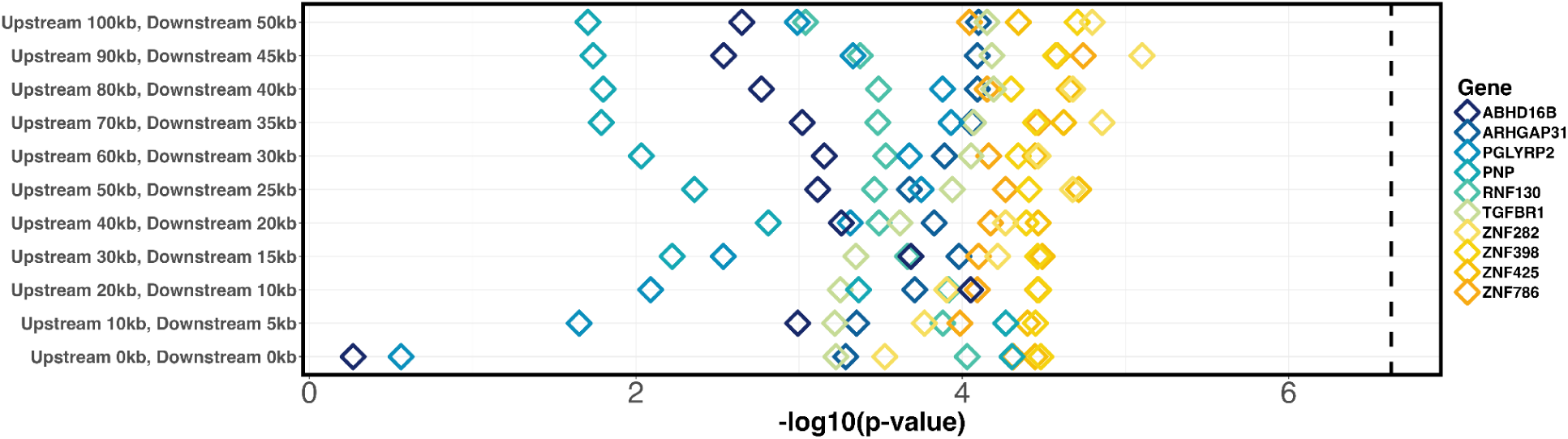
MAGMA gene tests for AML adjusted for the deprivation index. Using genome-wide association summary statistics for AML, adjusted for the standard set of covariates plus the deprivation index, MAGMA analysis was conducted to evaluate window-size effects across 11 iterations. The analysis began with 0 kb upstream and 0 kb downstream windows. At each subsequent iteration, the upstream window size was increased by 10 kb and the downstream window size by 5 kb, reaching final windows of 100 kb upstream and 50 kb downstream. The -log10 p-values for the top 10 genes from MAGMA gene-based association tests are displayed for each window configuration. The dashed black line represents the significance threshold after multiple hypothesis correction.

**Supplementary Figure 11.**
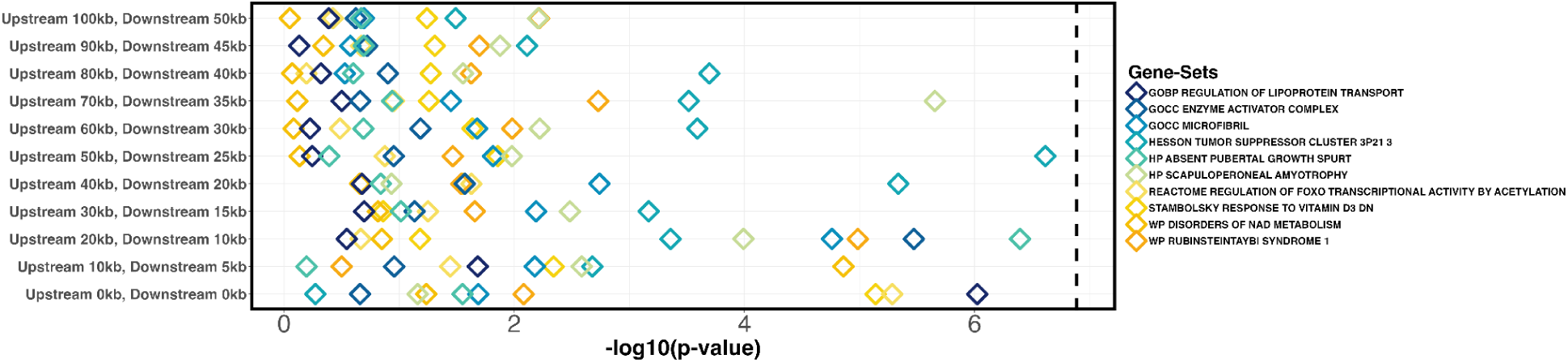
Supplementary Figure. MAGMA gene-set tests for AML adjusted for the deprivation index. Using genome-wide association summary statistics for AML, adjusted for the standard set of covariates plus the deprivation index, MAGMA analysis was conducted to evaluate window-size effects across 11 iterations. The analysis began with 0 kb upstream and 0 kb downstream windows. At each subsequent iteration, the upstream window size was increased by 10 kb and the downstream window size by 5 kb, reaching final windows of 100 kb upstream and 50 kb downstream. The -log10 p-values for the top 10 gene-sets from MAGMA gene-set association tests are displayed for each window configuration. The dashed black line represents the significance threshold after multiple hypothesis correction.

**Supplementary Figure 12.**
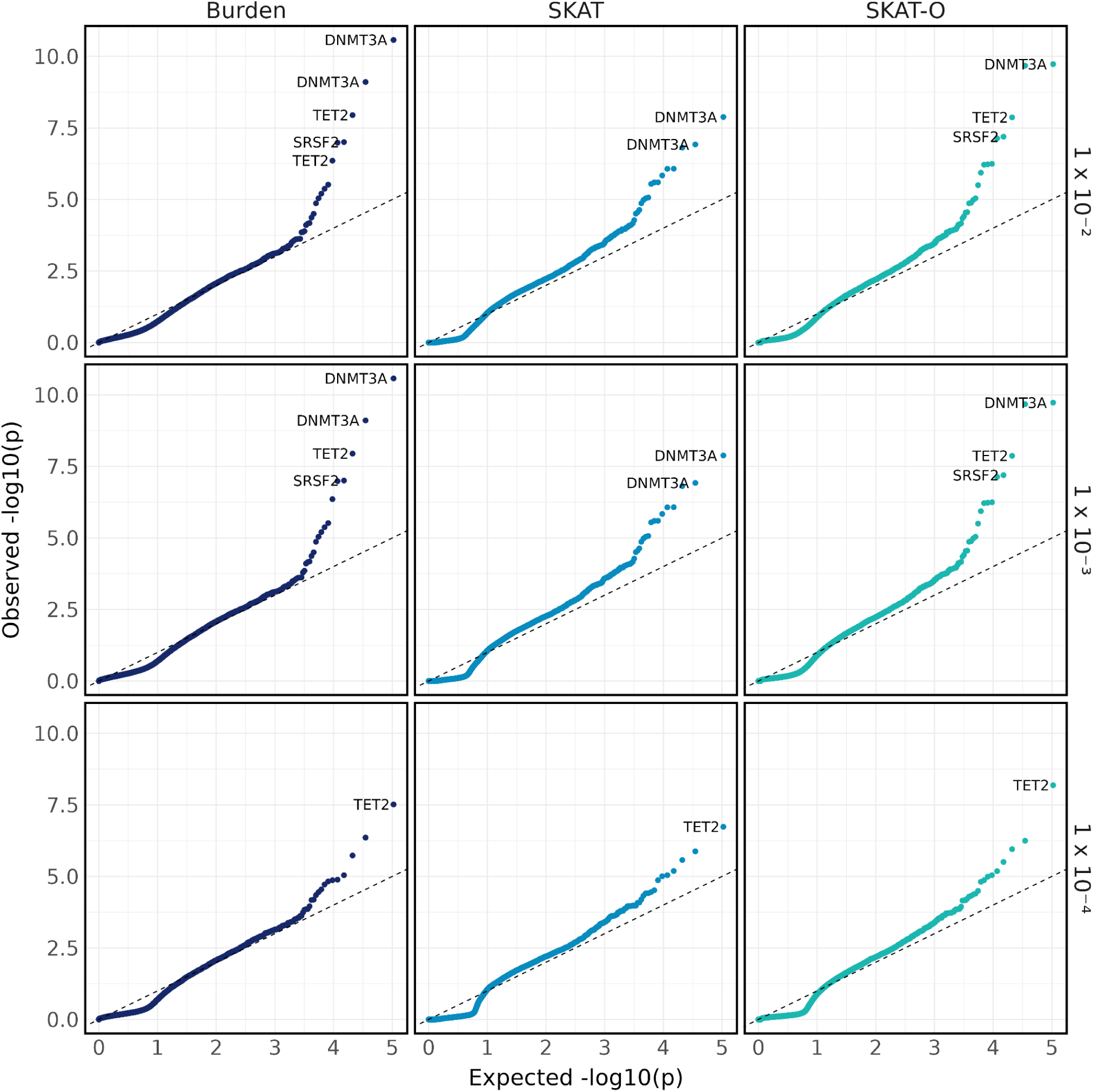
Quantile-quantile plot of AML set-based tests. Quantile-quantile plot of p-values from set-based tests, stratified by test type (Burden, SKAT, and SKAT-O) and maximum MAF thresholds (1%, 0.1%, and 0.01%).

**Supplementary Figure 13.**
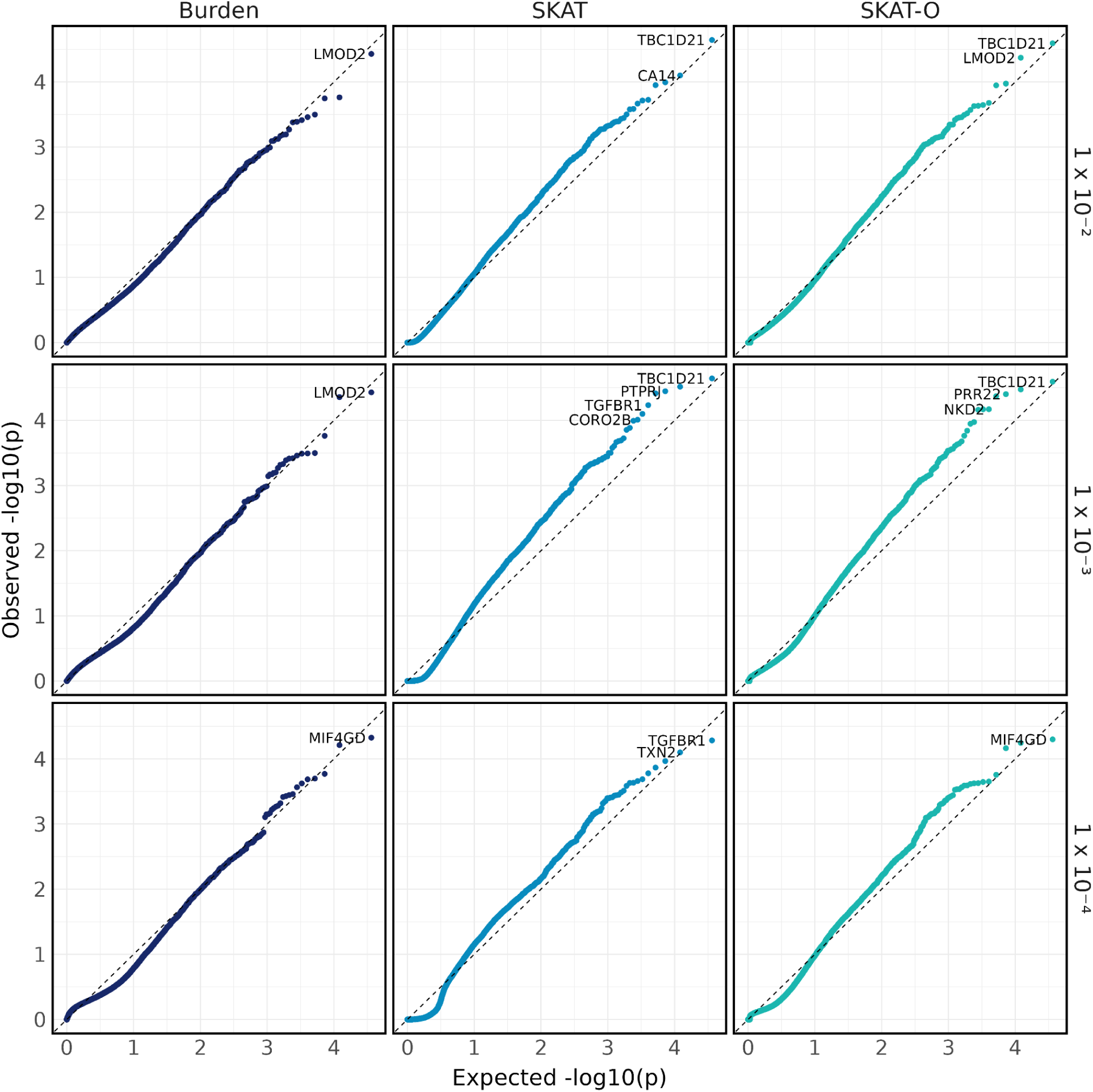
Quantile-quantile plot of AML synonymous set-based tests. Quantile-quantile plot of p-values from synonymous set-based tests, stratified by test type (Burden, SKAT, and SKAT-O) and maximum MAF thresholds (1%, 0.1%, and 0.01%).

**Supplementary Figure 14.**
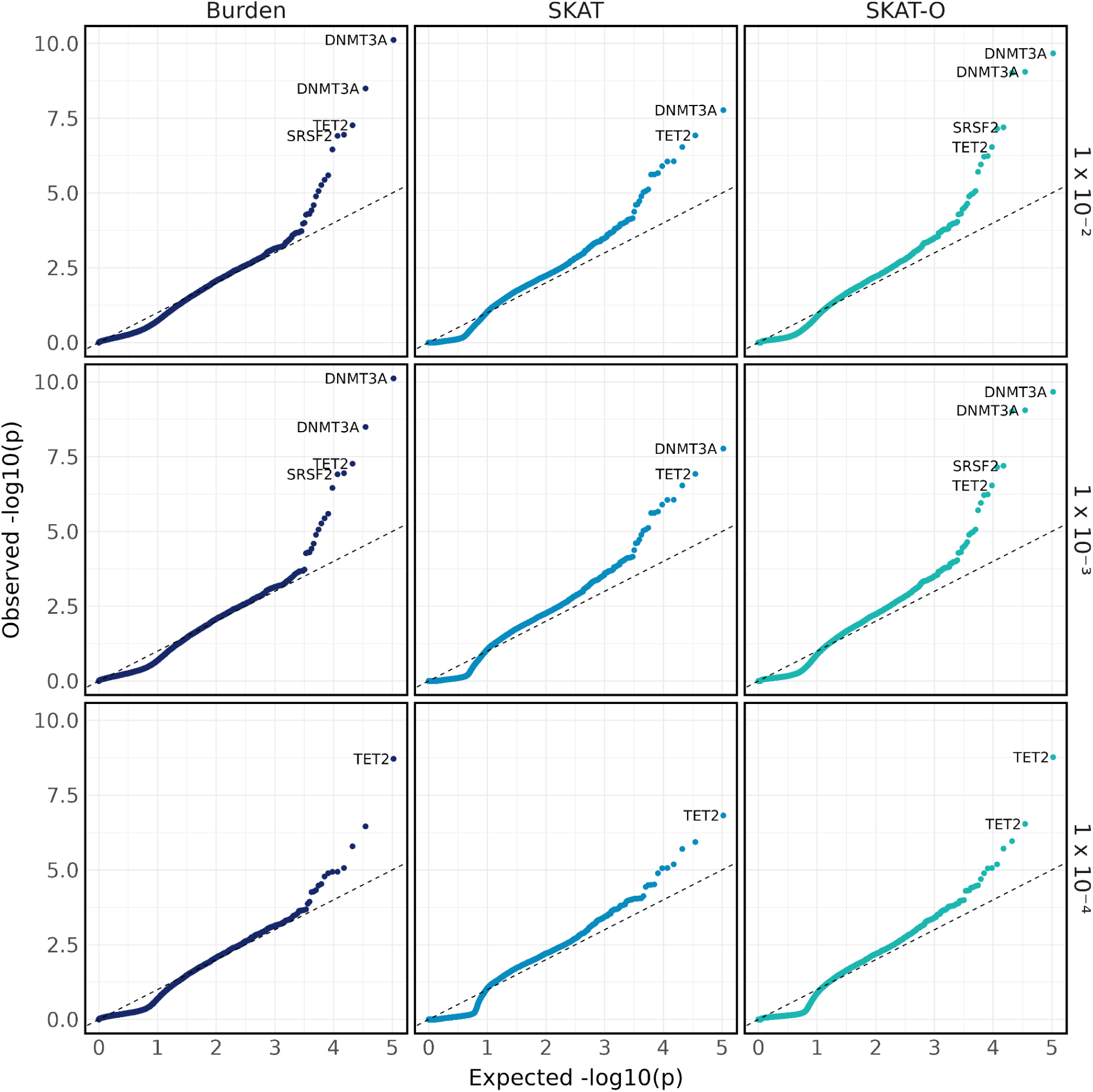
Quantile-quantile plot of AML set-based tests adjusted for the deprivation index. Quantile-quantile plot of p-values from set-based tests adjusting for the deprivation index, stratified by test type (Burden, SKAT, and SKAT-O) and maximum MAF thresholds (1%, 0.1%, and 0.01%).

**Supplementary Figure 15.**
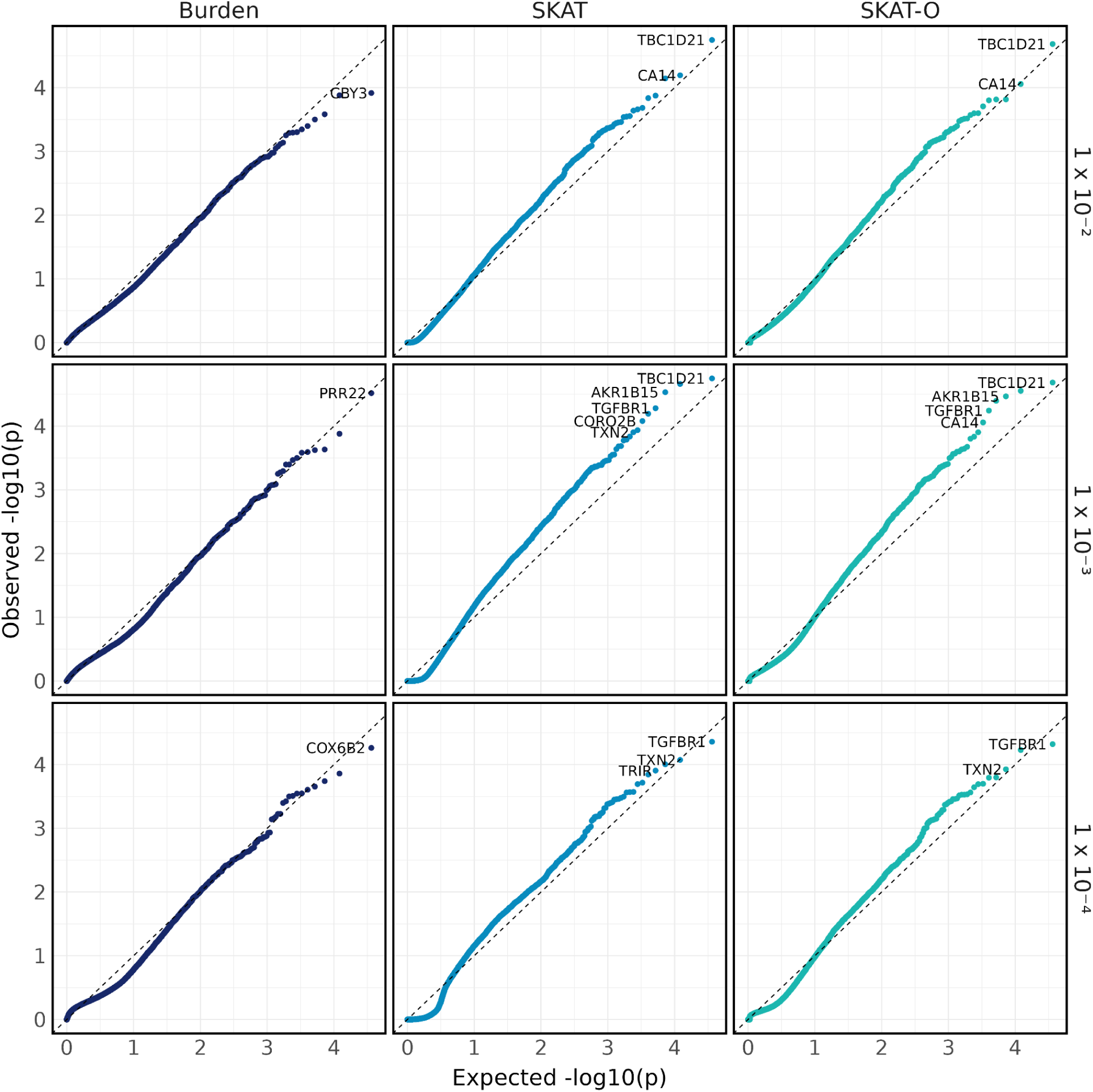
Quantile-quantile plot of AML synonymous set-based tests adjusted for the deprivation index. Quantile-quantile plot of p-values from synonymous set-based tests adjusting for the deprivation index, stratified by test type (Burden, SKAT, and SKAT-O) and maximum MAF thresholds (1%, 0.1%, and 0.01%).

**Supplementary Figure 16.**
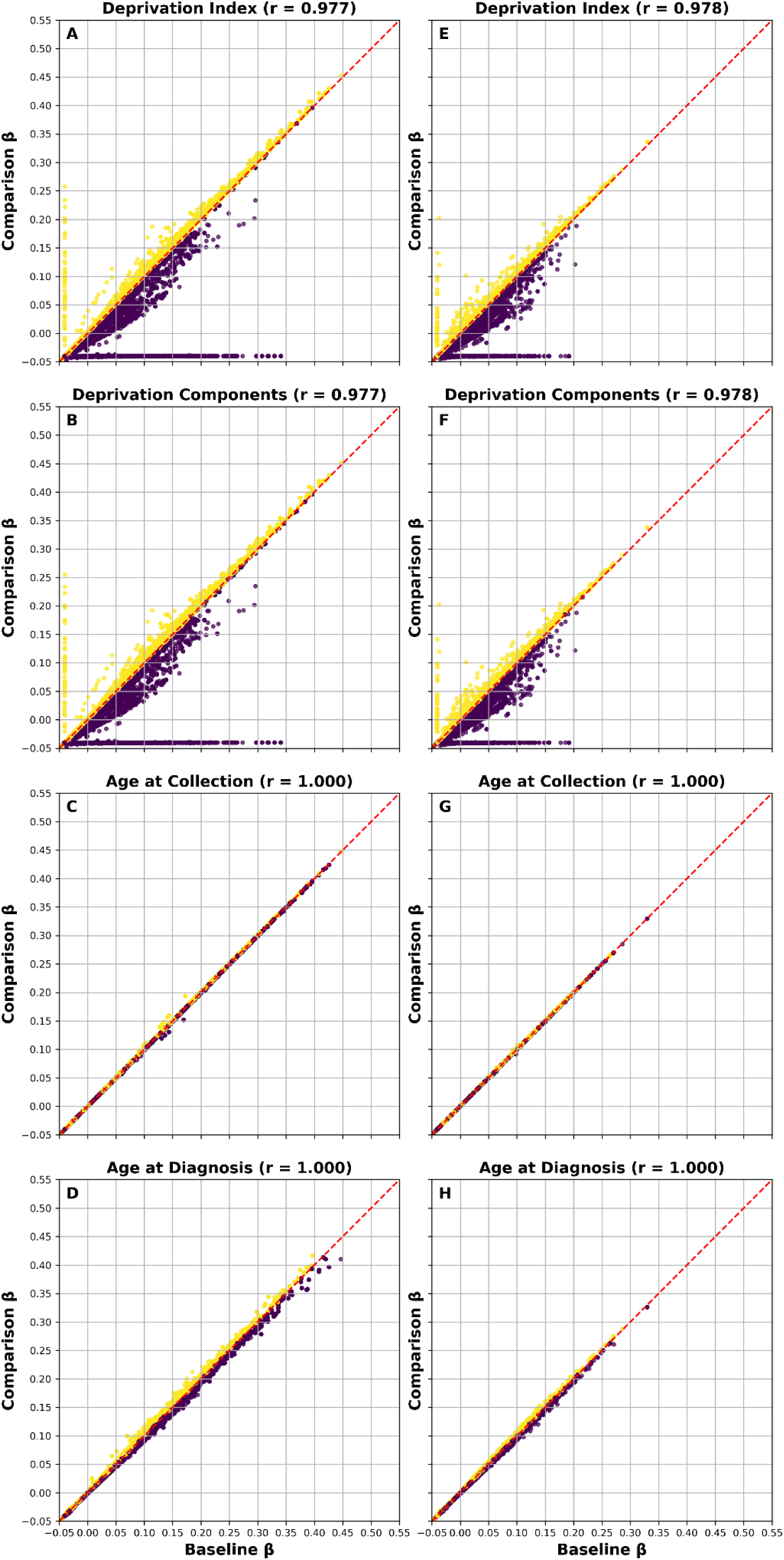
Correlations between burden coefficients from set-based tests. Baseline gene set-based tests, which used the standard set of covariates (age, sex, and principal components), were compared against models in which the covariates were modified. The “Deprivation Index” model (A & E) includes the deprivation index as an additional covariate, whereas the “Deprivation Components” model (B & F) includes the individual components of the deprivation index as covariates. The “Age at Collection” model (C & G) defines age based on age at biosample collection for whole-genome sequencing, while the “Age at Diagnosis” model (D & H) defines age using age at diagnosis for cases and age at the last EHR record for controls. The plots on the left (A-D) represent tests using the variant set predicted to be deleterious (loss-of-function or missense), while plots on the right (E-H) represent tests employing the synonymous variant set. The Pearson correlation coefficient was calculated and is reported at the top of each plot.

**Supplementary Figure 17.**
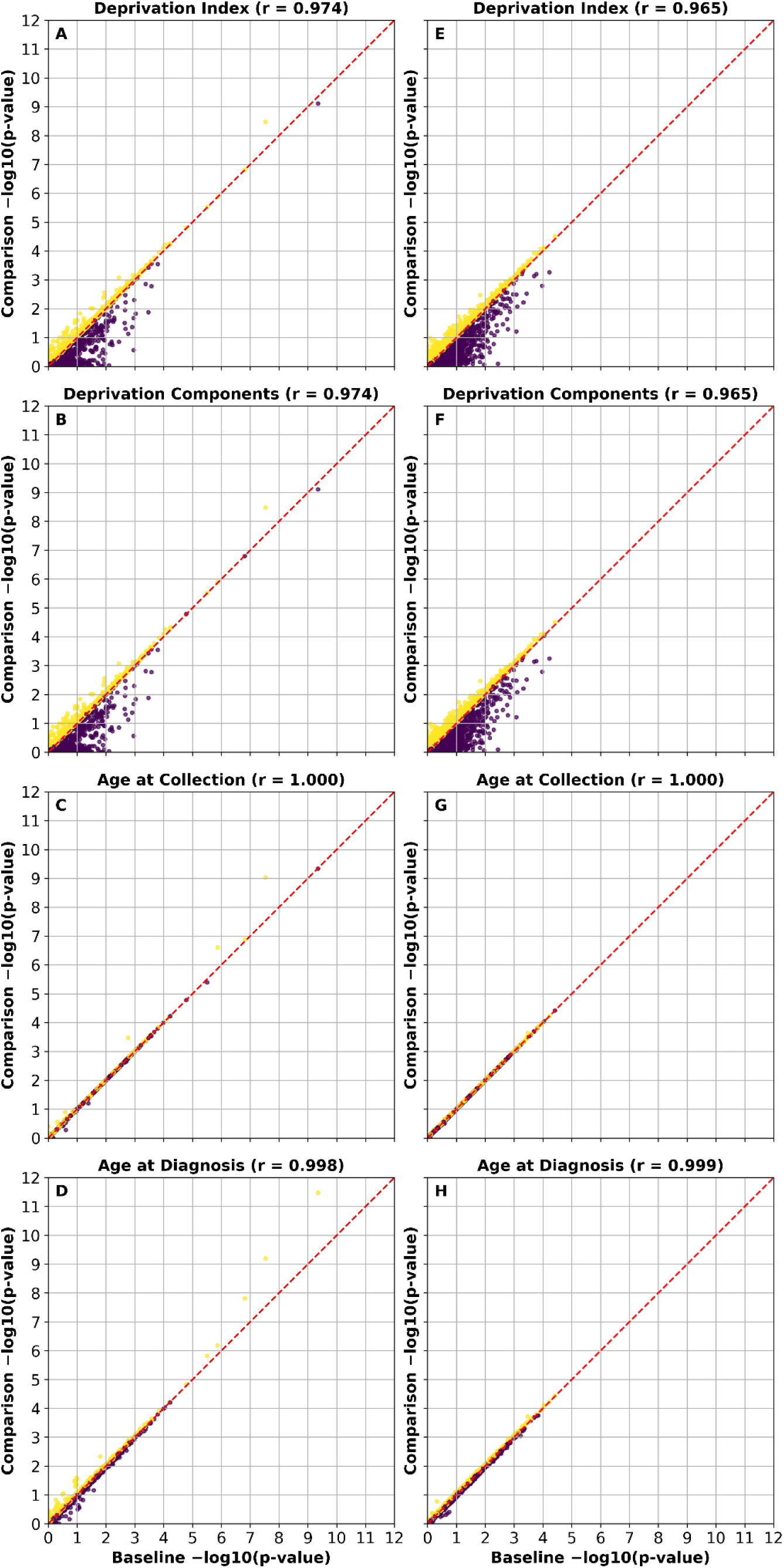
Correlations between Cauchy SKAT-O p-values from set-based tests. Baseline gene set-based tests, which used the standard set of covariates (age, sex, and principal components), were compared against models in which the covariates were modified. The “Deprivation Index” model (A & E) includes the deprivation index as an additional covariate, whereas the “Deprivation Components” model (B & F) includes the individual components of the deprivation index as covariates. The “Age at Collection” model (C & G) defines age based on age at biosample collection for whole-genome sequencing, while the “Age at Diagnosis” model (D & H) defines age using age at diagnosis for cases and age at the last EHR record for controls. The plots on the left (A-D) represent tests using the variant set predicted to be deleterious (loss-of-function or missense), while plots on the right (E-H) represent tests employing the synonymous variant set. The Pearson correlation coefficient was calculated and is reported at the top of each plot.

**Supplementary Figure 18.**
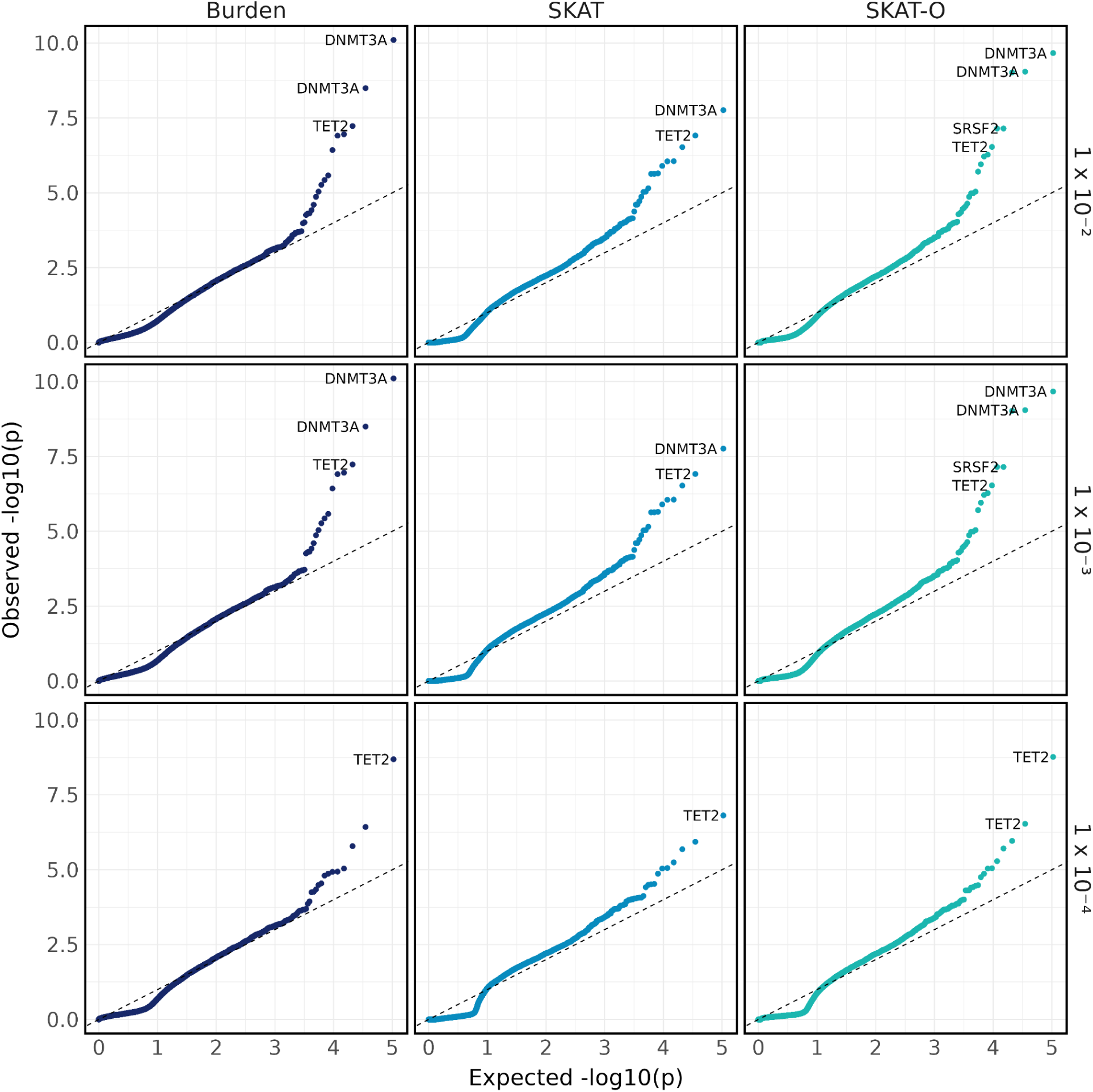
Quantile-quantile plot of AML set-based tests adjusted for the deprivation components. Quantile-quantile plot of p-values from set-based tests adjusting for the components of the deprivation index, stratified by test type (Burden, SKAT, and SKAT-O) and maximum MAF thresholds (1%, 0.1%, and 0.01%).

**Supplementary Figure 19.**
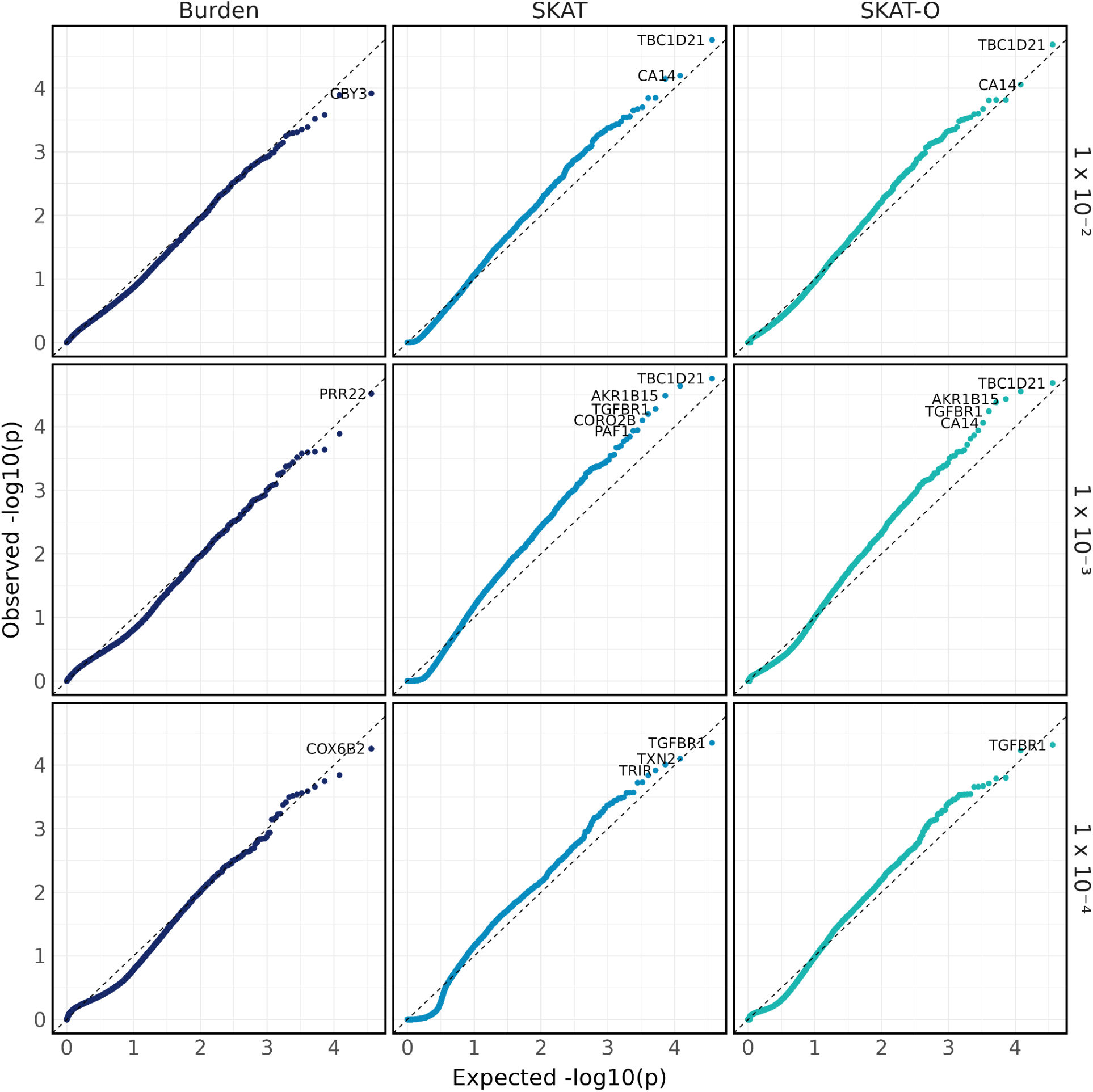
Quantile-quantile plot of AML synonymous set-based tests adjusted for the deprivation components. Quantile-quantile plot of p-values from synonymous set-based tests adjusting for the components of the deprivation index, stratified by test type (Burden, SKAT, and SKAT-O) and maximum MAF thresholds (1%, 0.1%, and 0.01%).

**Supplementary Figure 20.**
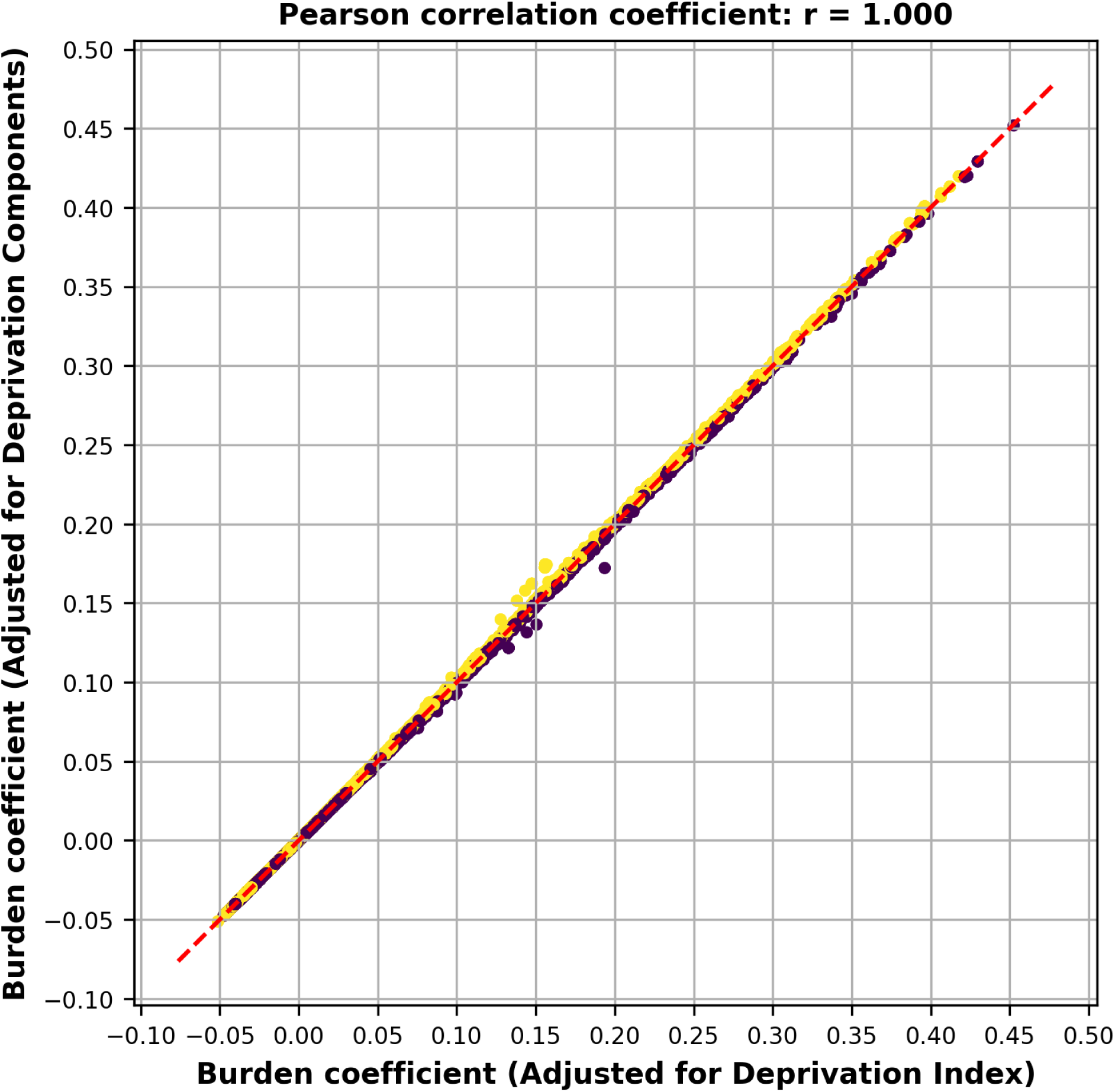
Burden coefficients from set-based tests adjusted for the deprivation index versus the deprivation components. Correlations between Burden coefficients p-values from the set-based tests are shown, with models adjusted for the deprivation index and for the individual components of the deprivation index as covariates. The Pearson correlation coefficient was calculated and is reported at the top of each plot.

**Supplementary Figure 21.**
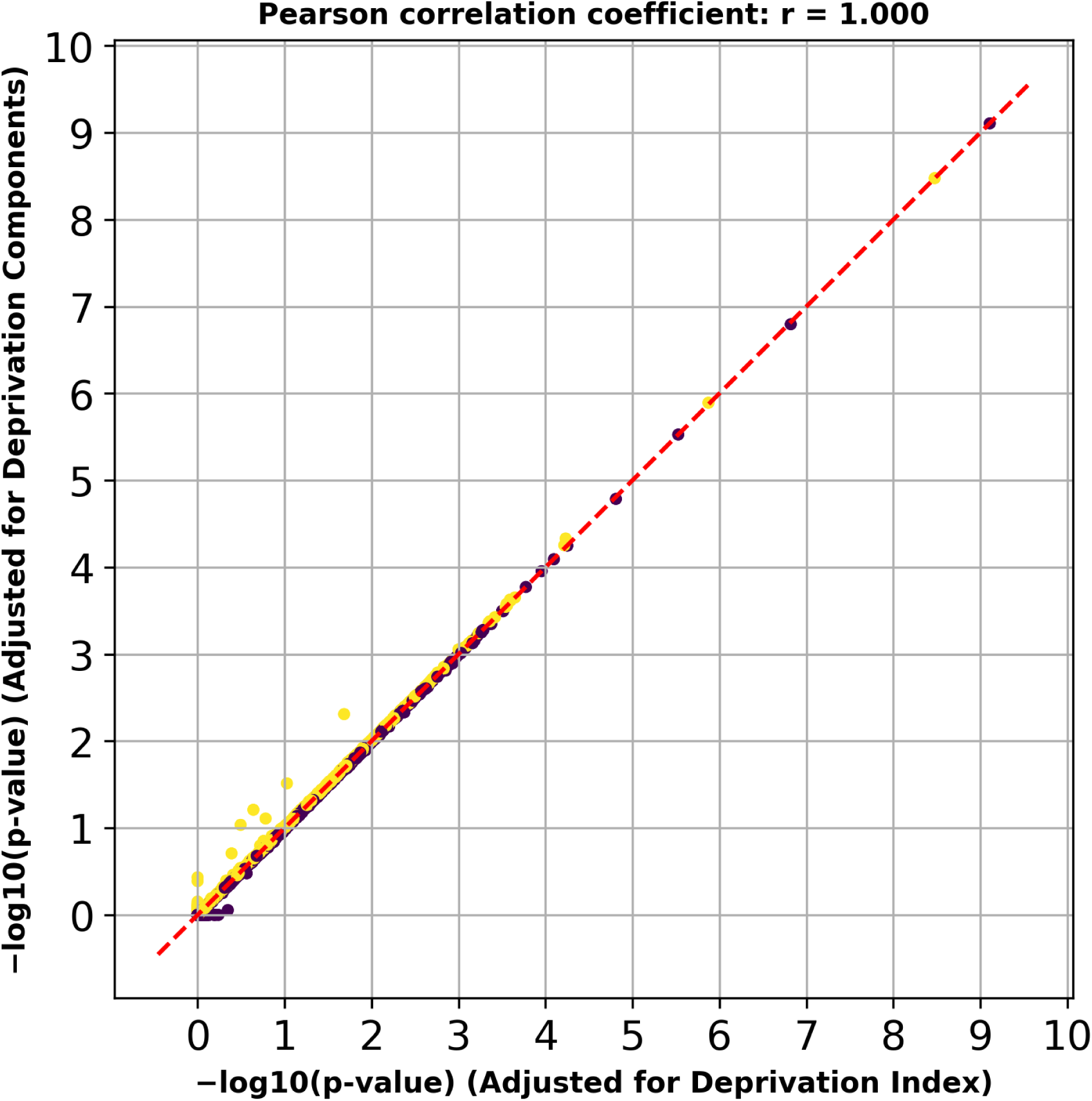
Cauchy SKAT-O p-values from set-based tests adjusted for the deprivation index versus the deprivation components. **Correlations between Cauchy** SKAT-O p-values from the set-based tests are shown, with models adjusted for the deprivation index and for the individual components of the deprivation index as covariates. The Pearson correlation coefficient was calculated and is reported at the top of each plot.

**Supplementary Figure 22.**
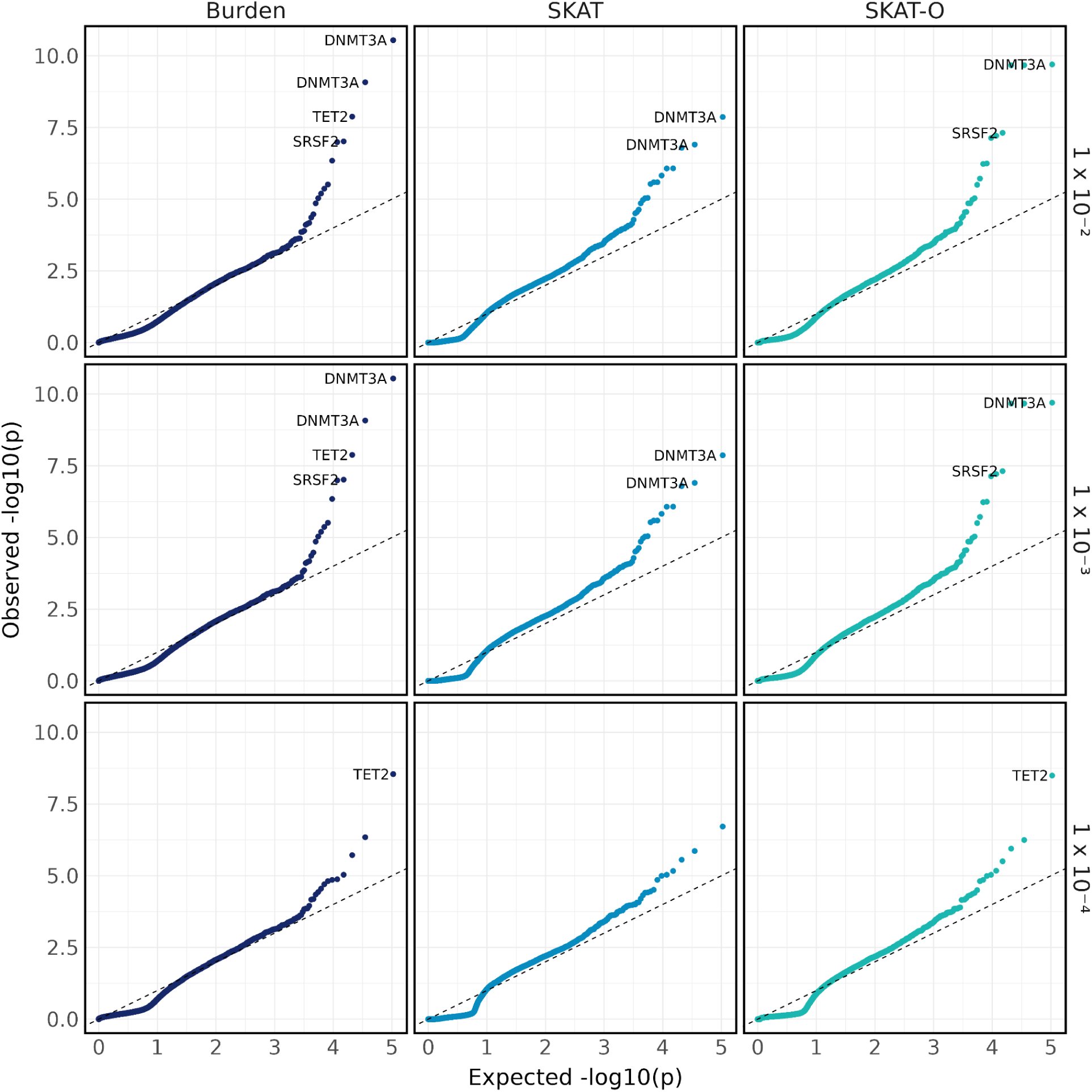
Quantile-quantile plot of AML set-based tests adjusted for the age at collection. Quantile-quantile plot of p-values from set-based tests adjusting for the age of biosample collection, stratified by test type (Burden, SKAT, and SKAT-O) and maximum MAF thresholds (1%, 0.1%, and 0.01%).

**Supplementary Figure 23.**
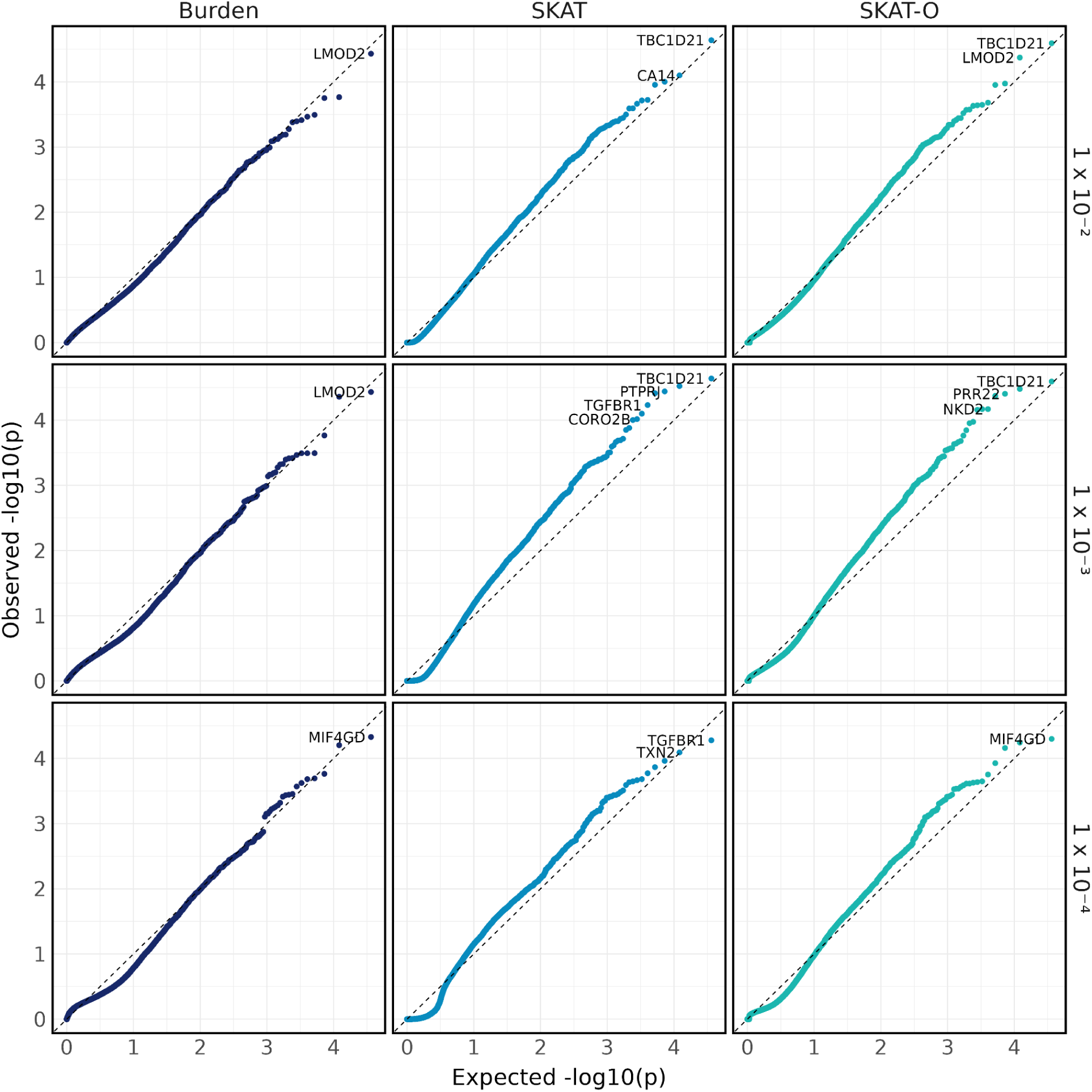
Quantile-quantile plot of AML synonymous set-based tests adjusted for the age at collection. Quantile-quantile plot of p-values from synonymous set-based tests adjusting for the age of biosample collection, stratified by test type (Burden, SKAT, and SKAT-O) and maximum MAF thresholds (1%, 0.1%, and 0.01%).

**Supplementary Figure 24.**
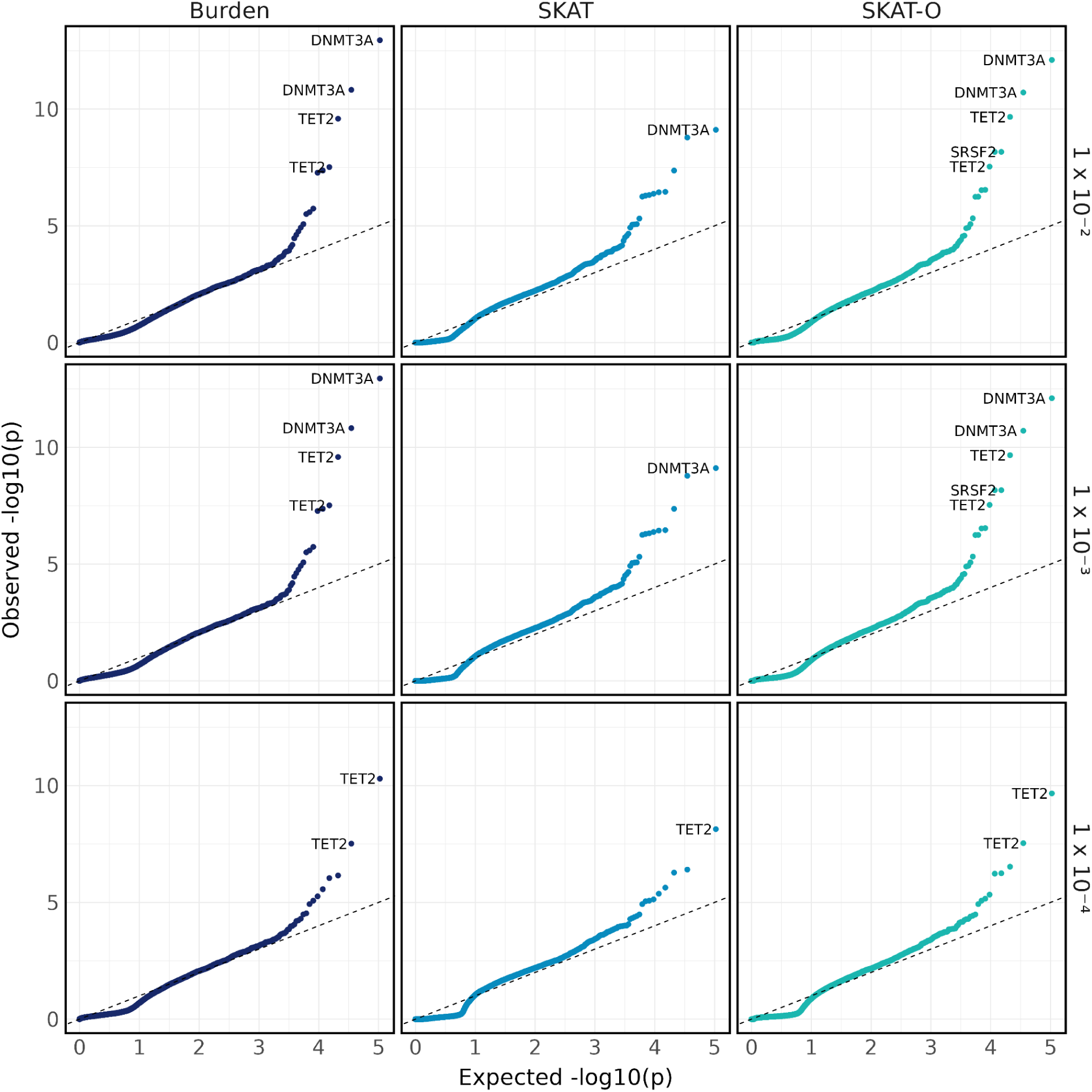
Quantile-quantile plot of AML set-based tests adjusted for the age at diagnosis. Quantile-quantile plot of p-values from set-based tests adjusting for the age at diagnosis, stratified by test type (Burden, SKAT, and SKAT-O) and maximum MAF thresholds (1%, 0.1%, and 0.01%).

**Supplementary Figure 25.**
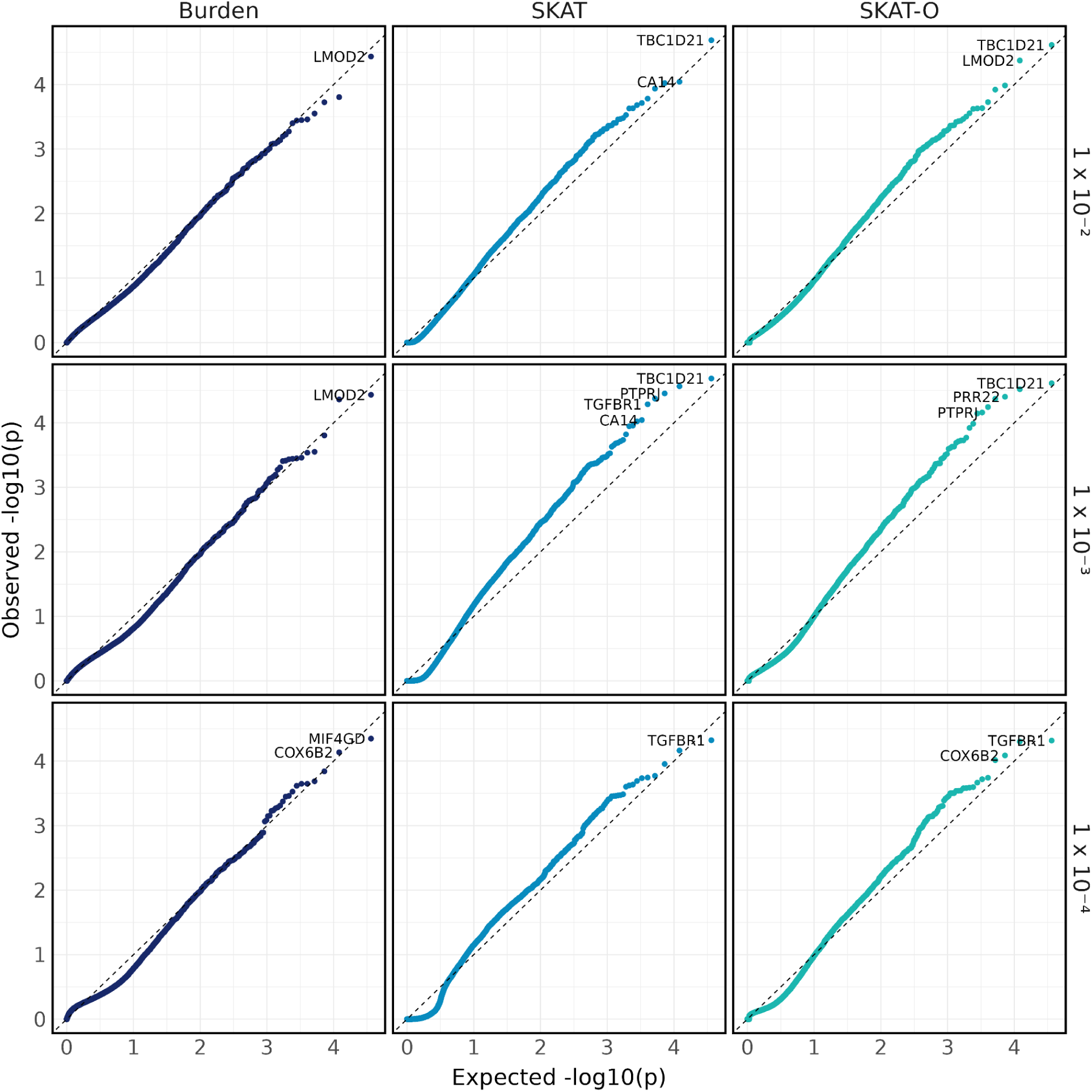
Quantile-quantile plot of AML synonymous set-based tests adjusted for the age at diagnosis. Quantile-quantile plot of p-values from synonymous set-based tests adjusting for the age at diagnosis, stratified by test type (Burden, SKAT, and SKAT-O) and maximum MAF thresholds (1%, 0.1%, and 0.01%).

**Supplementary Figure 26.**
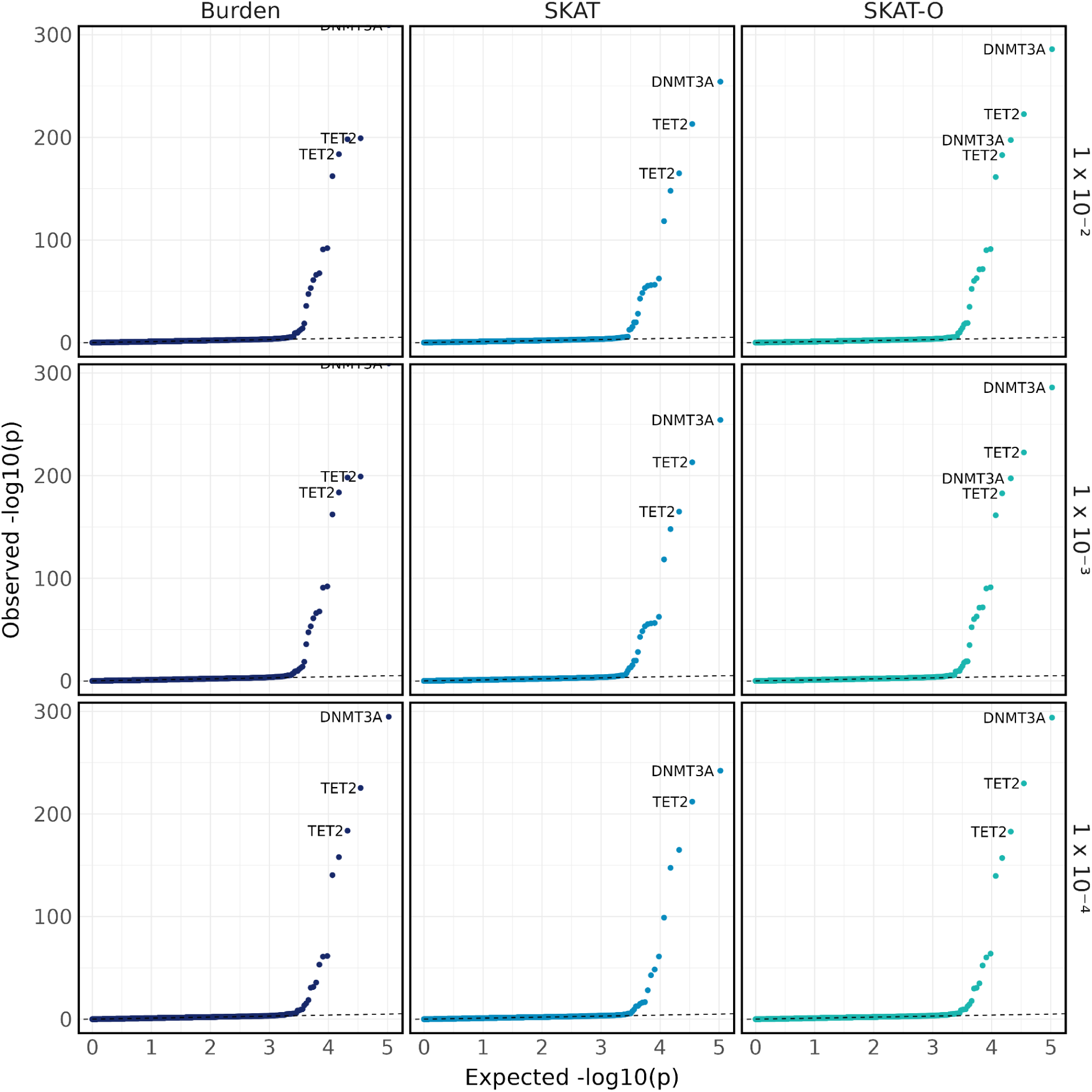
Quantile-quantile plot of age at biospecimen collection set-based tests. Quantile-quantile plot of p-values from set-based tests, stratified by test type (Burden, SKAT, and SKAT-O) and maximum MAF thresholds (1%, 0.1%, and 0.01%).

**Supplementary Figure 27.**
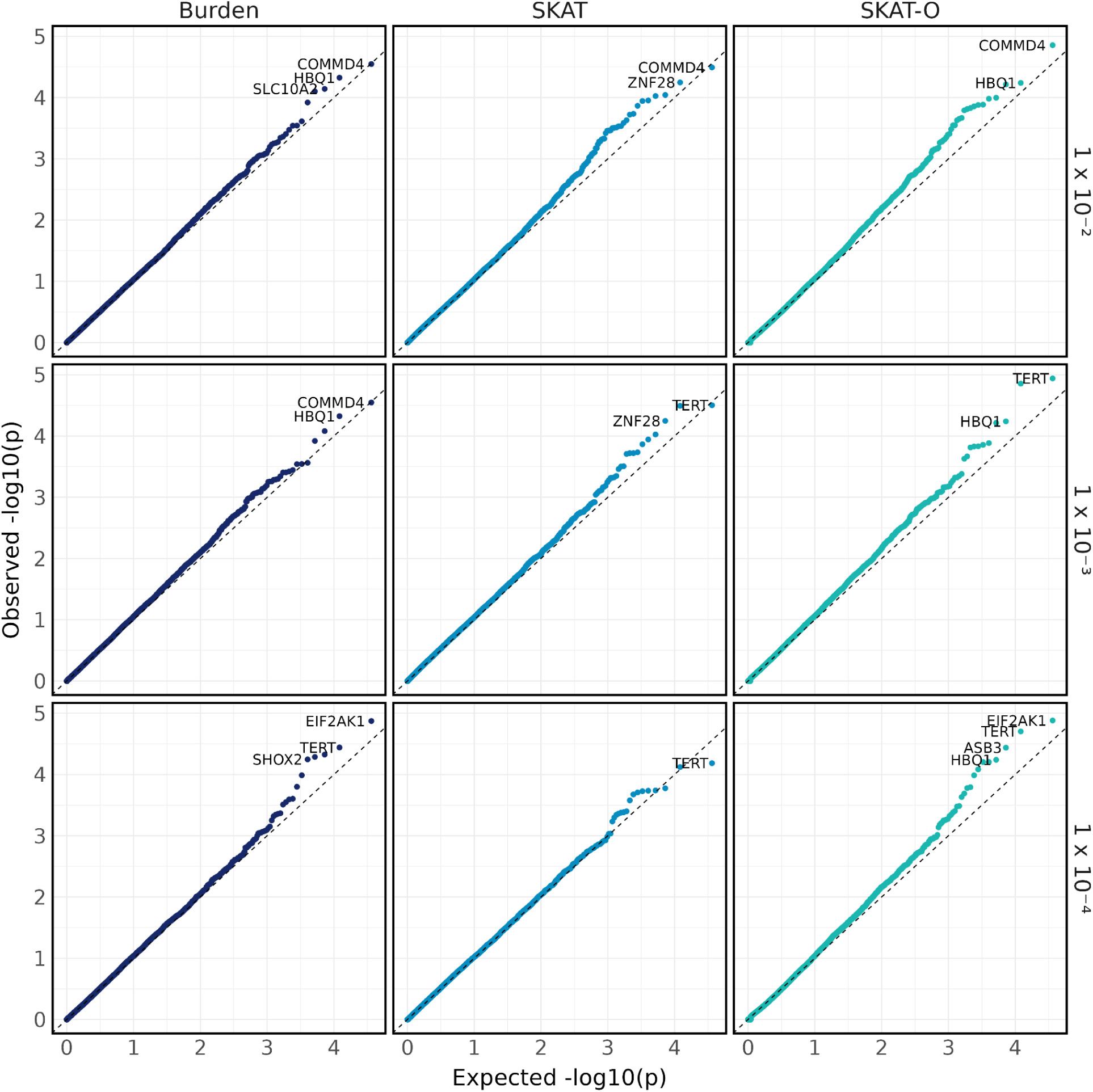
Quantile-quantile plot of age at biospecimen collection synonymous set-based tests. Quantile-quantile plot of p-values from synonymous set-based tests, stratified by test type (Burden, SKAT, and SKAT-O) and maximum MAF thresholds (1%, 0.1%, and 0.01%).

**Supplementary Figure 28.**
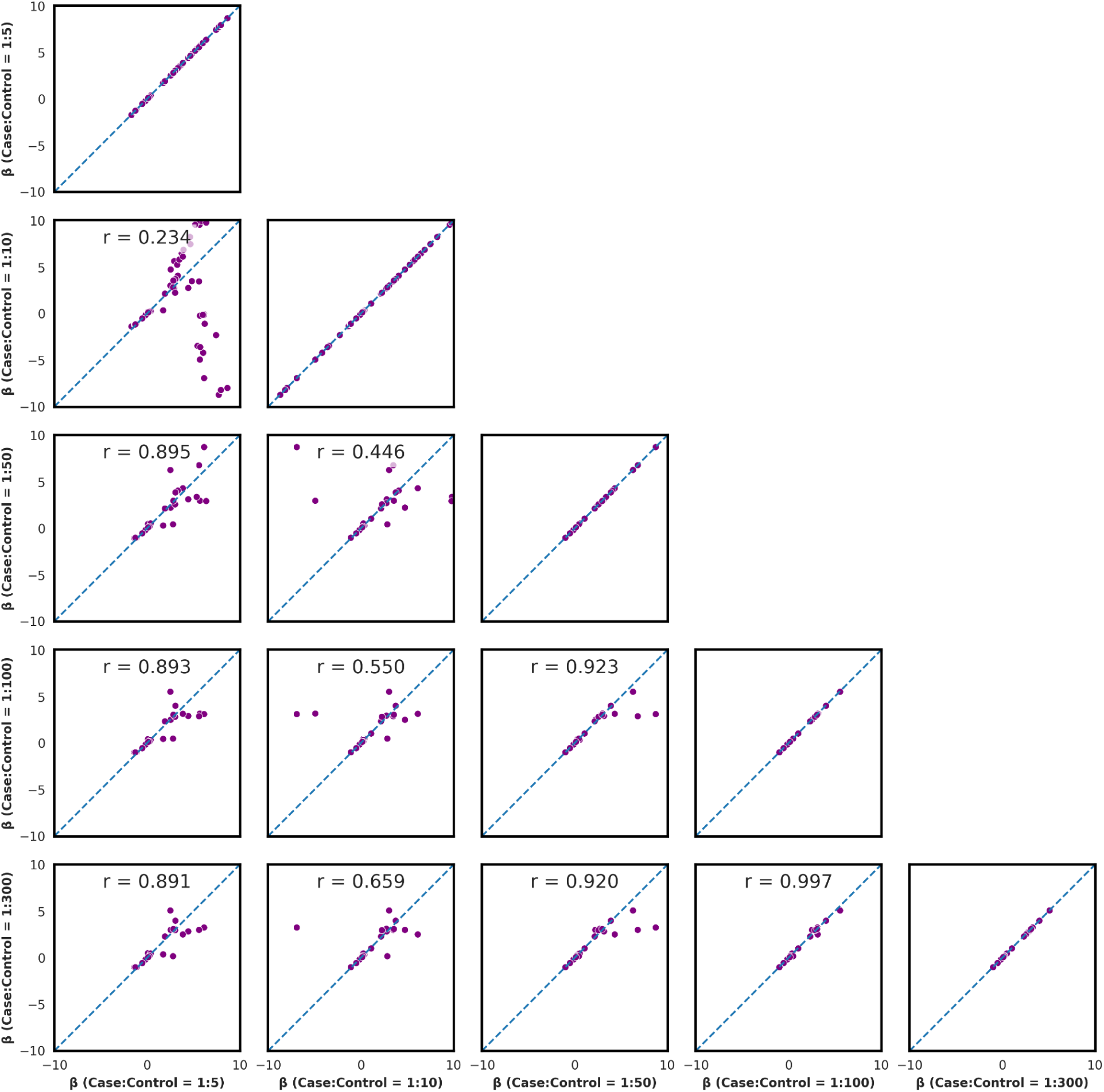
Rare single-variant coefficients in AML top genes in biospecimen-age matched cohorts. Single-variant tests were performed within genes that were significant in the AML set-based tests (*DNMT3A*, *TET2*, *SRSF2*, and *IDH2*), using AML as the phenotype and conducted within each age-at-biospecimen-collection matched cohort. Correlations of effect sizes for rare variants (0.0001 < MAF < 0.01) were calculated to ensure that the matching procedure did not alter effect size estimates across different matching ratios. The Pearson correlation coefficient is reported above each pairwise comparison.

**Supplementary Figure 29.**
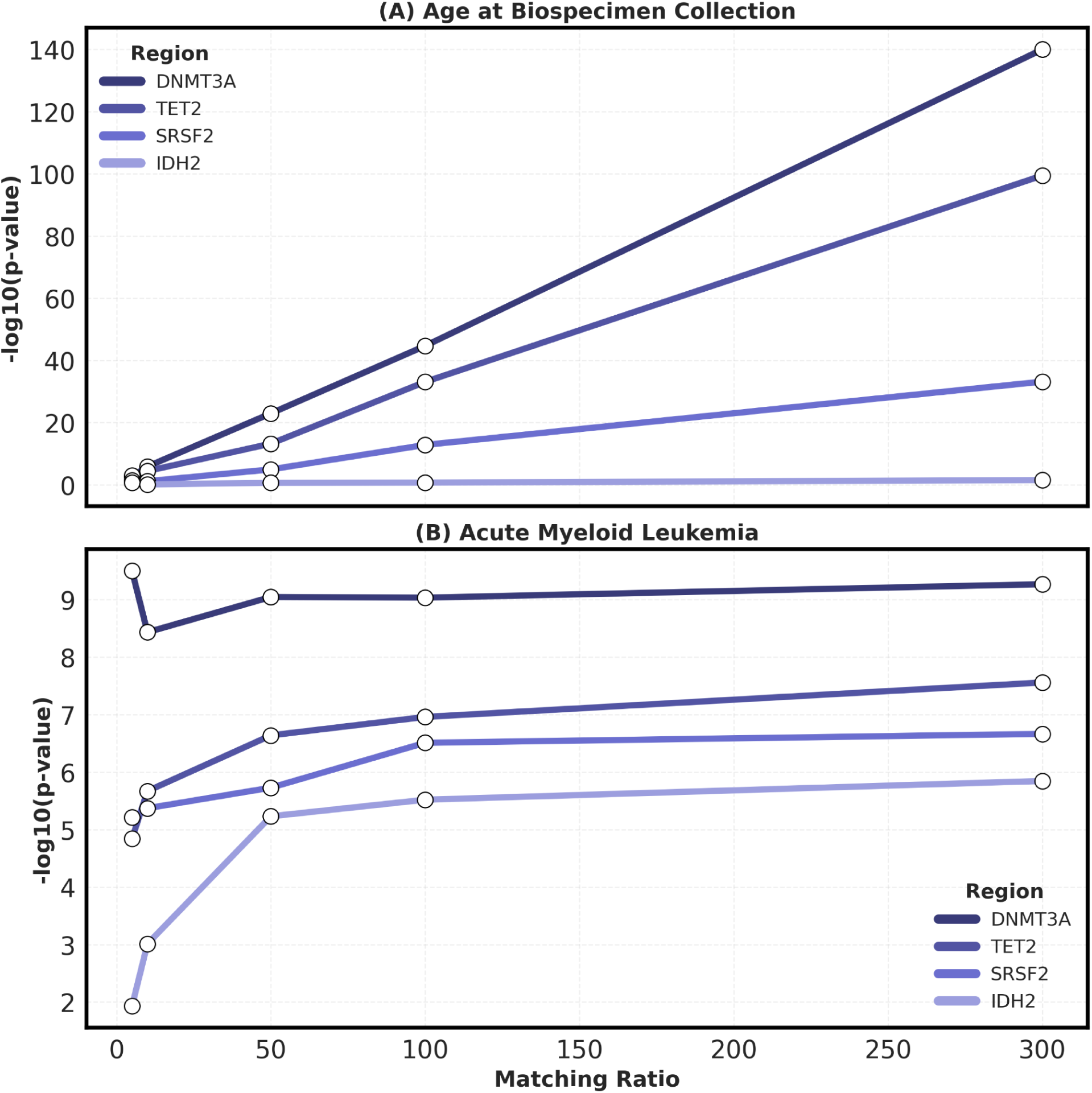
Rare set-based tests in AML top genes in biospecimen-age matched cohorts. The −log10 p-values from the set-based tests using the age-at-biospecimen-collection matched cohorts are displayed. (A) The −log10 p-values from set-based tests in top AML genes (*DNMT3A*, *TET2*, *SRSF2*, and *IDH2*) using age-at-biospecimen-collection as the phenotype and employing different matched cohort ratios. (B) The −log10 p-values from set-based tests in top AML genes (*DNMT3A*, *TET2*, *SRSF2*, and *IDH2*) using AML as the phenotype and employing different matched cohort ratios. The y-axis shows −log10 p-values, and the x-axis shows the case-control matching ratios used for the age-at-biospecimen-collection matched cohorts.

**Supplementary Figure 30.**
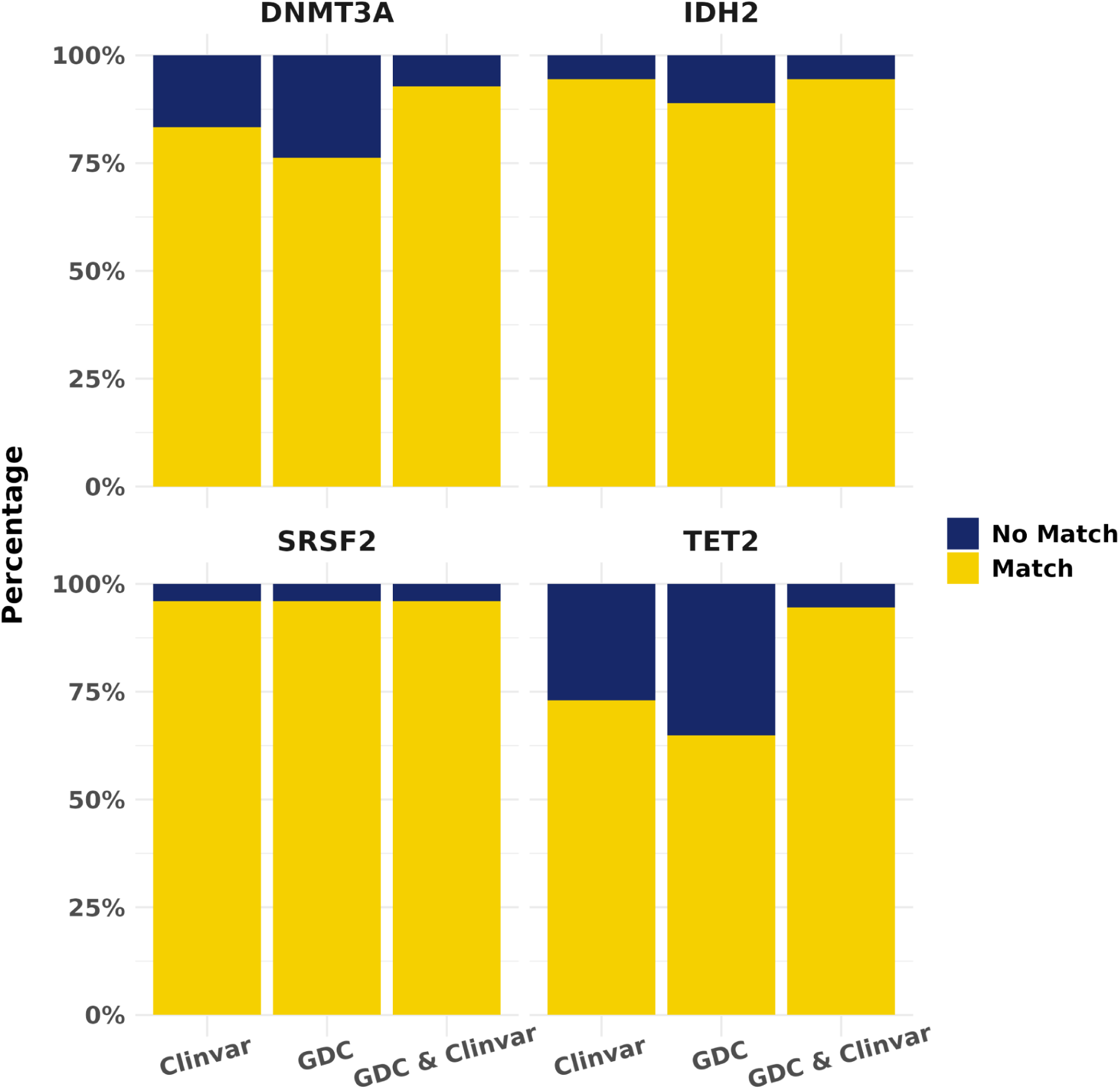
Cross-reference rare variants with the GDC and ClinVar. For the genes identified from the gene set-based tests, rare variants (MAF < 1% and MAC ≥ 10) from the variant sets were cross-referenced with variants from the GDC and ClinVar databases. The proportion of overlapping rare variants is displayed as a percentage.

**Supplementary Figure 31.**
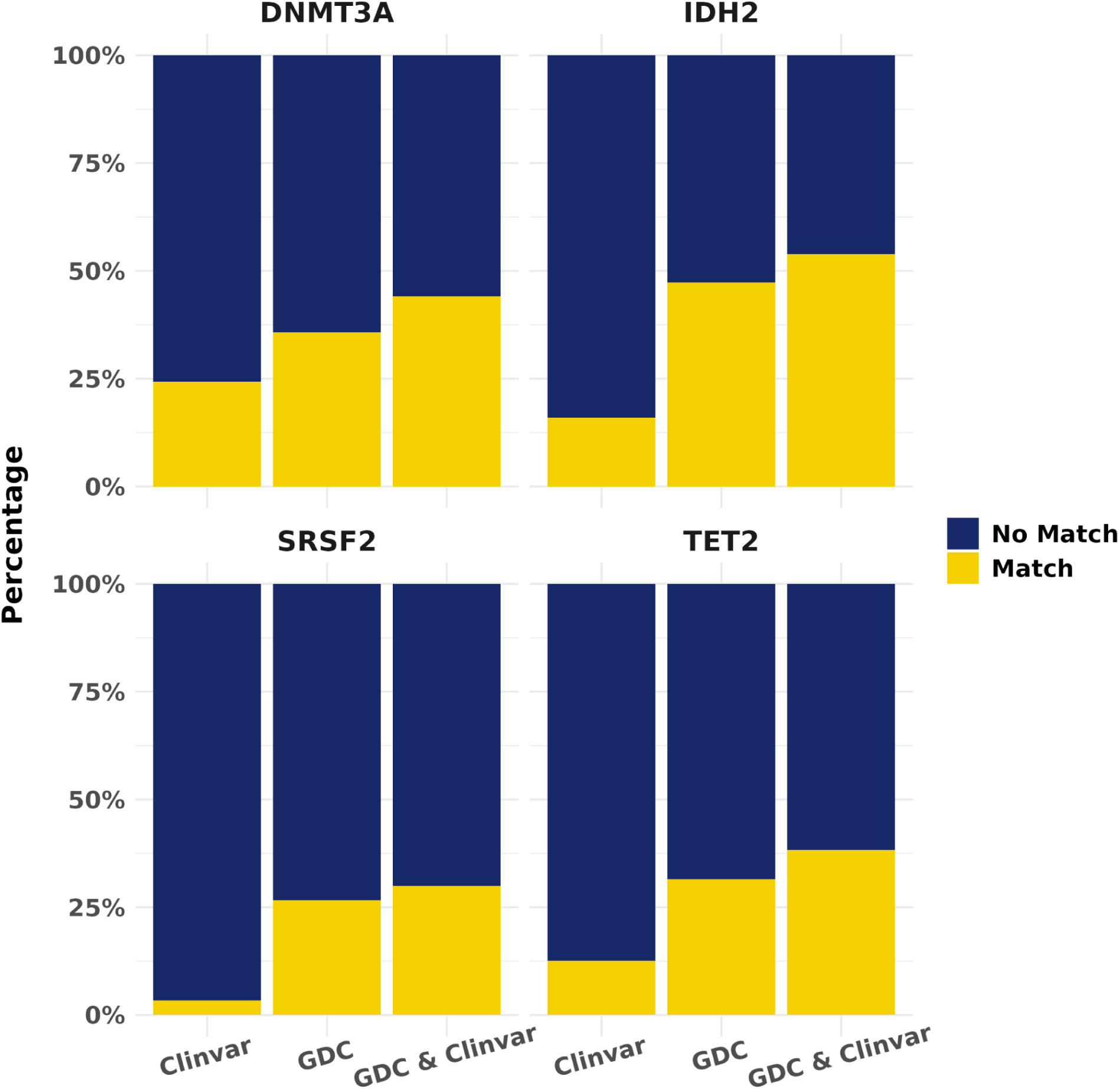
Cross-reference ultra-rare variants with the GDC and ClinVar. For the genes identified from the gene set-based tests, ultra-rare variants (MAC < 10) from the variant sets were cross-referenced with variants from the GDC and ClinVar databases. The proportion of overlapping ultra-rare variants is displayed as a percentage.

**Supplementary Figure 32.**
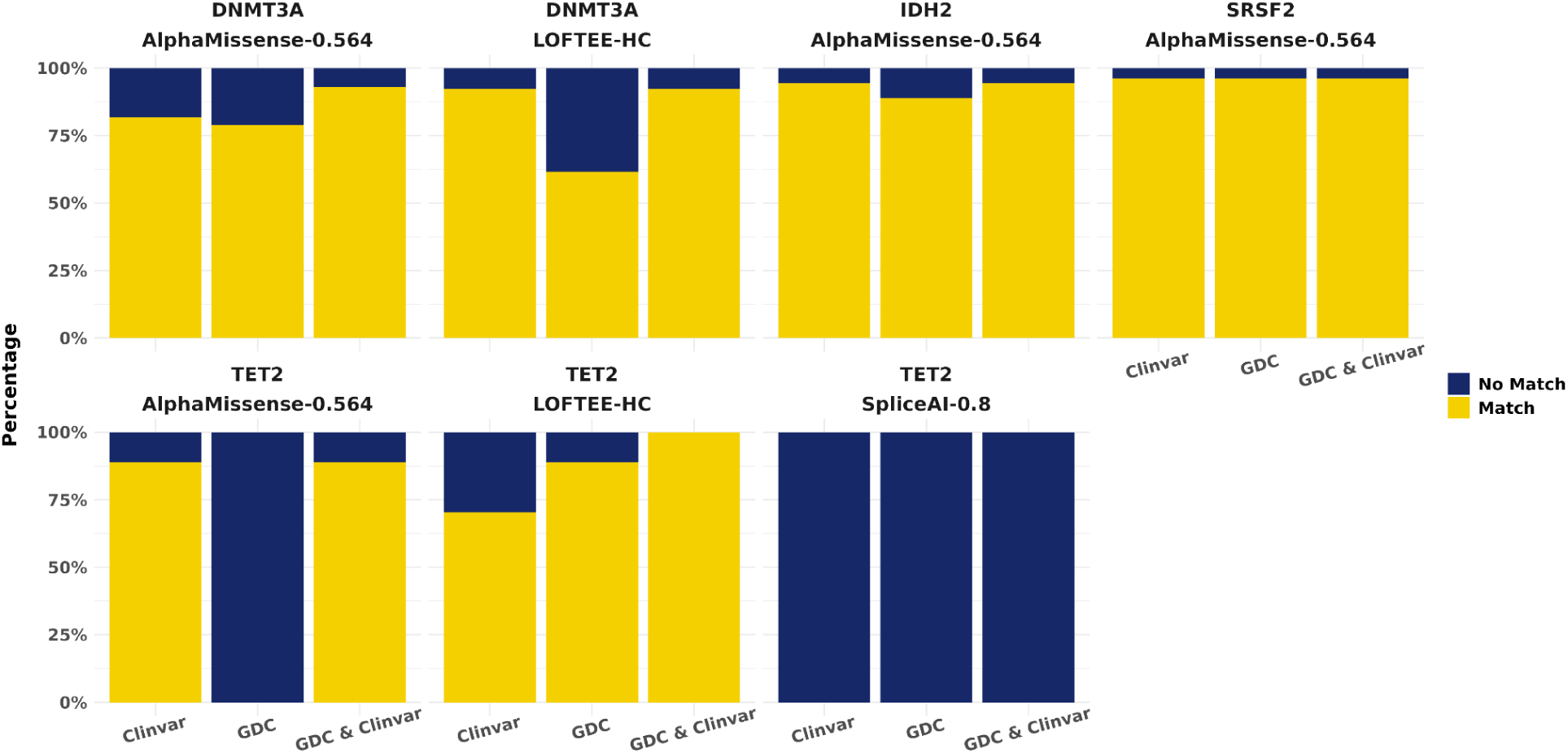
Cross-reference rare variants with the GDC and ClinVar, stratified by annotation tool. For the genes identified from the gene set-based tests, rare variants (MAF < 1% and MAC ≥ 10) from the variant sets were cross-referenced with variants from the GDC and ClinVar databases. The proportion of overlapping rare variants is displayed as a percentage, stratified by the annotation tools used to designate variant function (LOFTEE, SpliceAI, or AlphaMissense).

**Supplementary Figure 33.**
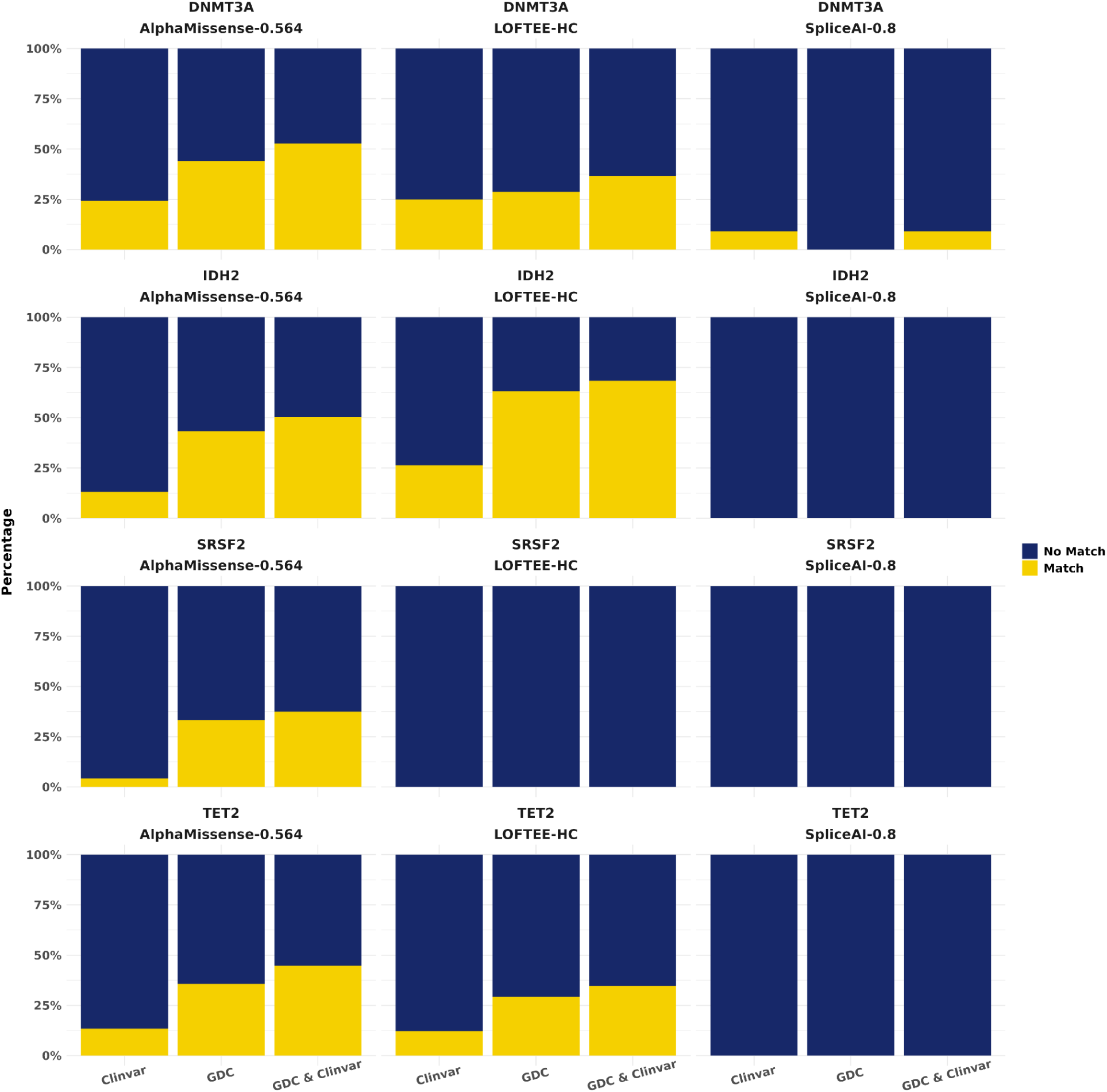
Cross-reference ultra-rare variants with the GDC and ClinVar, stratified by annotation tool. For the genes identified from the gene set-based tests, ultra-rare variants (MAC < 10) from the variant sets were cross-referenced with variants from the GDC and ClinVar databases. The proportion of overlapping ultra-rare variants is displayed as a percentage, stratified by the annotation tools used to designate variant function (LOFTEE, SpliceAI, or AlphaMissense).

**Supplementary Figure 34.**
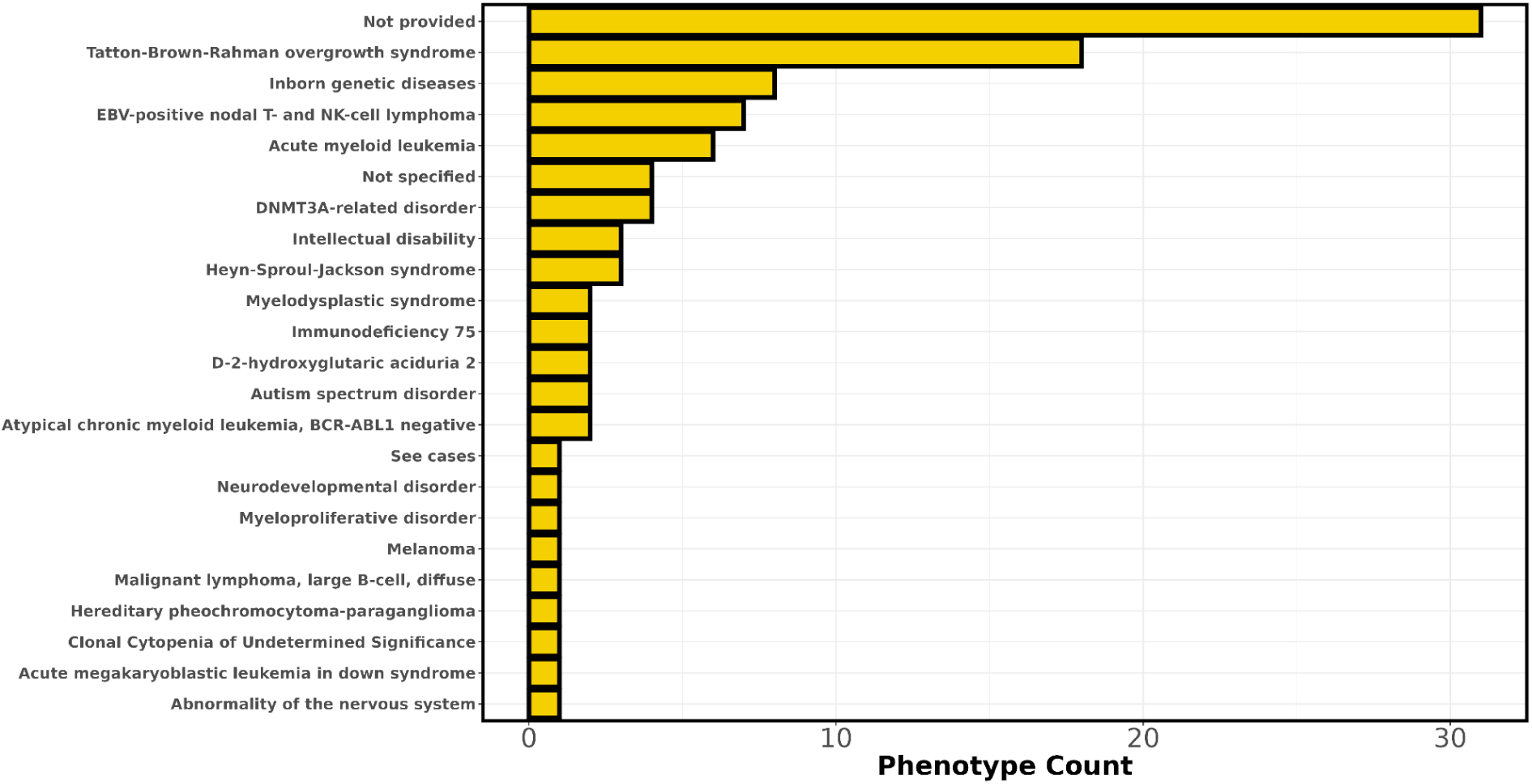
Cross-reference rare variants with associated disorders in ClinVar. Rare variants (MAF < 1% and MAC ≥ 10) identified in significant genes from the set-based tests were cross-referenced with variants in ClinVar. For variants present in ClinVar, the counts of the most commonly documented phenotype associations are displayed.

**Supplementary Figure 35.**
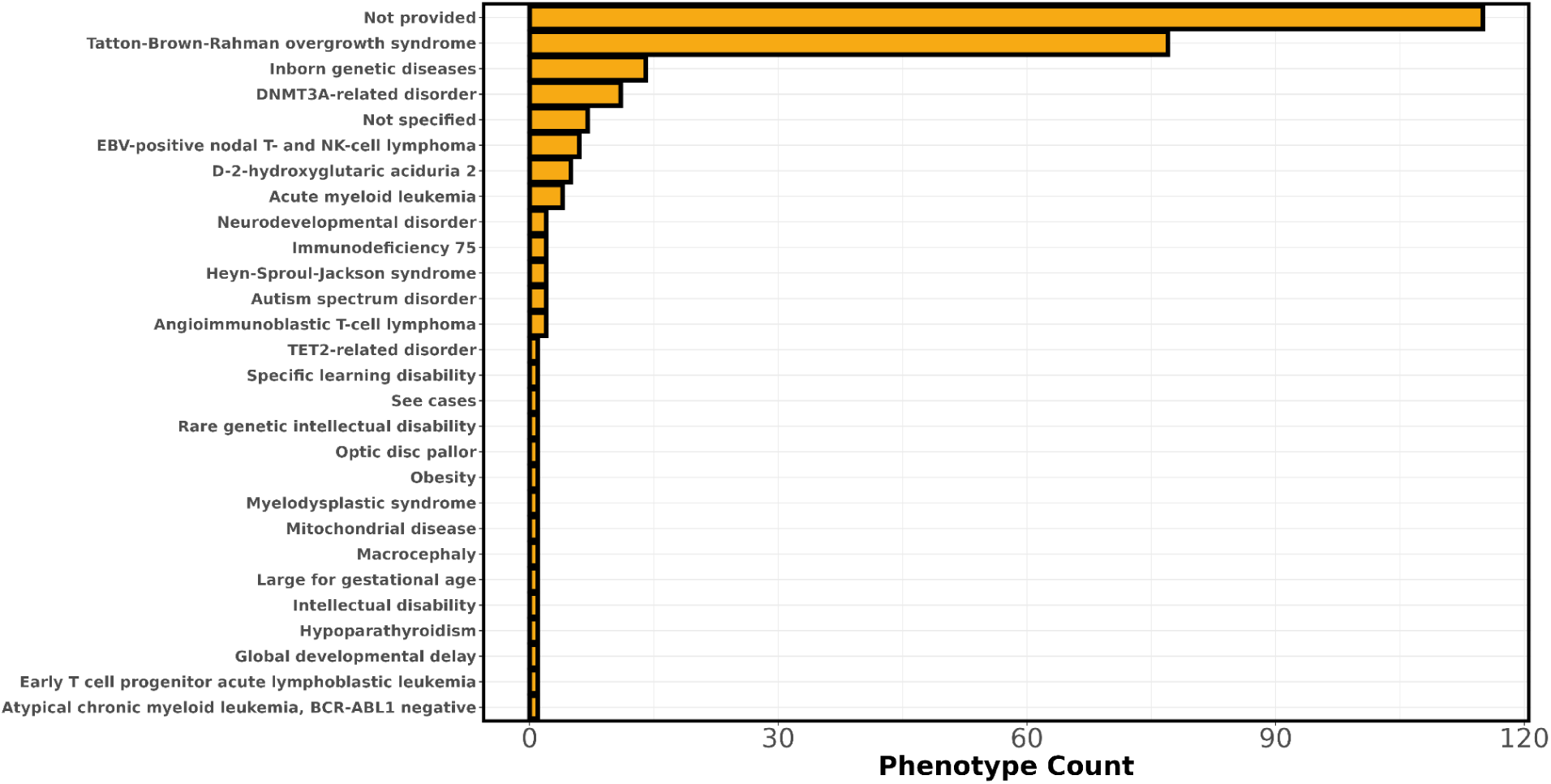
Cross-reference ultra-rare variants with associated disorders in ClinVar. Ultra-rare variants (MAC < 10) identified in significant genes from the set-based tests were cross-referenced with variants in ClinVar. For variants present in ClinVar, the counts of the most commonly documented phenotype associations are displayed.

**Supplementary Figure 36.**
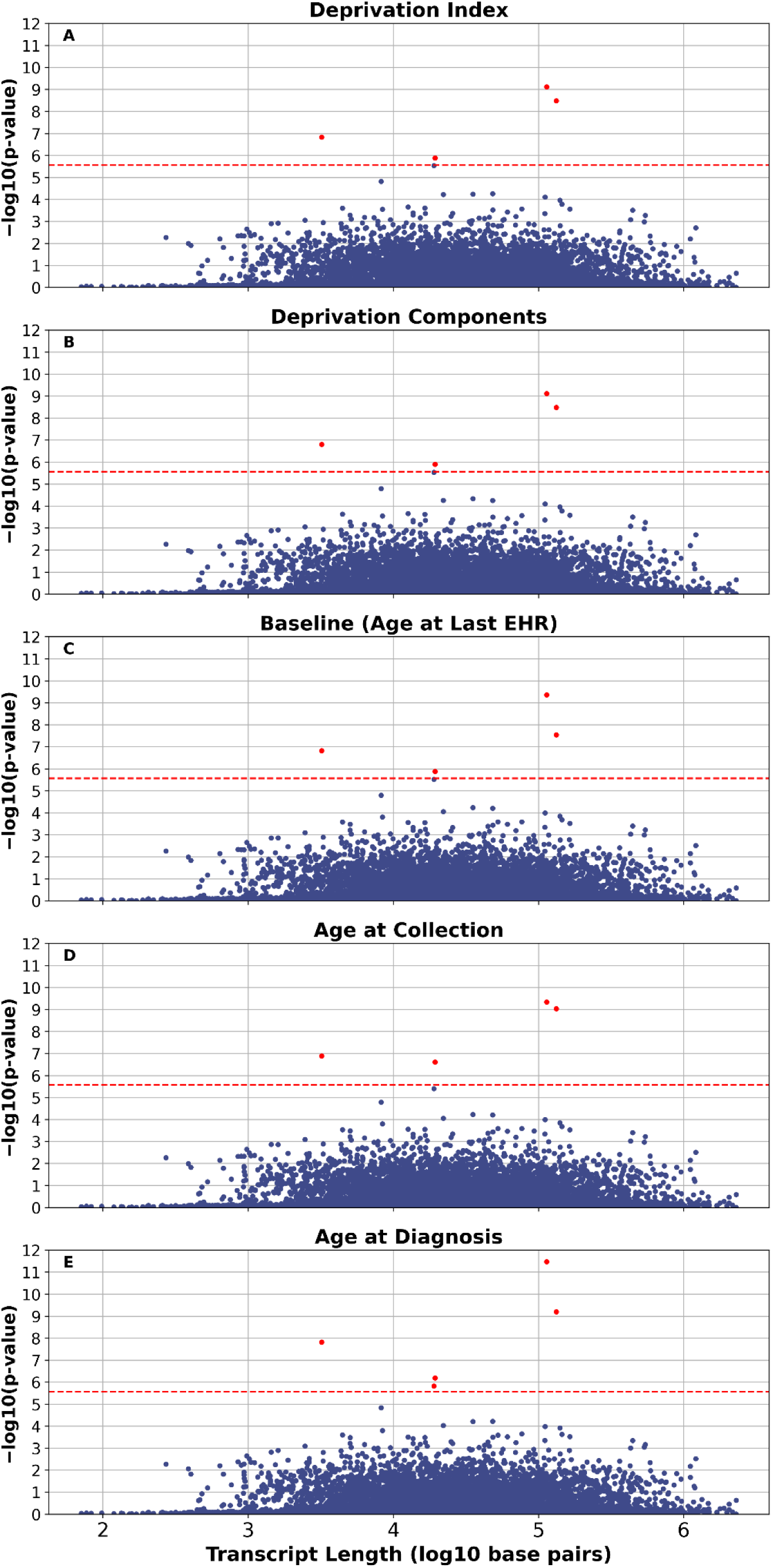
Transcript length comparison with Cauchy p-values. Cauchy SKAT-O p-values from the set-based tests against transcript length, expressed as log10 base-pair length. The “Deprivation Index” model (A) includes the deprivation index as an additional covariate, whereas the “Deprivation Components” model (B) includes the individual components of the deprivation index as covariates. The “Baseline” model (C) uses the standard set of covariates (age, sex, and principal components). The “Age at Collection” model (D) defines age based on age at biosample collection for whole-genome sequencing, while the “Age at Diagnosis” model (E) defines age using age at diagnosis for cases and age at the last EHR record for controls.

## Notes

### Competing Interest Statement

The authors have declared no competing interest.

### Author Declarations

The data used in this study are publicly available. Individual-level data from the AoU Curated Data Repository version 8 [https://www.researchallofus.org] were accessed following approval of the Data Use and Registration Agreement.

