## Supplementary Figures 1-36 for "Prioritizing Genes and Rare Protein-Coding Variants in Acute Myeloid Leukemia via Whole Genome Sequencing Data"

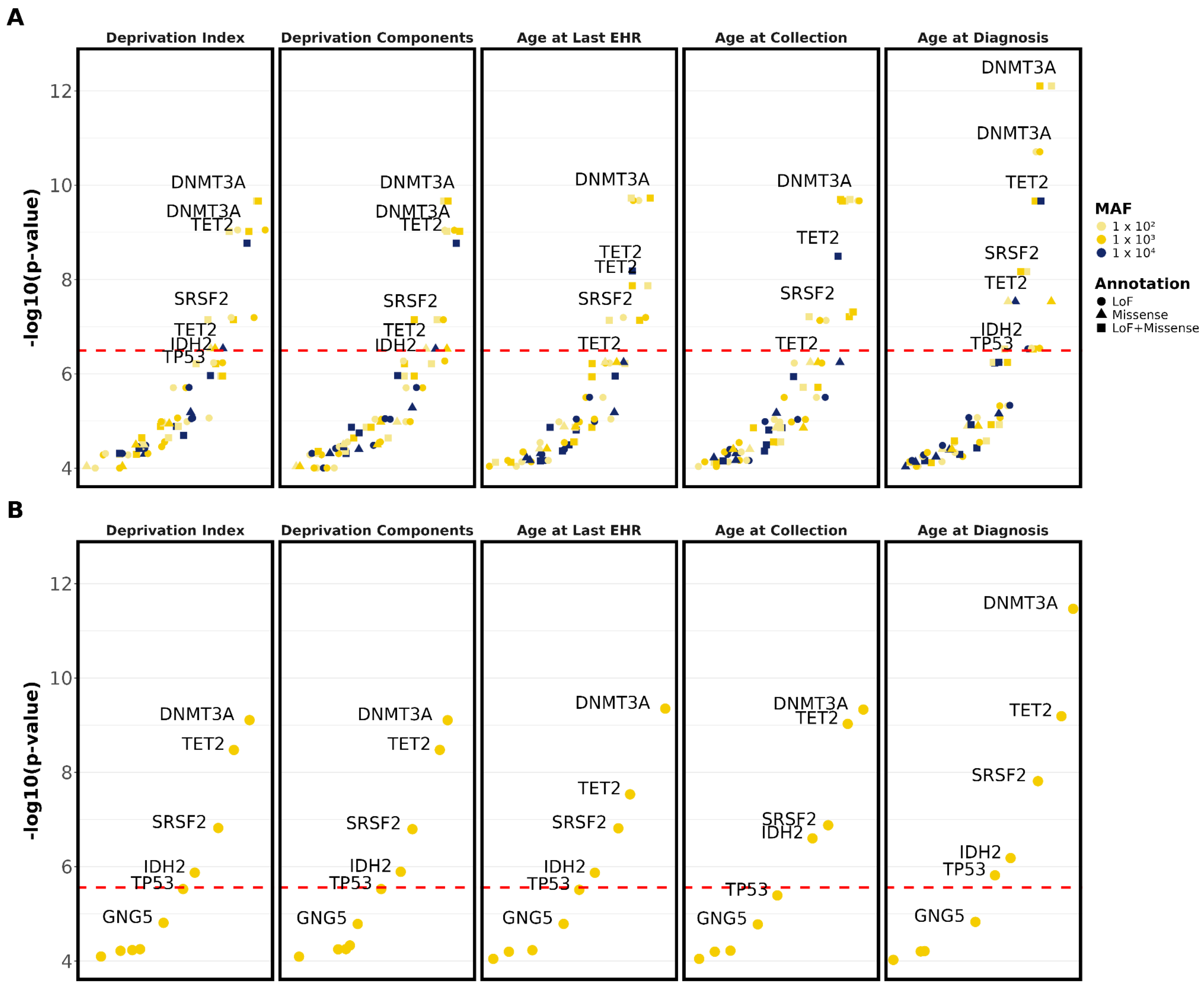


**Supplementary Figure 1. Top genes for AML set-based tests.** P-values of top genes from gene set-based tests are shown for SKAT-O analyses using non-Cauchy and Cauchy methods, stratified by analysis type. All displayed genes have p-values ≤ $1 \times 10^{-4}$. The top row (A) shows non-Cauchy SKAT-O p-values, and the bottom row (B) shows Cauchy SKAT-O p-values. The red dashed line represents genome-wide significance after multiple hypothesis correction. “Age at Last EHR” is the baseline model using standard covariates (age, sex, principal components). “Deprivation Index” and “Deprivation Components” include additional socioeconomic covariates beyond the standard covariates, where the Deprivation Index model includes the composite deprivation score and the Deprivation Components model includes its individual components. “Age at Collection” and “Age at Diagnosis” use the standard set of covariates; however, they differ from the baseline analysis in how age is defined. “Age at Collection” uses time from birth to biosample collection, and “Age at Diagnosis” uses age at diagnosis for cases and age at last EHR for controls.

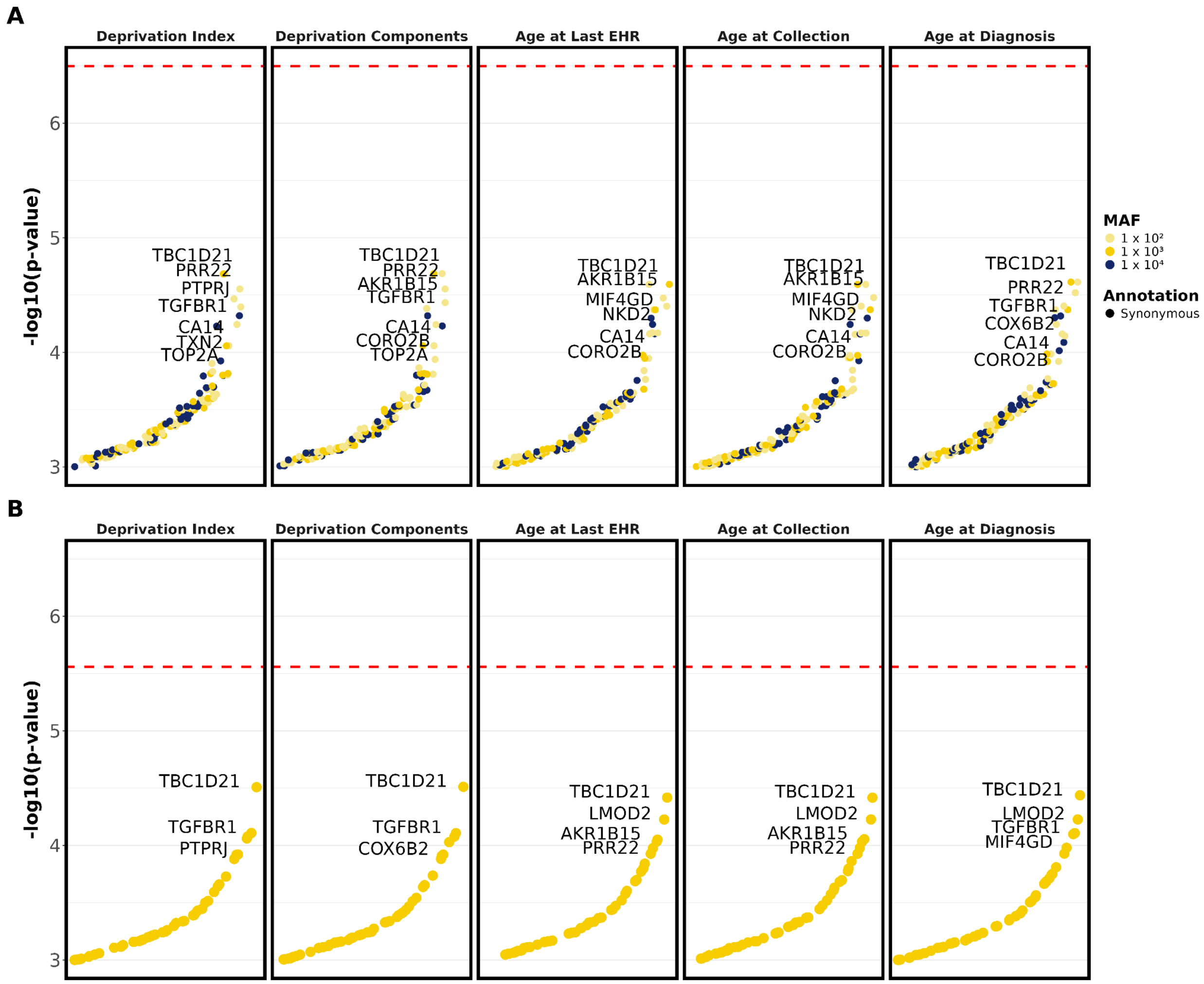


**Supplementary Figure 2. Top genes for AML synonymous set-based tests.** P-values of top genes from synonymous gene set-based tests are shown for SKAT-O analyses using non-Cauchy and Cauchy methods, stratified by analysis type. All displayed genes have p-values

≤ $1 \times 10^{-3}$. The top row (A) shows non-Cauchy SKAT-O p-values, and the bottom row (B) shows Cauchy SKAT-O p-values. The red dashed line represents genome-wide significance after multiple hypothesis correction. “Age at Last EHR” is the baseline model using standard covariates (age, sex, principal components). “Deprivation Index” and “Deprivation Components” include additional socioeconomic covariates beyond the standard covariates, where the Deprivation Index model includes the composite deprivation score and the Deprivation Components model includes its individual components. “Age at Collection” and “Age at Diagnosis” use the standard set of covariates; however, they differ from the baseline analysis in how age is defined. “Age at Collection” uses time from birth to biosample collection, and “Age at Diagnosis” uses age at diagnosis for cases and age at last EHR for controls.

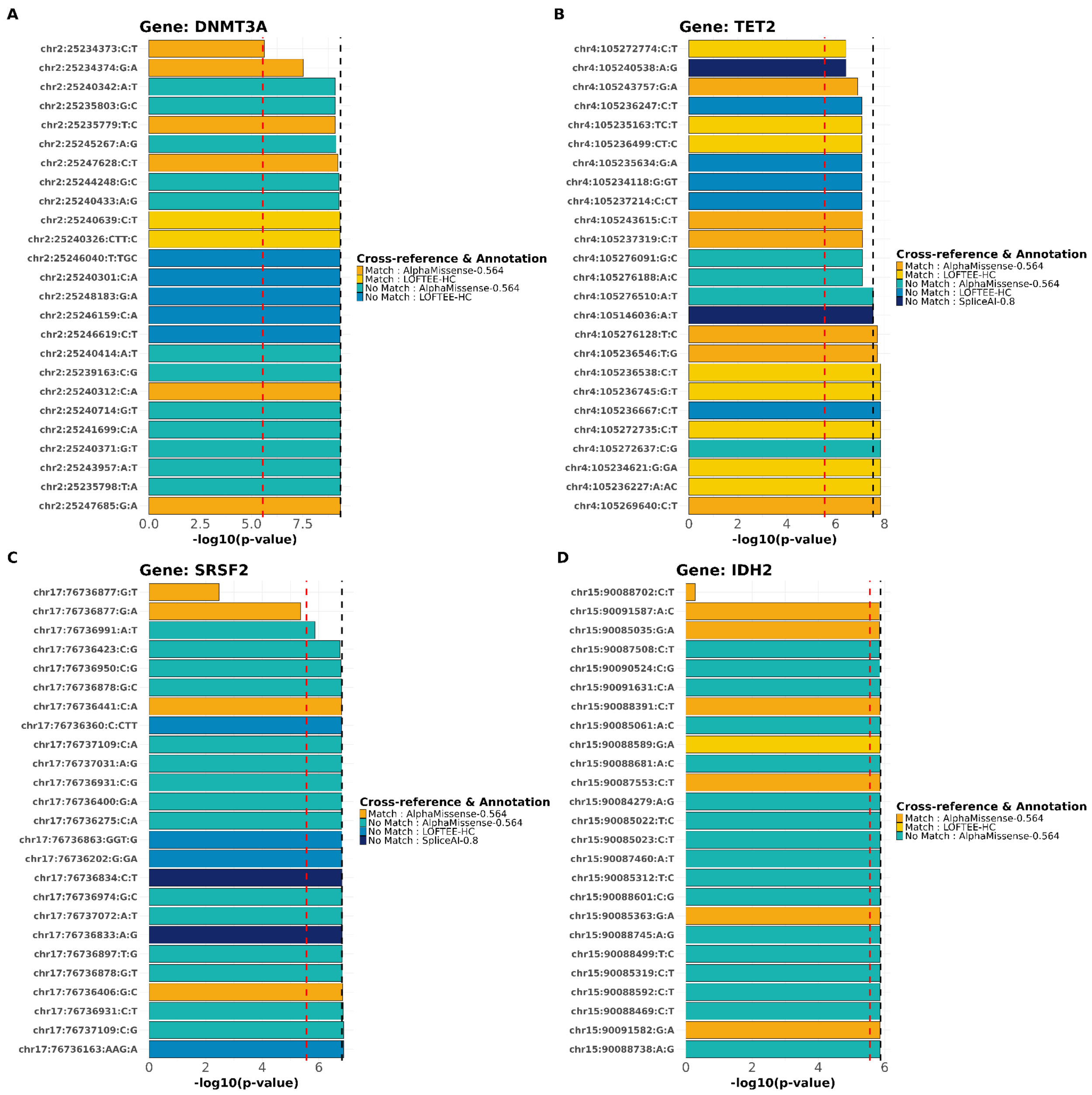


**Supplementary Figure 3. Leave-one-variant-out tests for AML.** Bar plot of Cauchy SKAT-O p-values from the leave-one-variant-out analysis of AML using standard covariates, displayed as -log10 p-values. The black dashed line represents the p-value from the standard analysis, where no variants are removed from the gene-based test. The bars display the p-values obtained after removing one variant at a time in the leave-one-variant-out analysis. The red dashed line indicates the genome-wide significance threshold. A reduction in significance such that the bar lies between the black dashed line and the red dashed line suggests that the removed variant contributes to the gene-level signal but is not solely responsible for genome-wide significance. In contrast, if the bar falls below the red dashed line, this indicates that the removed variant is largely or solely driving the genome-wide significant association for the gene.

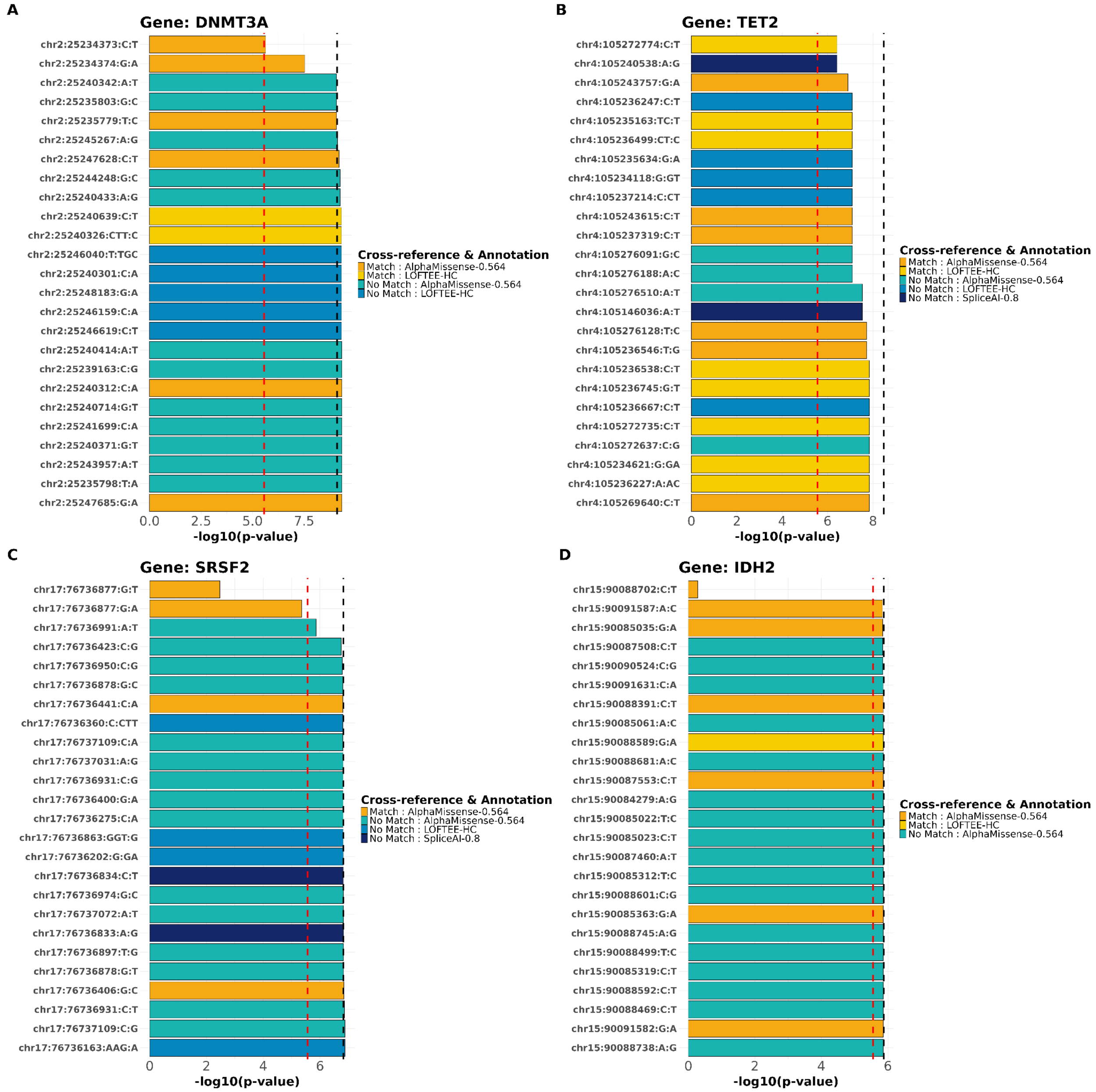


**Supplementary Figure 4. Leave-one-variant-out tests for AML adjusted for the deprivation index.** Bar plot of Cauchy SKAT-O p-values from the leave-one-variant-out analysis of AML using standard covariates plus the deprivation index, displayed as -log10 p-values. The black dashed line represents the p-value from the standard analysis, where no variants are removed from the gene-based test. The bars display the p-values obtained after removing one variant at a time in the leave-one-variant-out analysis. The red dashed line indicates the genome-wide significance threshold. A reduction in significance such that the bar lies between the black dashed line and the red dashed line suggests that the removed variant contributes to the gene-level signal but is not solely responsible for genome-wide significance. In contrast, if the bar falls below the red dashed line, this indicates that the removed variant is largely or solely driving the genome-wide significant association for the gene.

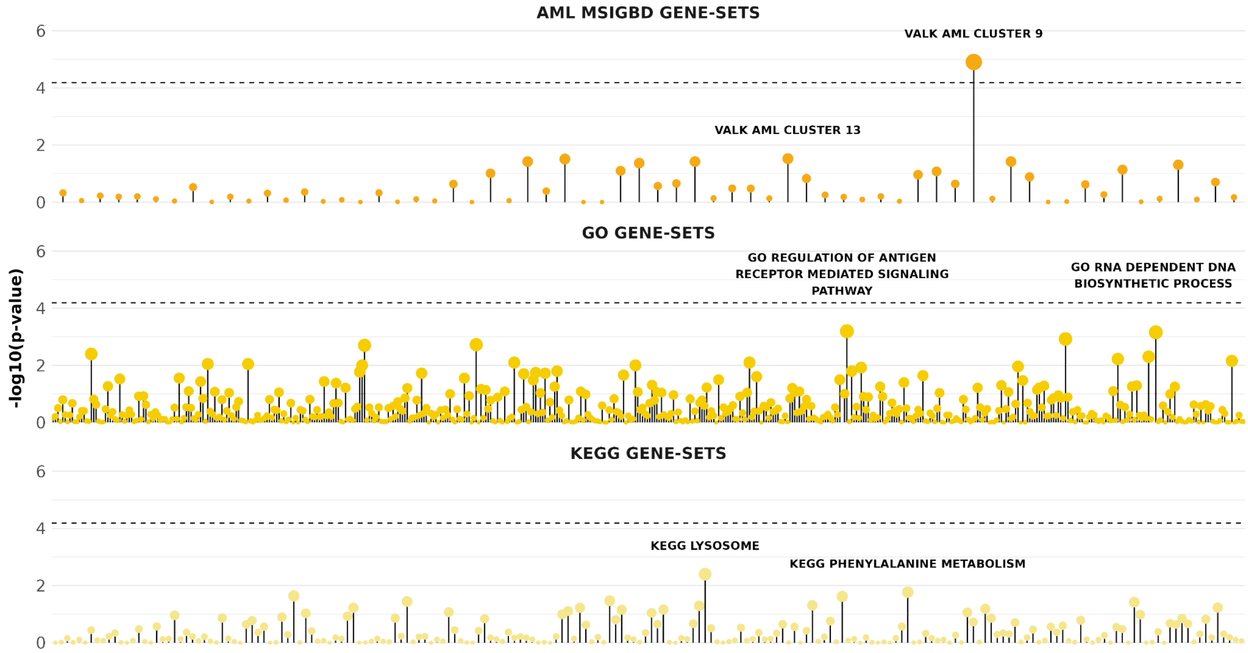


**Supplementary Figure 5. Gene-set results from GAUSS for MSigDB, GO, and KEGG.** The plot displays -log10 p-values from gene-set association analyses conducted using GAUSS. The dashed black line represents the significance threshold after multiple hypothesis correction across all gene sets. Analyses were performed using both the GO (middle panel) and KEGG (bottom panel) pathway collections. Additionally, gene sets from the MSigDB were evaluated; however, due to overlap among MSigDB gene sets, the analysis was restricted to AML-related gene sets. In total, 64 AML-specific gene sets were tested (top panel).

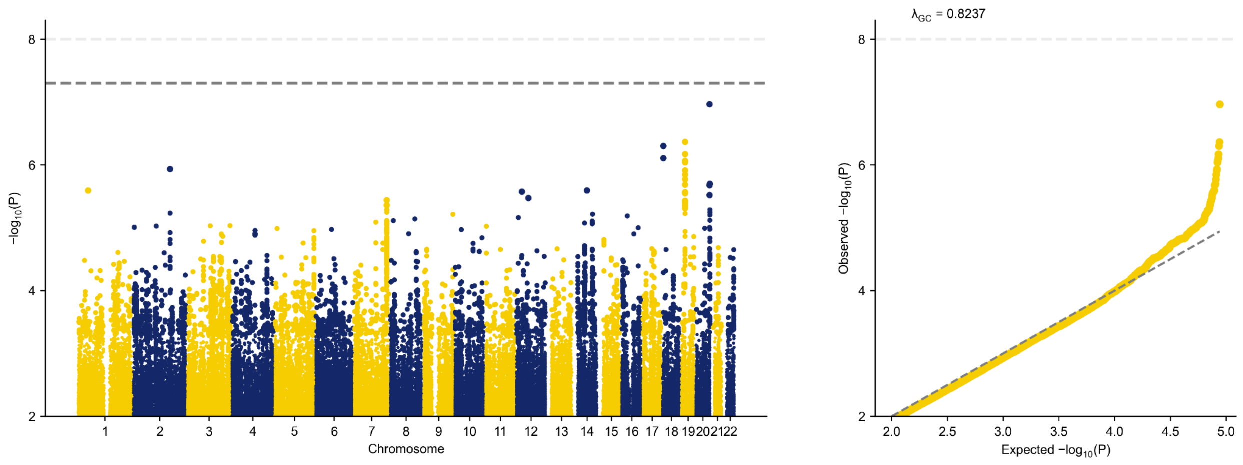


**Supplementary Figure 6. Single-variant tests for AML.** Manhattan plot (left panel) and quantile-quantile (QQ) plot (right panel) for common variants (MAF > 1%) in AML, adjusted for the standard set of covariates. Genome-wide significance was defined as $p < 5 \times 10^{-8}$ and is indicated by the dashed light grey line. The QQ plot also illustrates the genomic inflation of the test statistics.

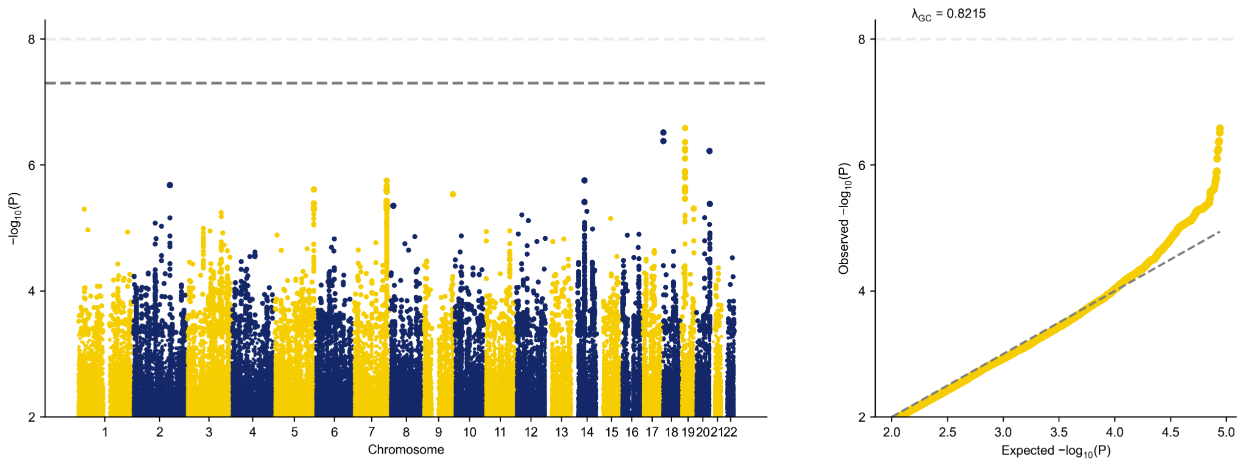


**Supplementary Figure 7. Single-variant tests for AML adjusted for the deprivation index.** Manhattan plot (left panel) and quantile-quantile (QQ) plot (right panel) for common variants (MAF > 1%) in AML, adjusted for the standard set of covariates plus the deprivation index.

Genome-wide significance was defined as $p < 5 \times 10^{-8}$ and is indicated by the dashed light grey line. The QQ plot also illustrates the genomic inflation of the test statistics.

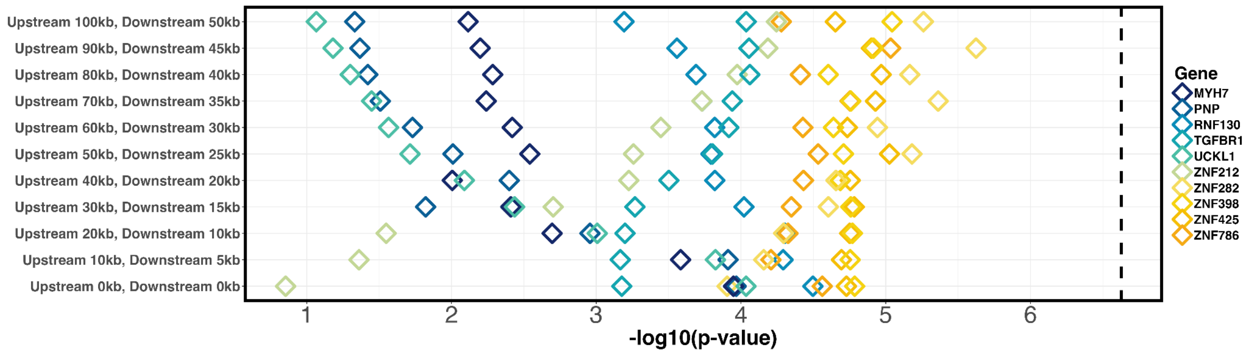


**Supplementary Figure 8. MAGMA gene tests for AML.** Using genome-wide association summary statistics for AML, adjusted for the standard set of covariates, MAGMA analysis was conducted to evaluate window-size effects across 11 iterations. The analysis began with 0 kb upstream and 0 kb downstream windows. At each subsequent iteration, the upstream window size was increased by 10 kb and the downstream window size by 5 kb, reaching final windows of 100 kb upstream and 50 kb downstream. The -log10 p-values for the top 10 genes from MAGMA gene-based association tests are displayed for each window configuration. The dashed black line represents the significance threshold after multiple hypothesis correction.

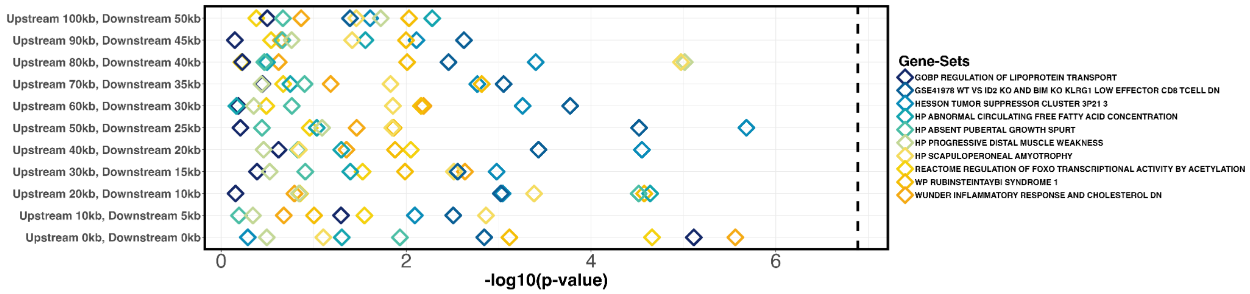


**Supplementary Figure 9. MAGMA gene-set tests for AML.** Using genome-wide association summary statistics for AML, adjusted for the standard set of covariates, MAGMA analysis was conducted to evaluate window-size effects across 11 iterations. The analysis began with 0 kb upstream and 0 kb downstream windows. At each subsequent iteration, the upstream window size was increased by 10 kb and the downstream window size by 5 kb, reaching final windows of 100 kb upstream and 50 kb downstream. The -log10 p-values for the top 10 gene-sets from MAGMA gene-set association tests are displayed for each window configuration. The dashed black line represents the significance threshold after multiple hypothesis correction.

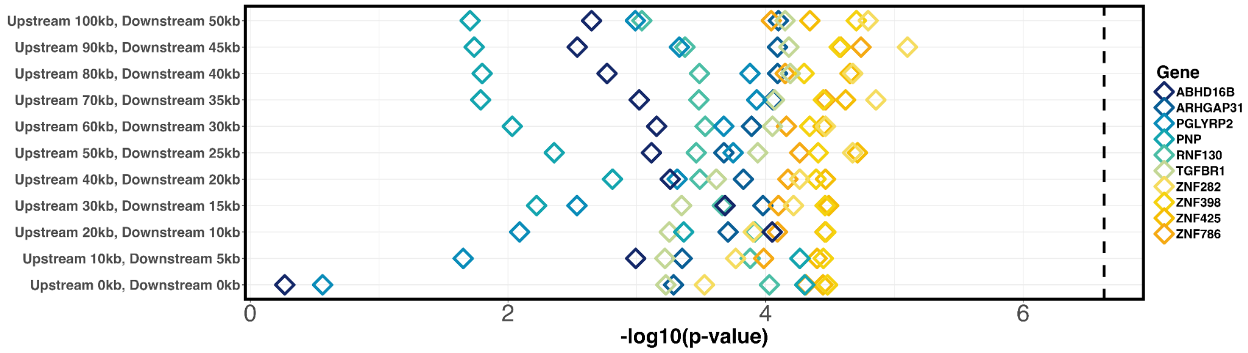


**Supplementary Figure 10. MAGMA gene tests for AML adjusted for the deprivation index.** Using genome-wide association summary statistics for AML, adjusted for the standard set of covariates plus the deprivation index, MAGMA analysis was conducted to evaluate window-size effects across 11 iterations. The analysis began with 0 kb upstream and 0 kb downstream windows. At each subsequent iteration, the upstream window size was increased by 10 kb and the downstream window size by 5 kb, reaching final windows of 100 kb upstream and 50 kb downstream. The -log10 p-values for the top 10 genes from MAGMA gene-based association tests are displayed for each window configuration. The dashed black line represents the significance threshold after multiple hypothesis correction.

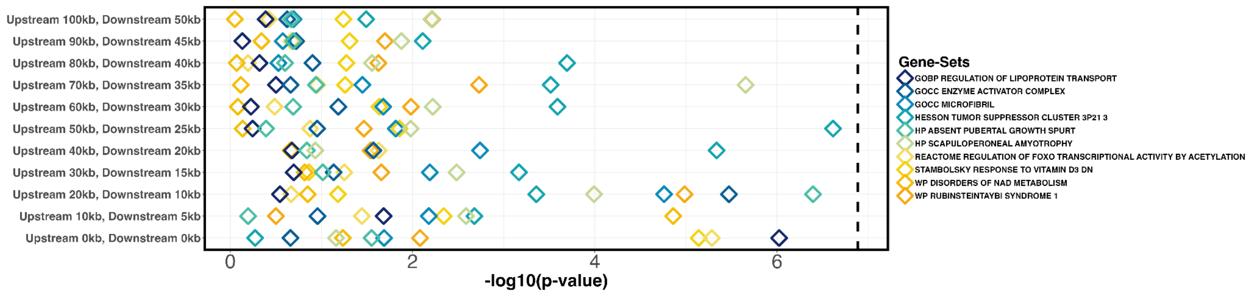


**Supplementary Figure 11. Supplementary Figure. MAGMA gene-set tests for AML adjusted for the deprivation index.** Using genome-wide association summary statistics for AML, adjusted for the standard set of covariates plus the deprivation index, MAGMA analysis was conducted to evaluate window-size effects across 11 iterations. The analysis began with 0 kb upstream and 0 kb downstream windows. At each subsequent iteration, the upstream window size was increased by 10 kb and the downstream window size by 5 kb, reaching final windows of 100 kb upstream and 50 kb downstream. The -log10 p-values for the top 10 gene-sets from MAGMA gene-set association tests are displayed for each window configuration. The dashed black line represents the significance threshold after multiple hypothesis correction.

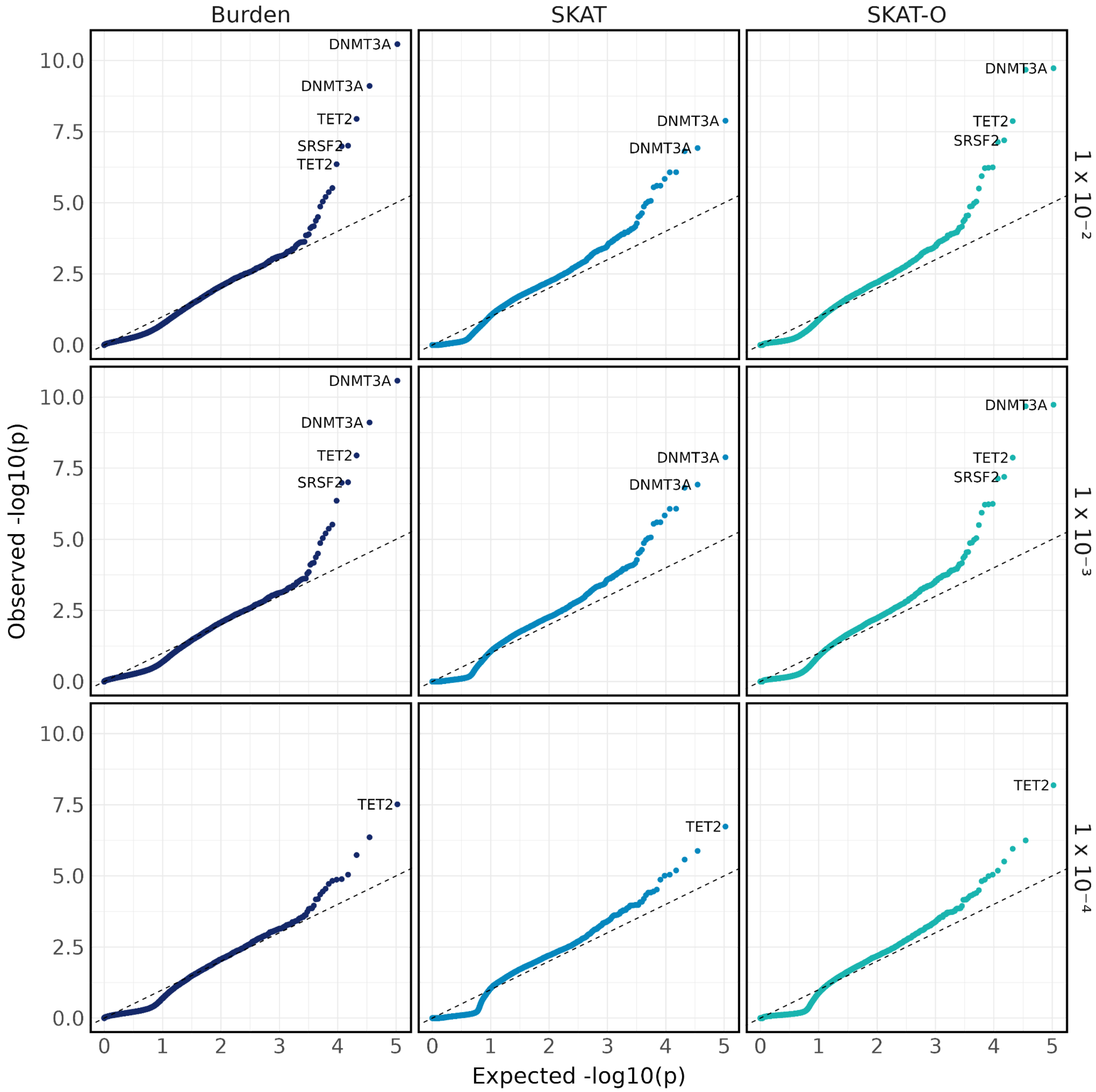


**Supplementary Figure 12. Quantile-quantile plot of AML set-based tests.** Quantile-quantile plot of p-values from set-based tests, stratified by test type (Burden, SKAT, and SKAT-O) and maximum MAF thresholds (1%, 0.1%, and 0.01%).

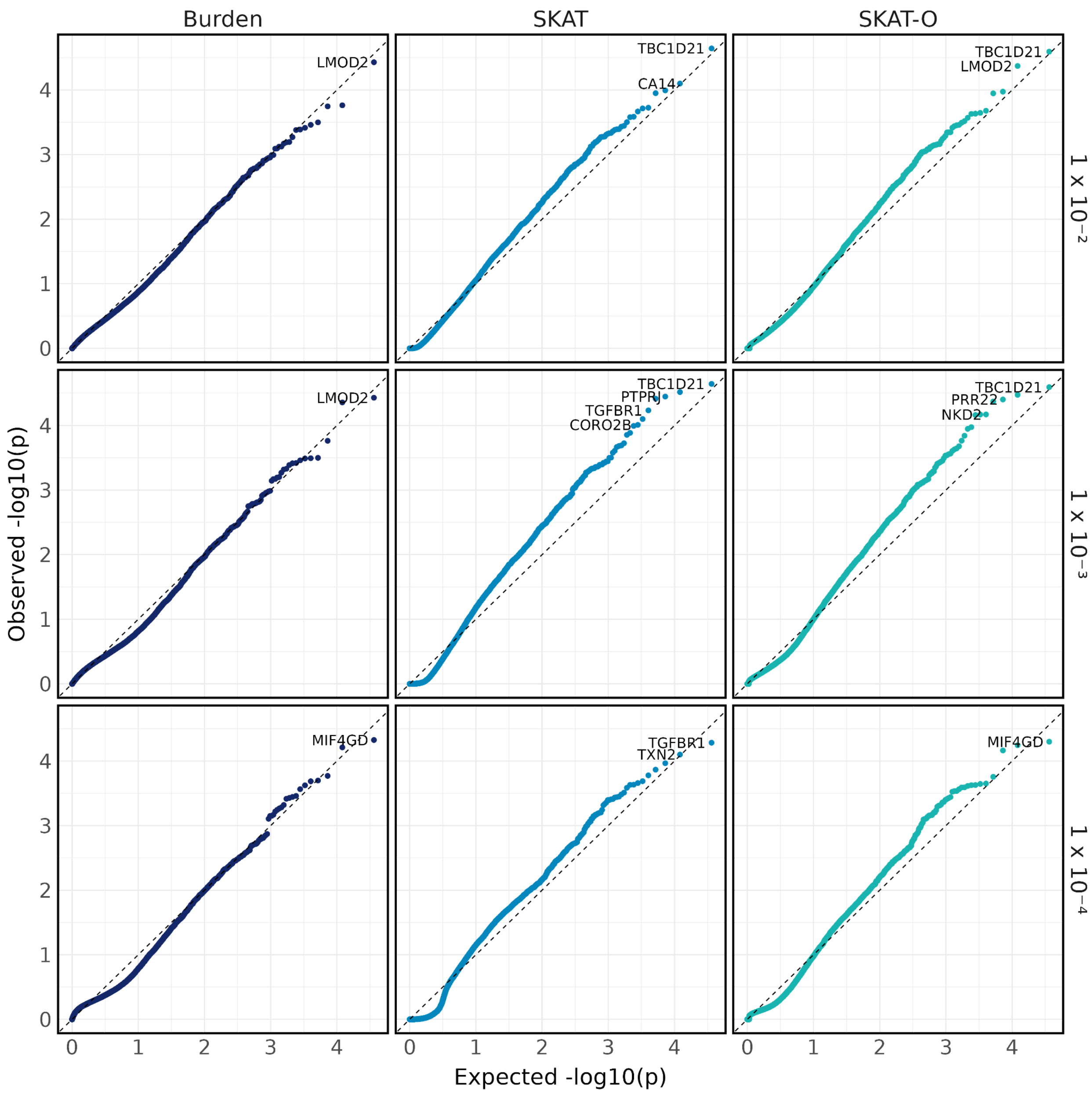


**Supplementary Figure 13. Quantile-quantile plot of AML synonymous set-based tests.**

Quantile-quantile plot of p-values from synonymous set-based tests, stratified by test type (Burden, SKAT, and SKAT-O) and maximum MAF thresholds (1%, 0.1%, and 0.01%).

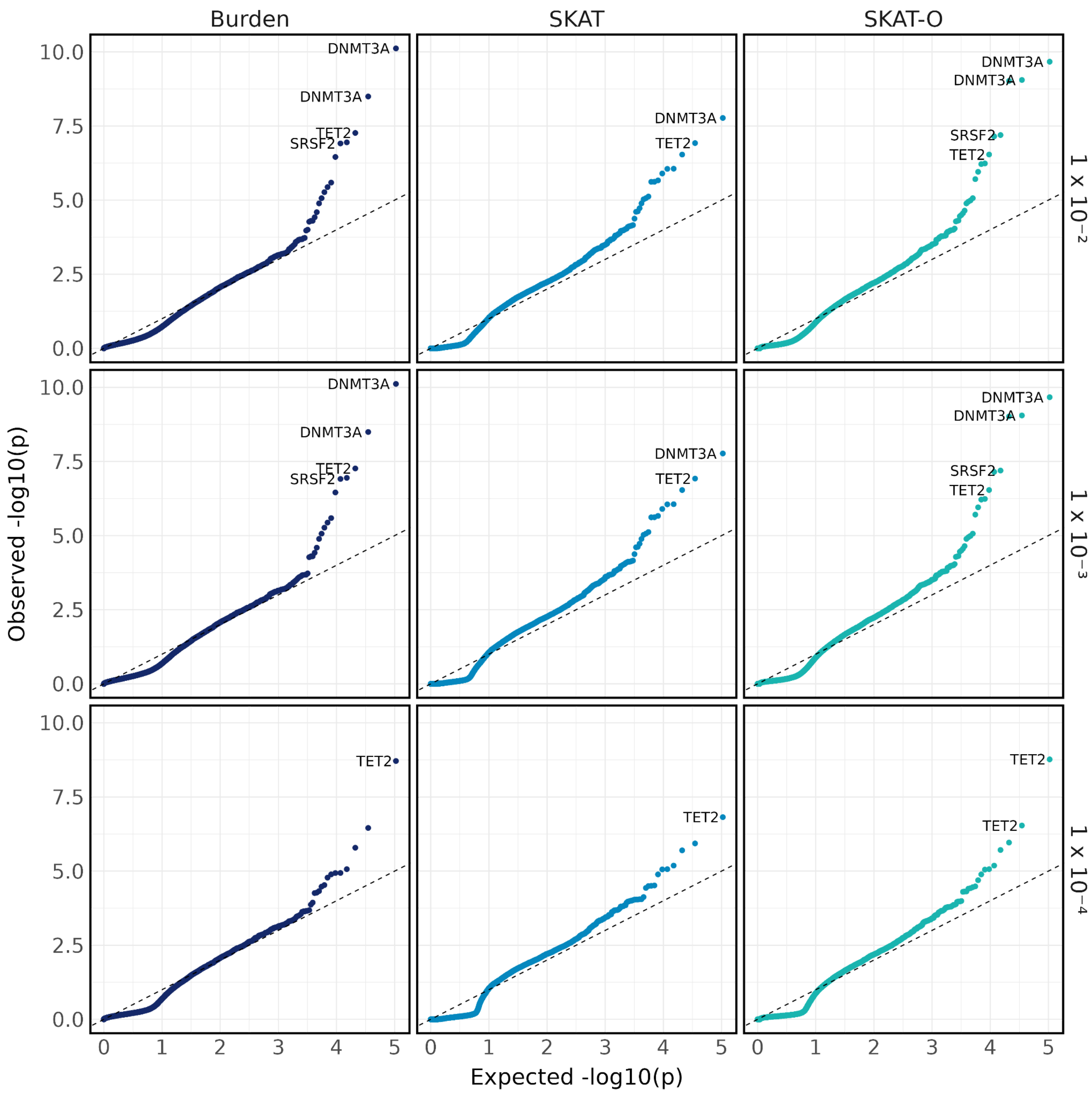


**Supplementary Figure 14. Quantile-quantile plot of AML set-based tests adjusted for the deprivation index.** Quantile-quantile plot of p-values from set-based tests adjusting for the deprivation index, stratified by test type (Burden, SKAT, and SKAT-O) and maximum MAF thresholds (1%, 0.1%, and 0.01%).

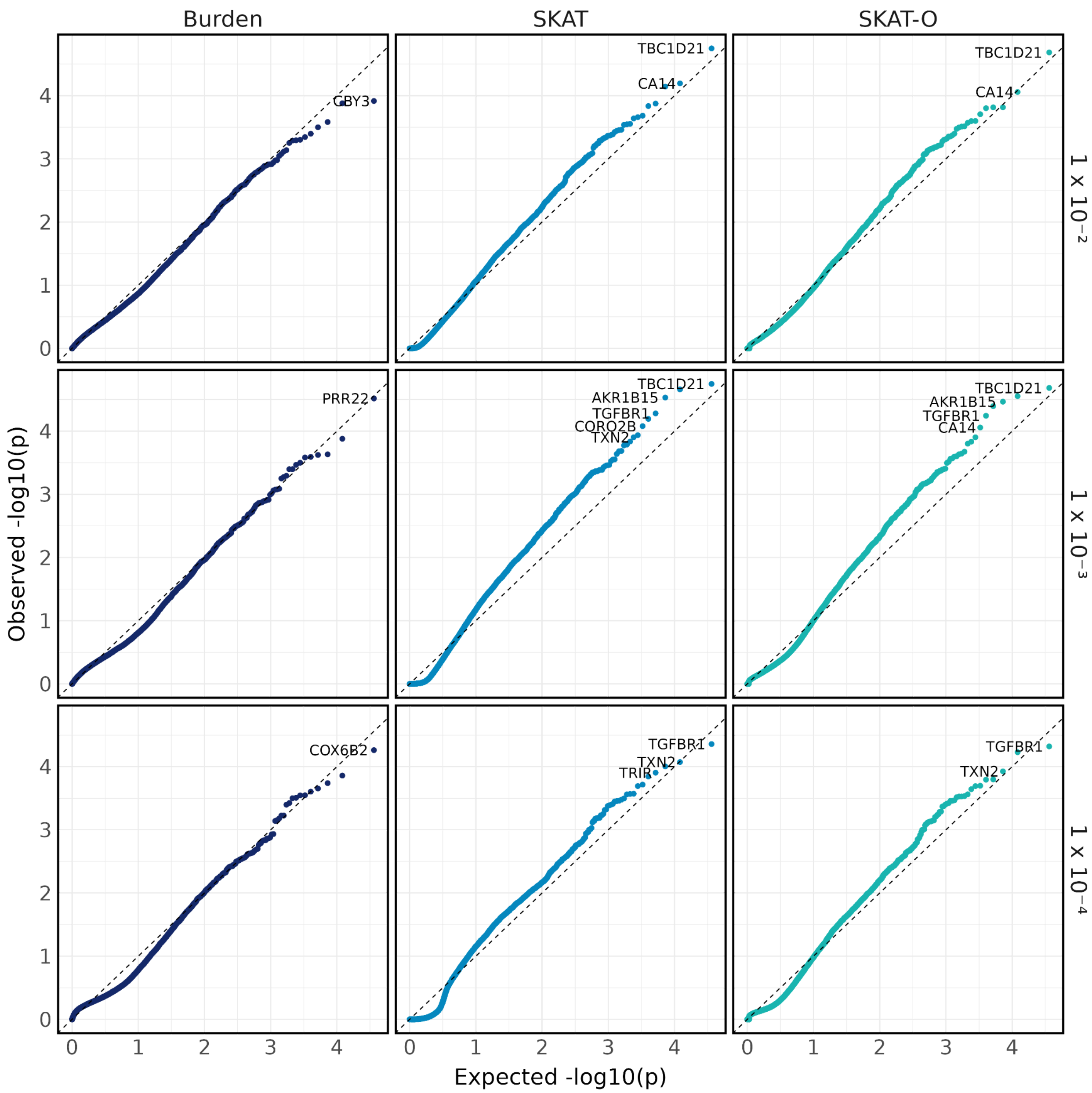


**Supplementary Figure 15. Quantile-quantile plot of AML synonymous set-based tests adjusted for the deprivation index.** Quantile-quantile plot of p-values from synonymous set-based tests adjusting for the deprivation index, stratified by test type (Burden, SKAT, and SKAT-O) and maximum MAF thresholds (1%, 0.1%, and 0.01%).

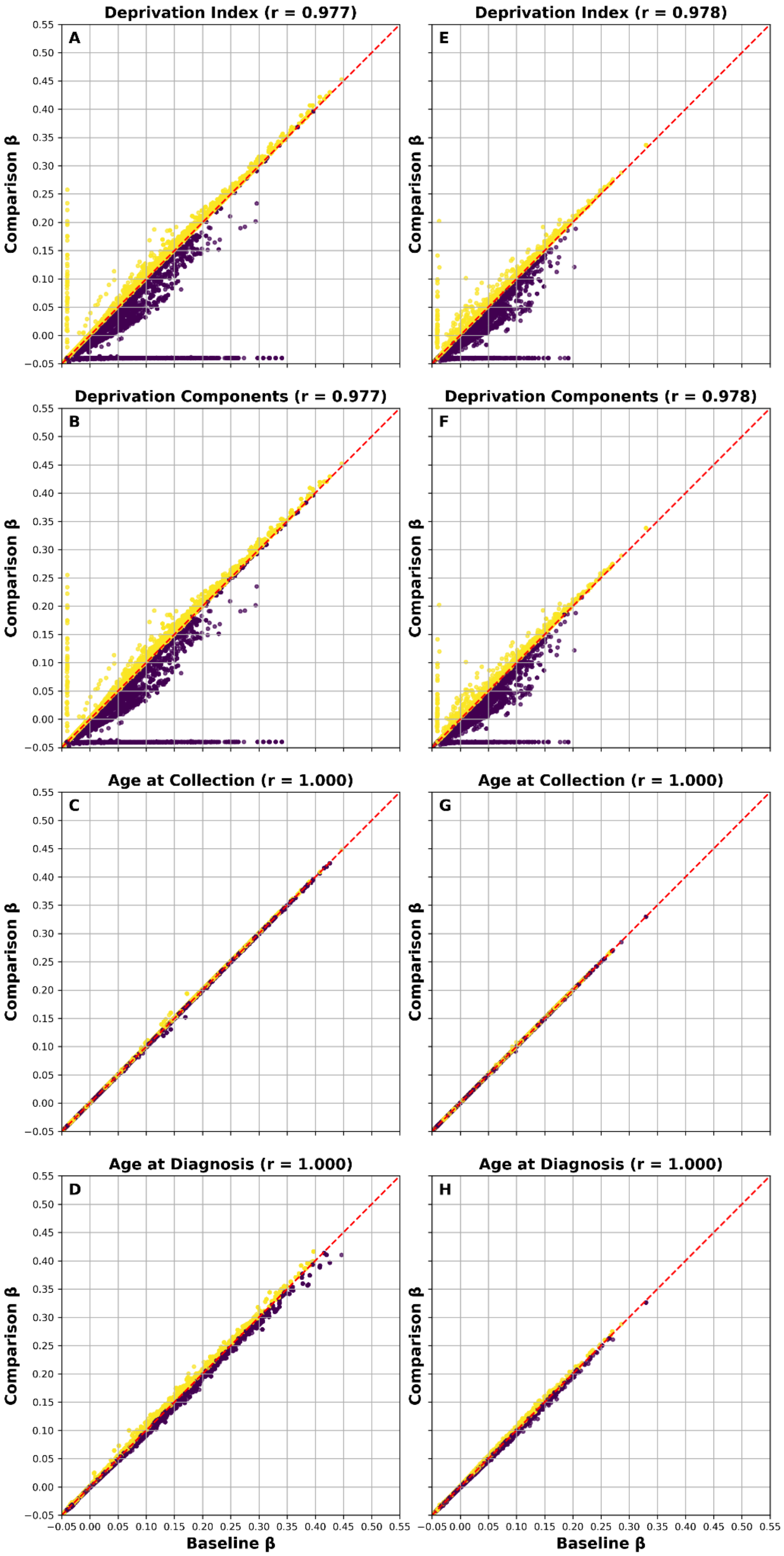
Deprivation Index (r = 0.977)
Deprivation Index (r = 0.978)
Deprivation Components (r = 0.977)
Deprivation Components (r = 0.978)
Age at Collection (r = 1.000)
Age at Collection (r = 1.000)
Age at Diagnosis (r = 1.000)
Age at Diagnosis (r = 1.000)
A
B
C
D
E
F
G
H
Comparison β
Baseline β
−0.05 0.00 0.05 0.10 0.15 0.20 0.25 0.30 0.35 0.40 0.45 0.50 0.55

**Supplementary Figure 16. Correlations between burden coefficients from set-based tests.** Baseline gene set-based tests, which used the standard set of covariates (age, sex, and principal components), were compared against models in which the covariates were modified. The “Deprivation Index” model (A & E) includes the deprivation index as an additional covariate, whereas the “Deprivation Components” model (B & F) includes the individual components of the deprivation index as covariates. The “Age at Collection” model (C & G) defines age based on age at biosample collection for whole-genome sequencing, while the “Age at Diagnosis” model (D & H) defines age using age at diagnosis for cases and age at the last EHR record for controls. The plots on the left (A-D) represent tests using the variant set predicted to be deleterious (loss-of-function or missense), while plots on the right (E-H) represent tests employing the synonymous variant set. The Pearson correlation coefficient was calculated and is reported at the top of each plot.

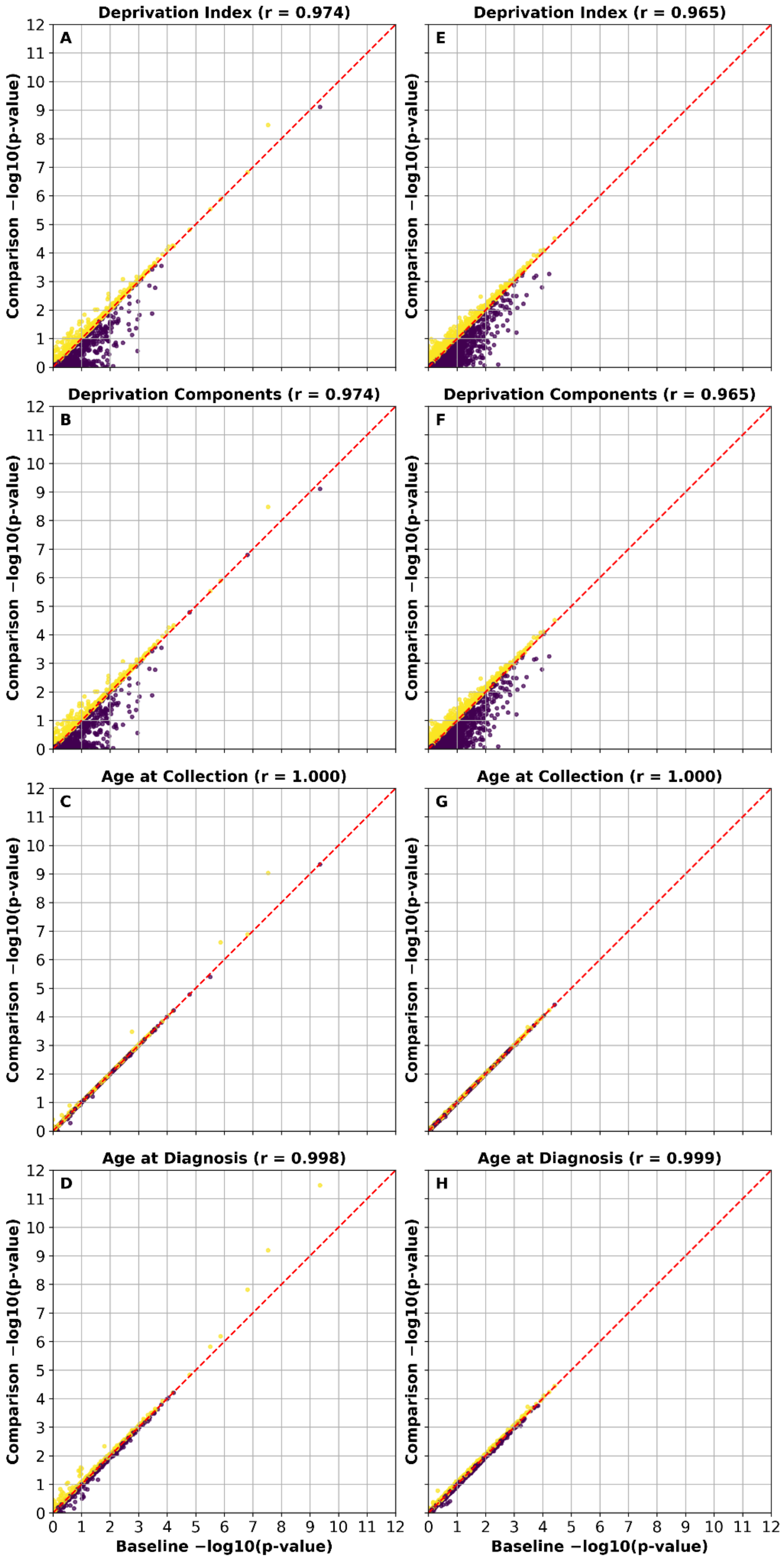
Deprivation Index (r = 0.974)
A
Deprivation Index (r = 0.965)
E
Deprivation Components (r = 0.974)
B
Deprivation Components (r = 0.965)
F
Age at Collection (r = 1.000)
C
Age at Collection (r = 1.000)
G
Age at Diagnosis (r = 0.998)
D
Age at Diagnosis (r = 0.999)
H
Comparison −log10(p-value)
Baseline −log10(p-value)

**Supplementary Figure 17. Correlations between Cauchy SKAT-O p-values from set-based tests.** Baseline gene set-based tests, which used the standard set of covariates (age, sex, and principal components), were compared against models in which the covariates were modified. The “Deprivation Index” model (A & E) includes the deprivation index as an additional covariate, whereas the “Deprivation Components” model (B & F) includes the individual components of the deprivation index as covariates. The “Age at Collection” model (C & G) defines age based on age at biosample collection for whole-genome sequencing, while the “Age at Diagnosis” model (D & H) defines age using age at diagnosis for cases and age at the last EHR record for controls. The plots on the left (A-D) represent tests using the variant set predicted to be deleterious (loss-of-function or missense), while plots on the right (E-H) represent tests employing the synonymous variant set. The Pearson correlation coefficient was calculated and is reported at the top of each plot.

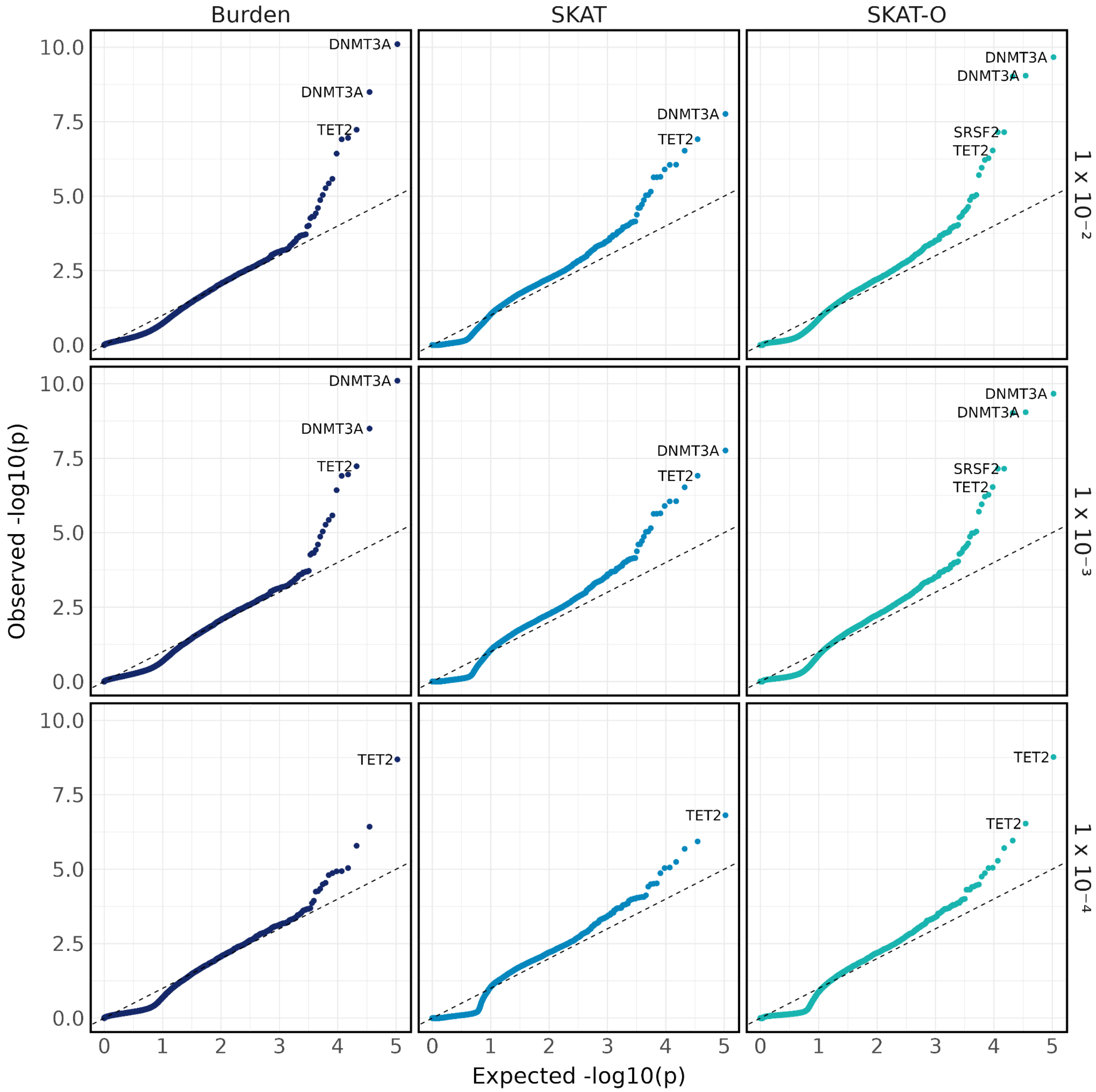


**Supplementary Figure 18. Quantile-quantile plot of AML set-based tests adjusted for the deprivation components.** Quantile-quantile plot of p-values from set-based tests adjusting for the components of the deprivation index, stratified by test type (Burden, SKAT, and SKAT-O) and maximum MAF thresholds (1%, 0.1%, and 0.01%).

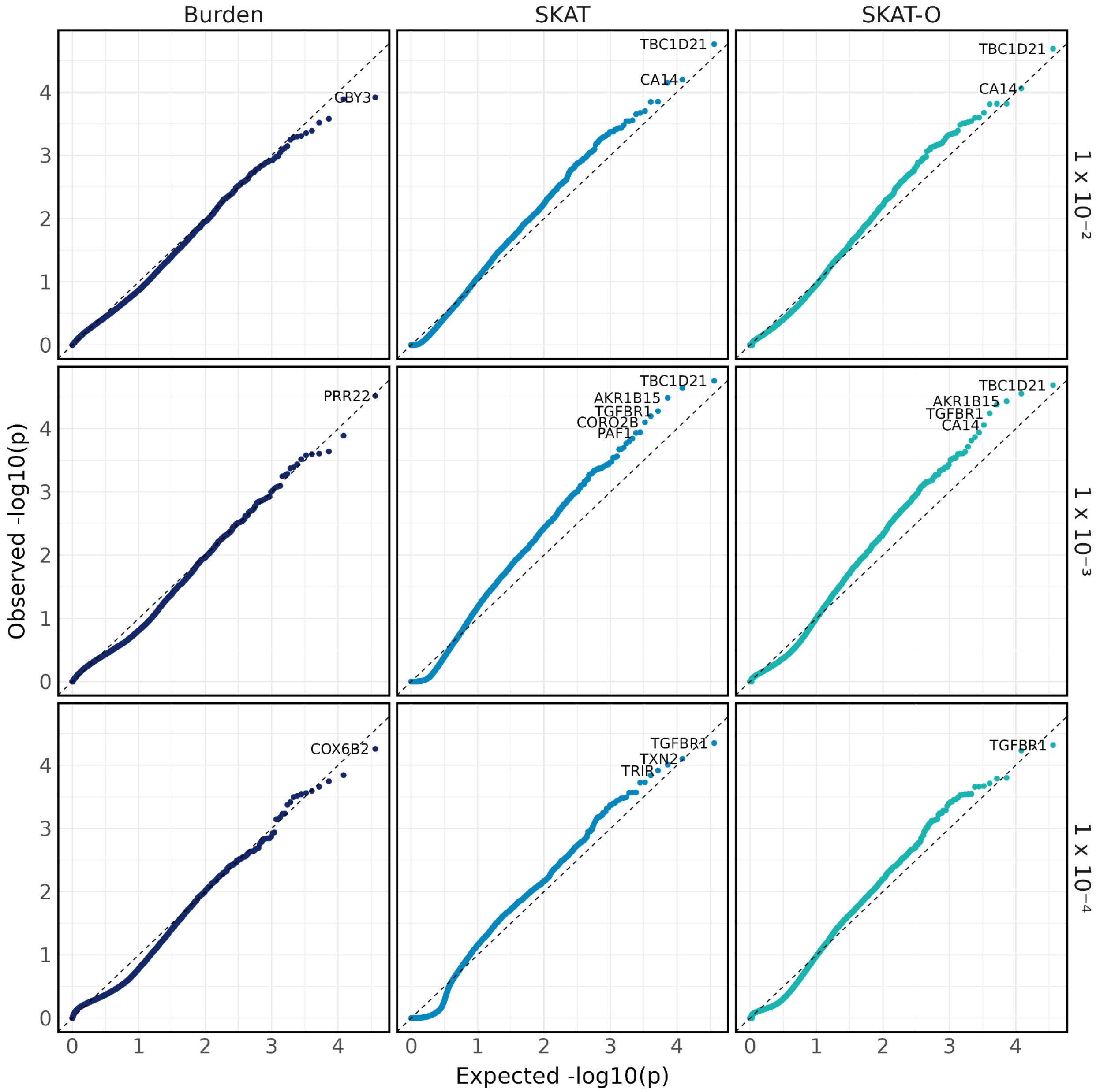


**Supplementary Figure 19. Quantile-quantile plot of AML synonymous set-based tests adjusted for the deprivation components.** Quantile-quantile plot of p-values from synonymous set-based tests adjusting for the components of the deprivation index, stratified by test type (Burden, SKAT, and SKAT-O) and maximum MAF thresholds (1%, 0.1%, and 0.01%).

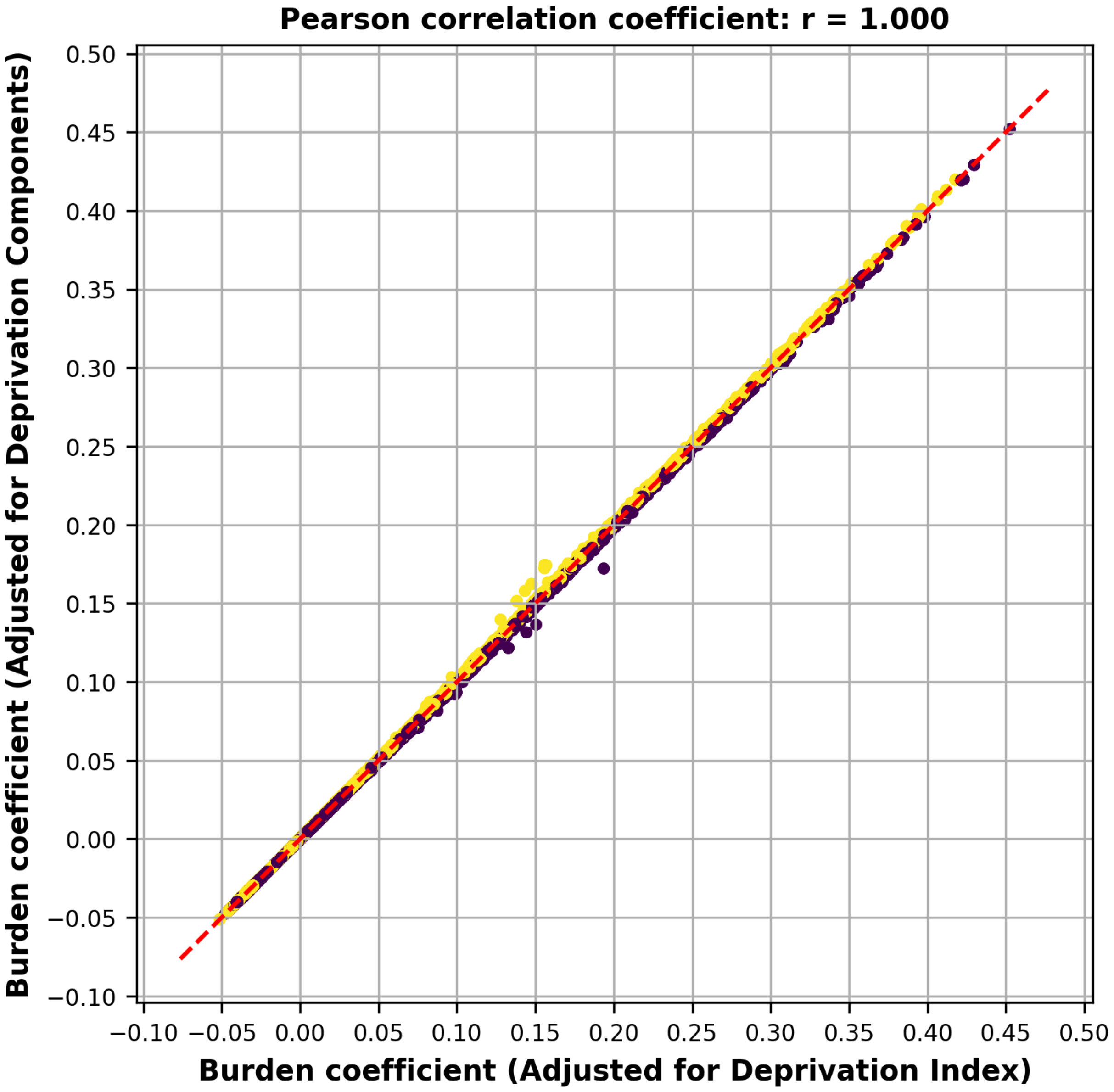


**Supplementary Figure 20. Burden coefficients from set-based tests adjusted for the deprivation index versus the deprivation components.** Correlations between Burden coefficients p-values from the set-based tests are shown, with models adjusted for the deprivation index and for the individual components of the deprivation index as covariates. The Pearson correlation coefficient was calculated and is reported at the top of each plot.

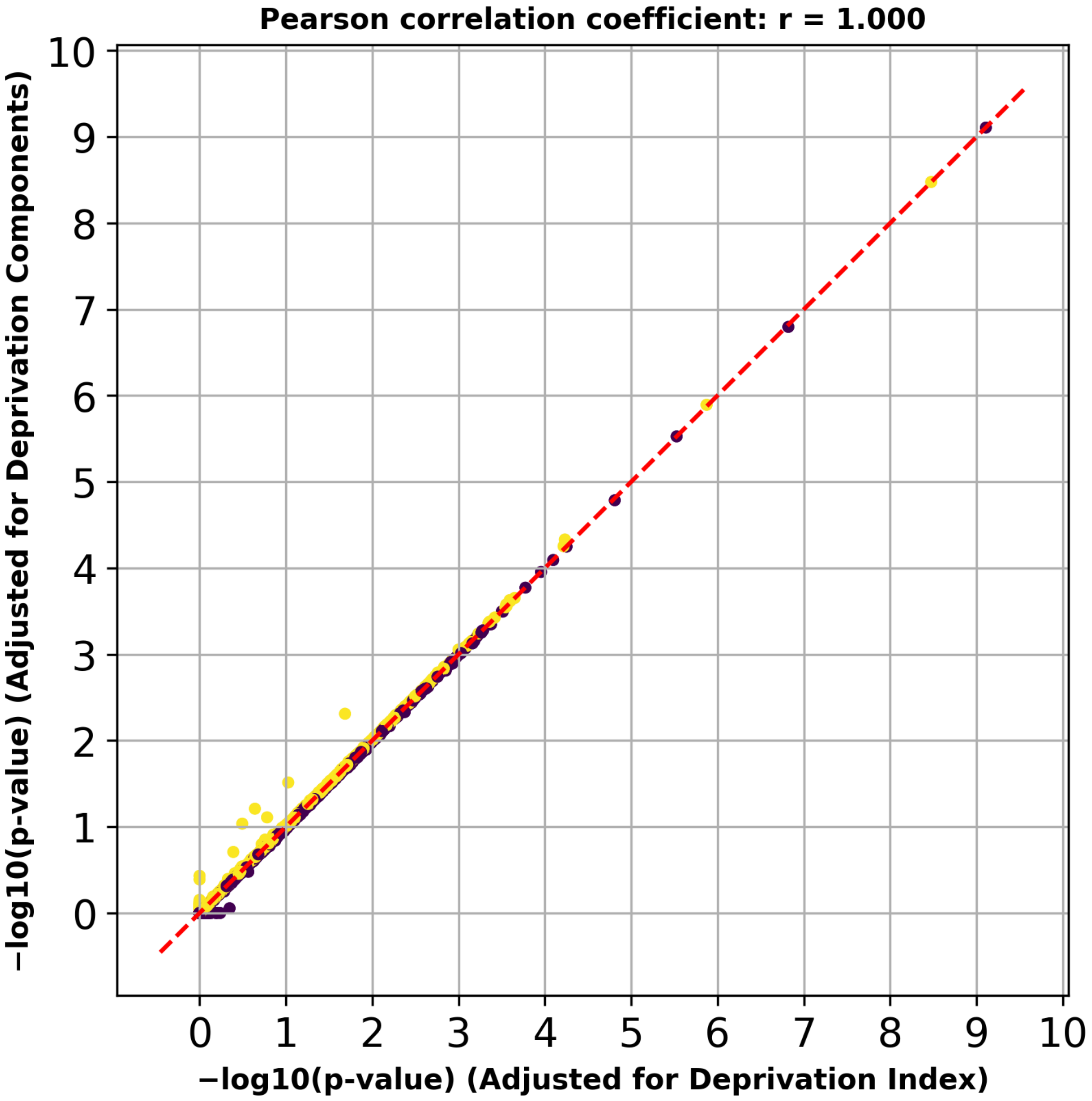


**Supplementary Figure 21. Cauchy SKAT-O p-values from set-based tests adjusted for the deprivation index versus the deprivation components.** Correlations between Cauchy SKAT-O p-values from the set-based tests are shown, with models adjusted for the deprivation index and for the individual components of the deprivation index as covariates. The Pearson correlation coefficient was calculated and is reported at the top of each plot.

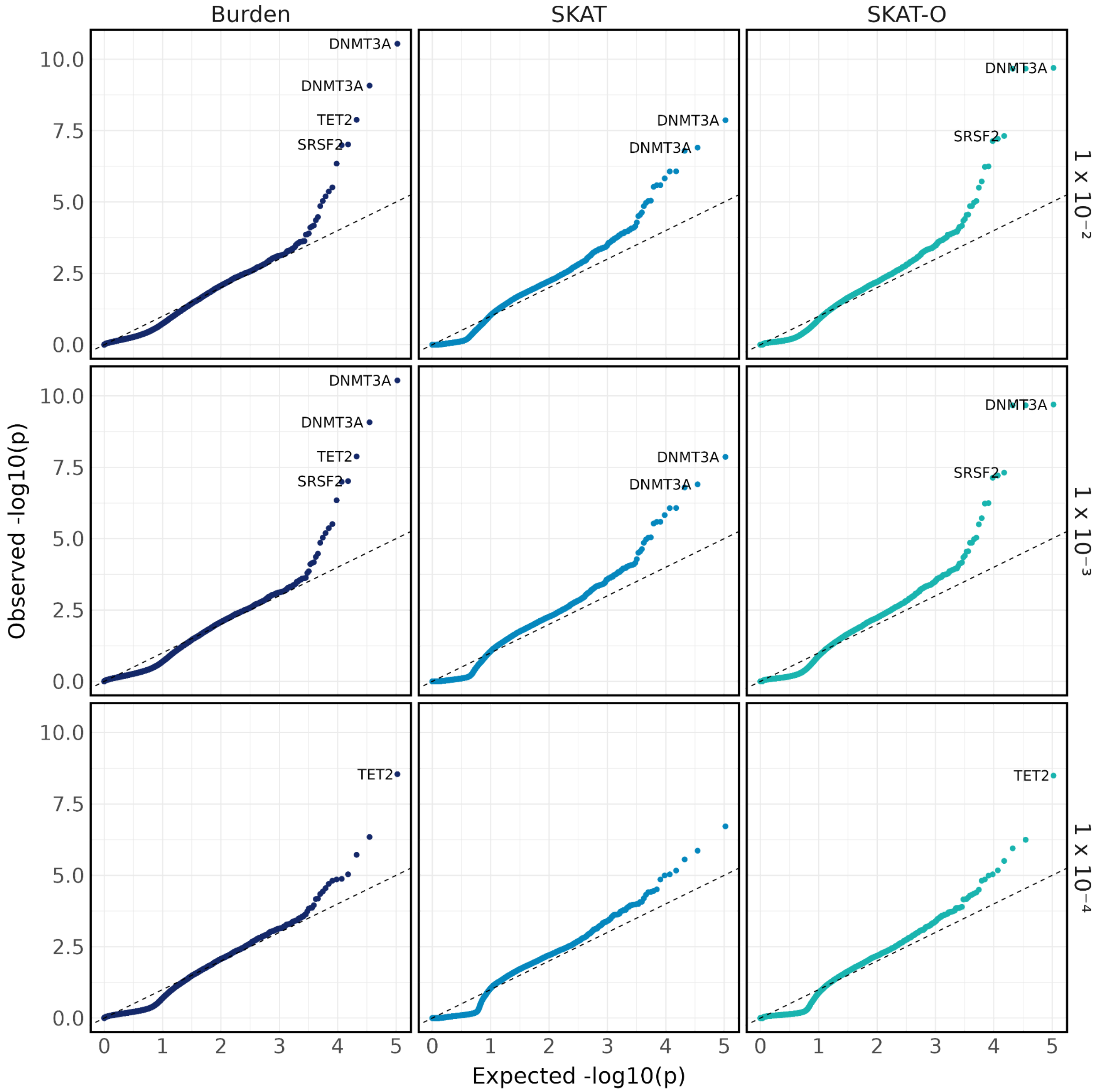


**Supplementary Figure 22. Quantile-quantile plot of AML set-based tests adjusted for the age at collection.** Quantile-quantile plot of p-values from set-based tests adjusting for the age of biosample collection, stratified by test type (Burden, SKAT, and SKAT-O) and maximum MAF thresholds (1%, 0.1%, and 0.01%).

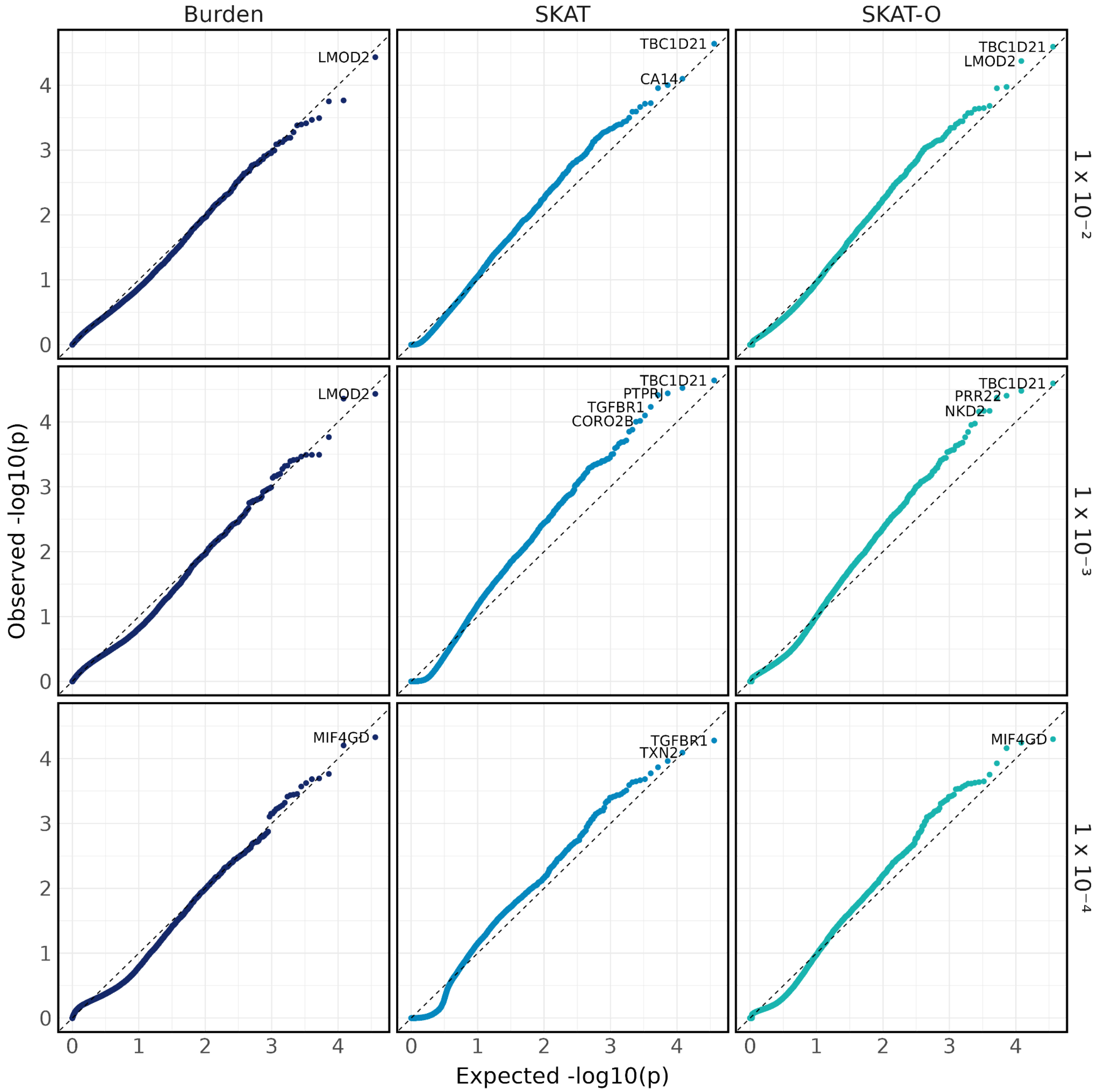


**Supplementary Figure 23. Quantile-quantile plot of AML synonymous set-based tests adjusted for the age at collection.** Quantile-quantile plot of p-values from synonymous set-based tests adjusting for the age of biosample collection, stratified by test type (Burden, SKAT, and SKAT-O) and maximum MAF thresholds (1%, 0.1%, and 0.01%).

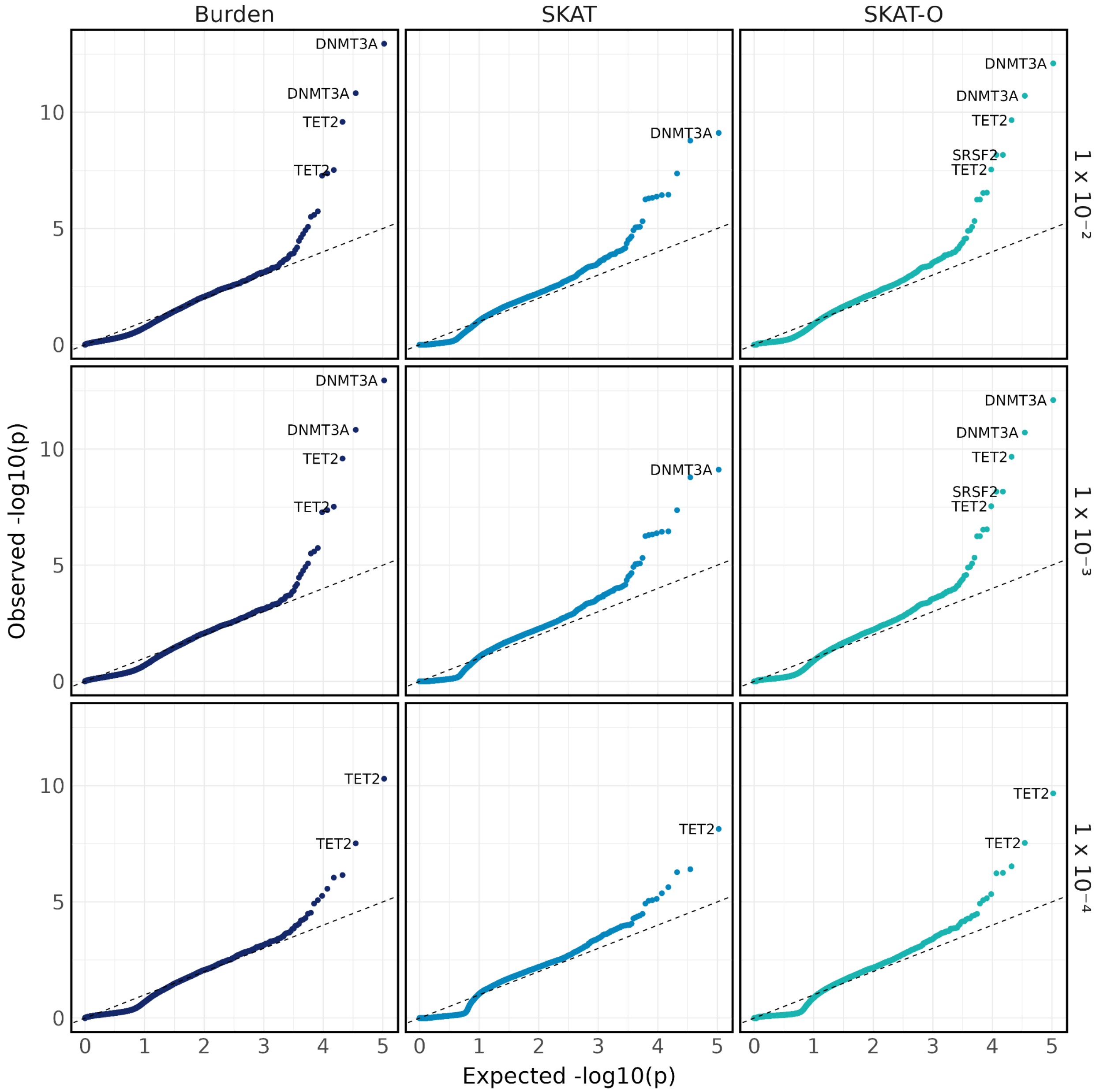


**Supplementary Figure 24. Quantile-quantile plot of AML set-based tests adjusted for the age at diagnosis.** Quantile-quantile plot of p-values from set-based tests adjusting for the age at diagnosis, stratified by test type (Burden, SKAT, and SKAT-O) and maximum MAF thresholds (1%, 0.1%, and 0.01%).

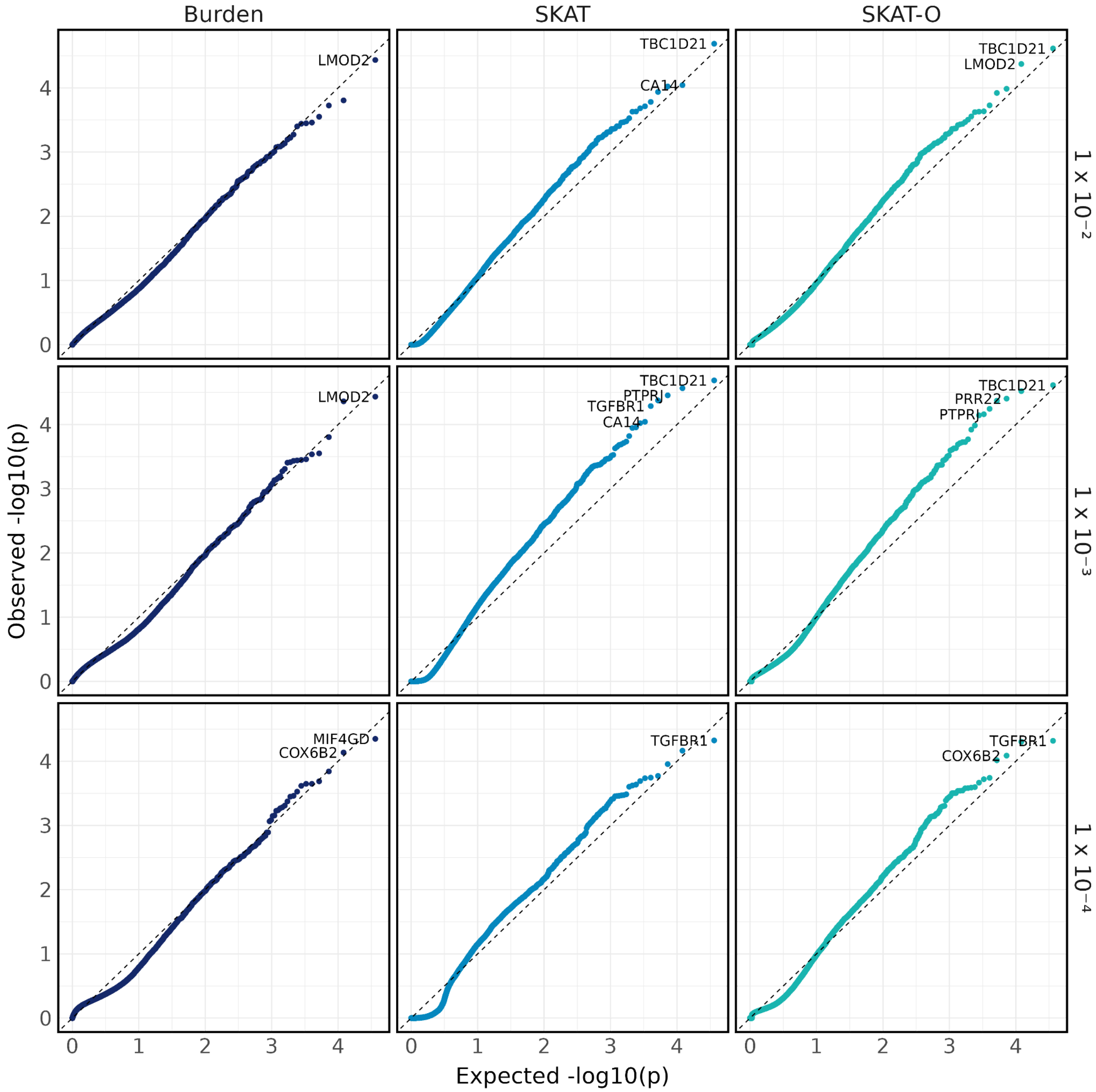


**Supplementary Figure 25. Quantile-quantile plot of AML synonymous set-based tests adjusted for the age at diagnosis.** Quantile-quantile plot of p-values from synonymous set-based tests adjusting for the age at diagnosis, stratified by test type (Burden, SKAT, and SKAT-O) and maximum MAF thresholds (1%, 0.1%, and 0.01%).

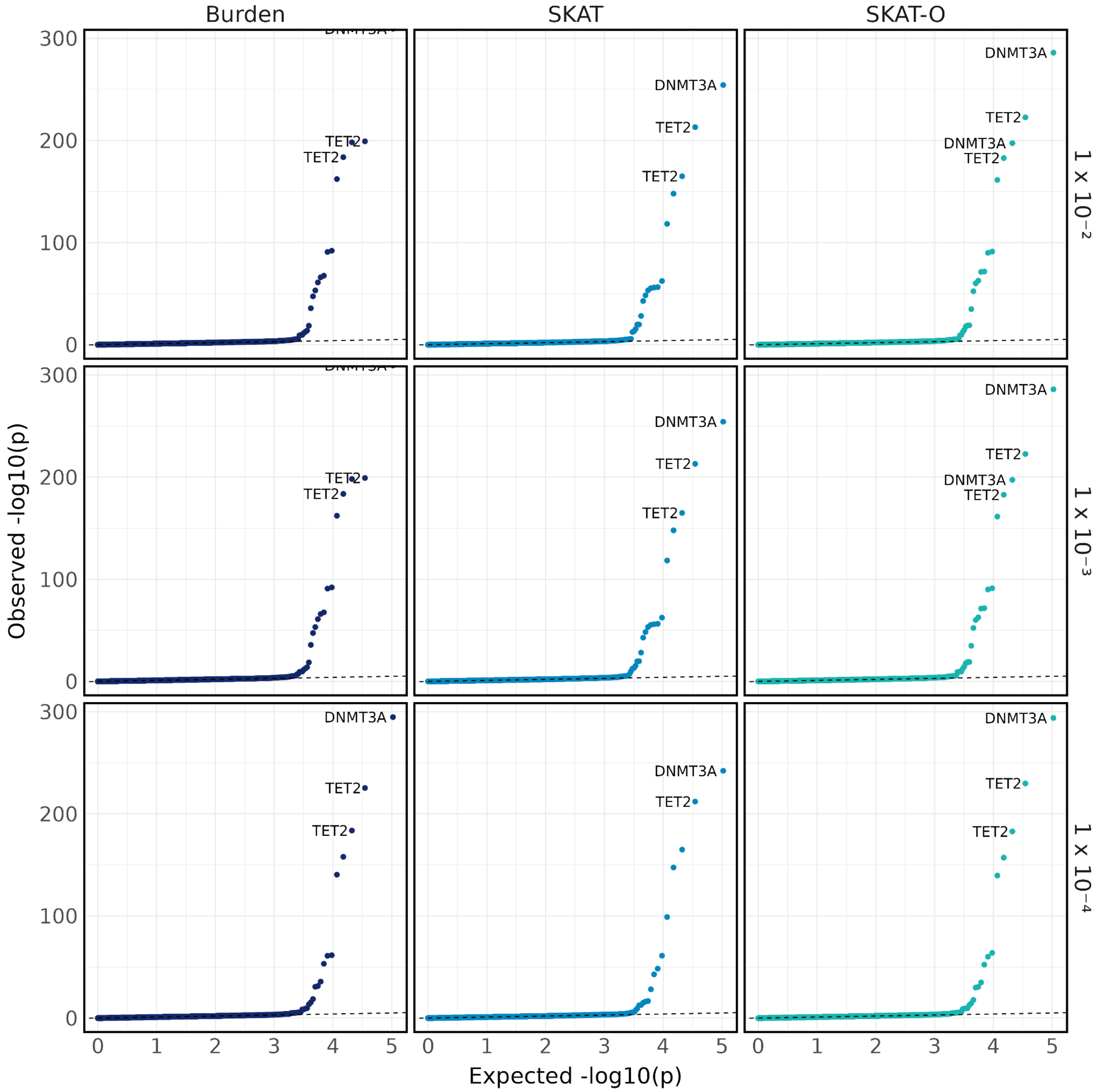


**Supplementary Figure 26. Quantile-quantile plot of age at biospecimen collection set-based tests.** Quantile-quantile plot of p-values from set-based tests, stratified by test type (Burden, SKAT, and SKAT-O) and maximum MAF thresholds (1%, 0.1%, and 0.01%).

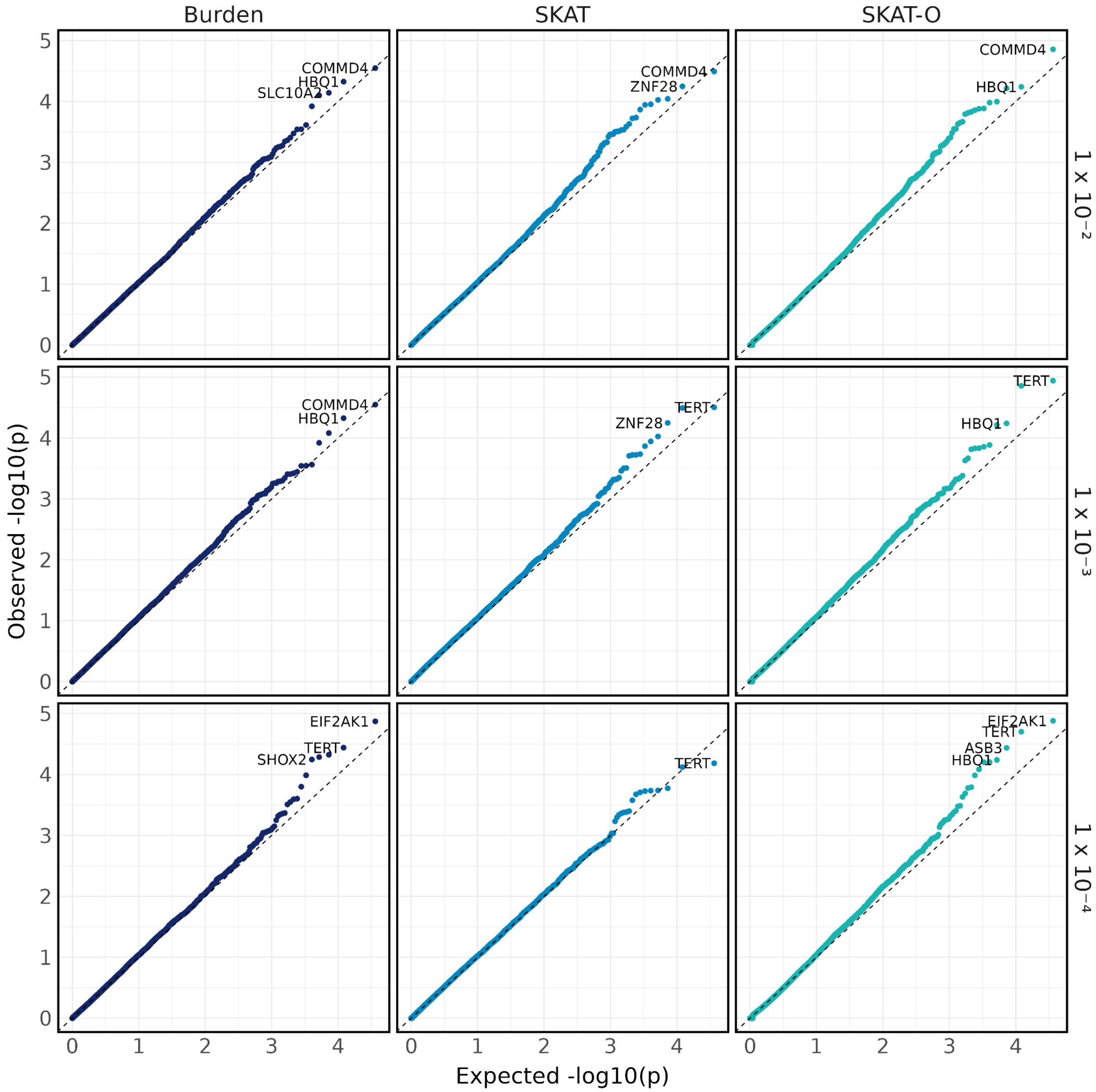


**Supplementary Figure 27. Quantile-quantile plot of age at biospecimen collection synonymous set-based tests.** Quantile-quantile plot of p-values from synonymous set-based tests, stratified by test type (Burden, SKAT, and SKAT-O) and maximum MAF thresholds (1%, 0.1%, and 0.01%).

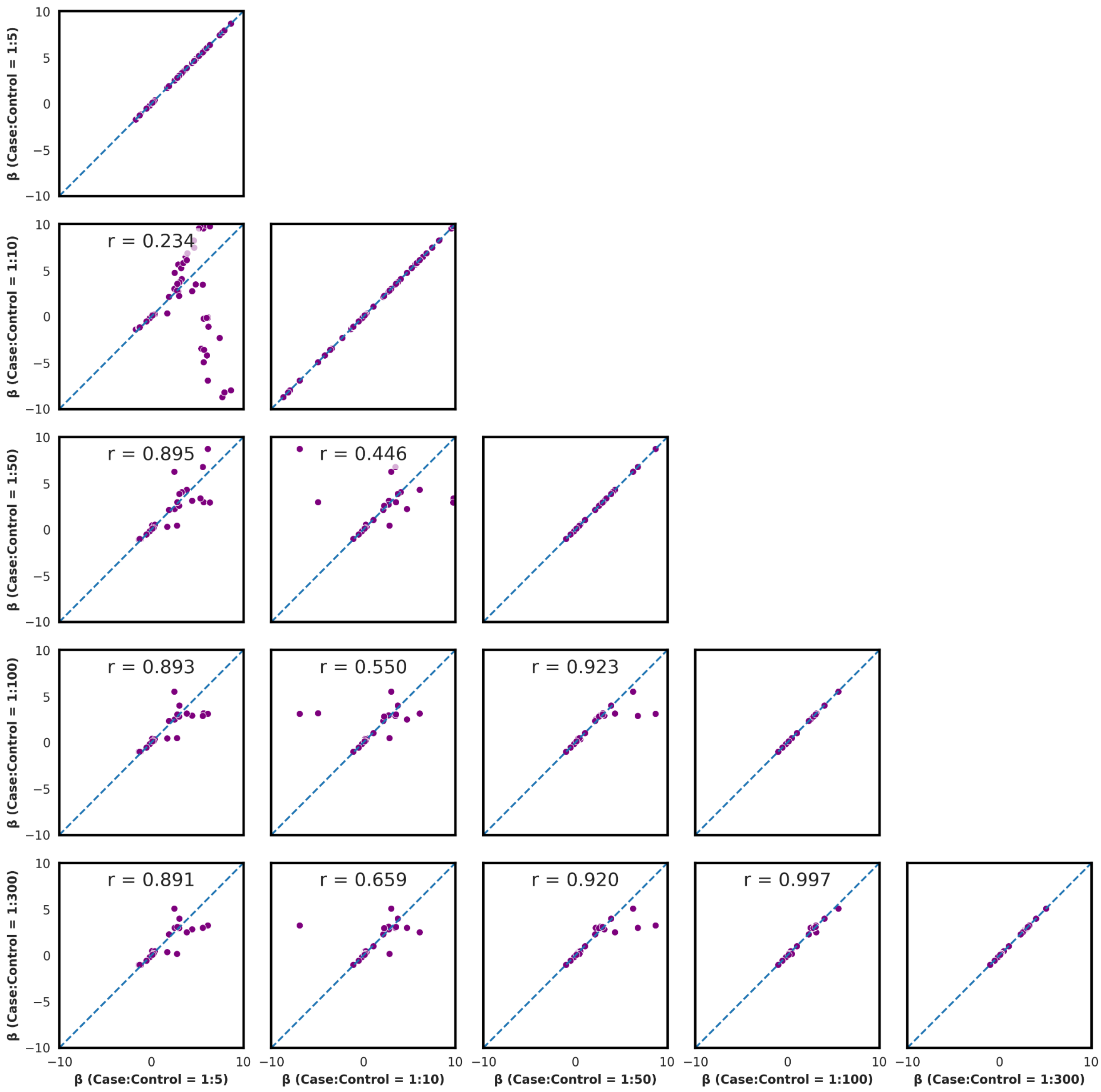


**Supplementary Figure 28. Rare single-variant coefficients in AML top genes in biospecimen-age matched cohorts.** Single-variant tests were performed within genes that were significant in the AML set-based tests (*DNMT3A*, *TET2*, *SRSF2*, and *IDH2*), using AML as the phenotype and conducted within each age-at-biospecimen-collection matched cohort. Correlations of effect sizes for rare variants (0.0001 < MAF < 0.01) were calculated to ensure that the matching procedure did not alter effect size estimates across different matching ratios. The Pearson correlation coefficient is reported above each pairwise comparison.

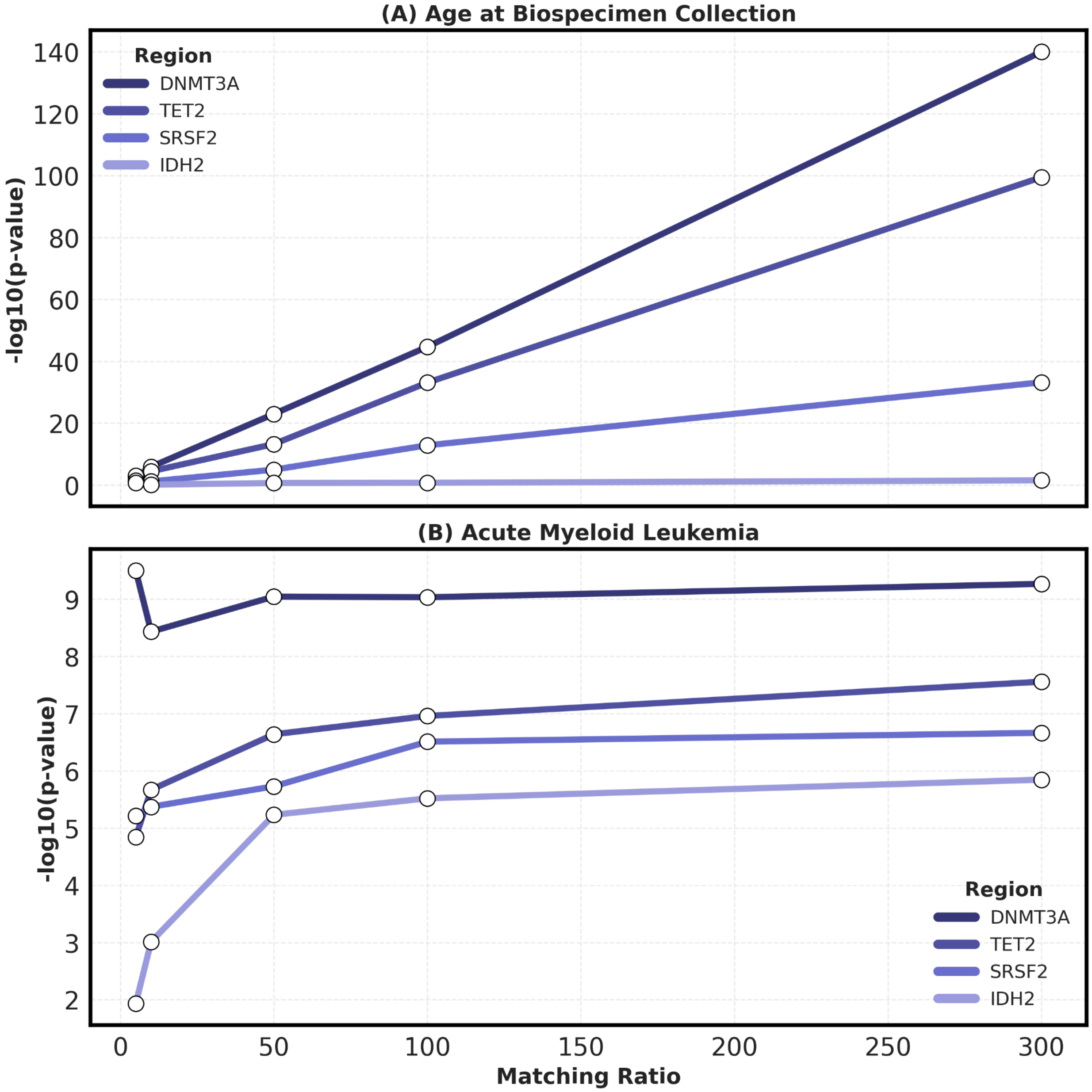


**Supplementary Figure 29. Rare set-based tests in AML top genes in biospecimen-age matched cohorts.** The −log10 p-values from the set-based tests using the age-at-biospecimen-collection matched cohorts are displayed. (A) The −log10 p-values from set-based tests in top AML genes (*DNMT3A*, *TET2*, *SRSF2*, and *IDH2*) using age-at-biospecimen-collection as the phenotype and employing different matched cohort ratios.

(B) The −log10 p-values from set-based tests in top AML genes (*DNMT3A*, *TET2*, *SRSF2*, and *IDH2*) using AML as the phenotype and employing different matched cohort ratios. The y-axis shows −log10 p-values, and the x-axis shows the case-control matching ratios used for the age-at-biospecimen-collection matched cohorts.

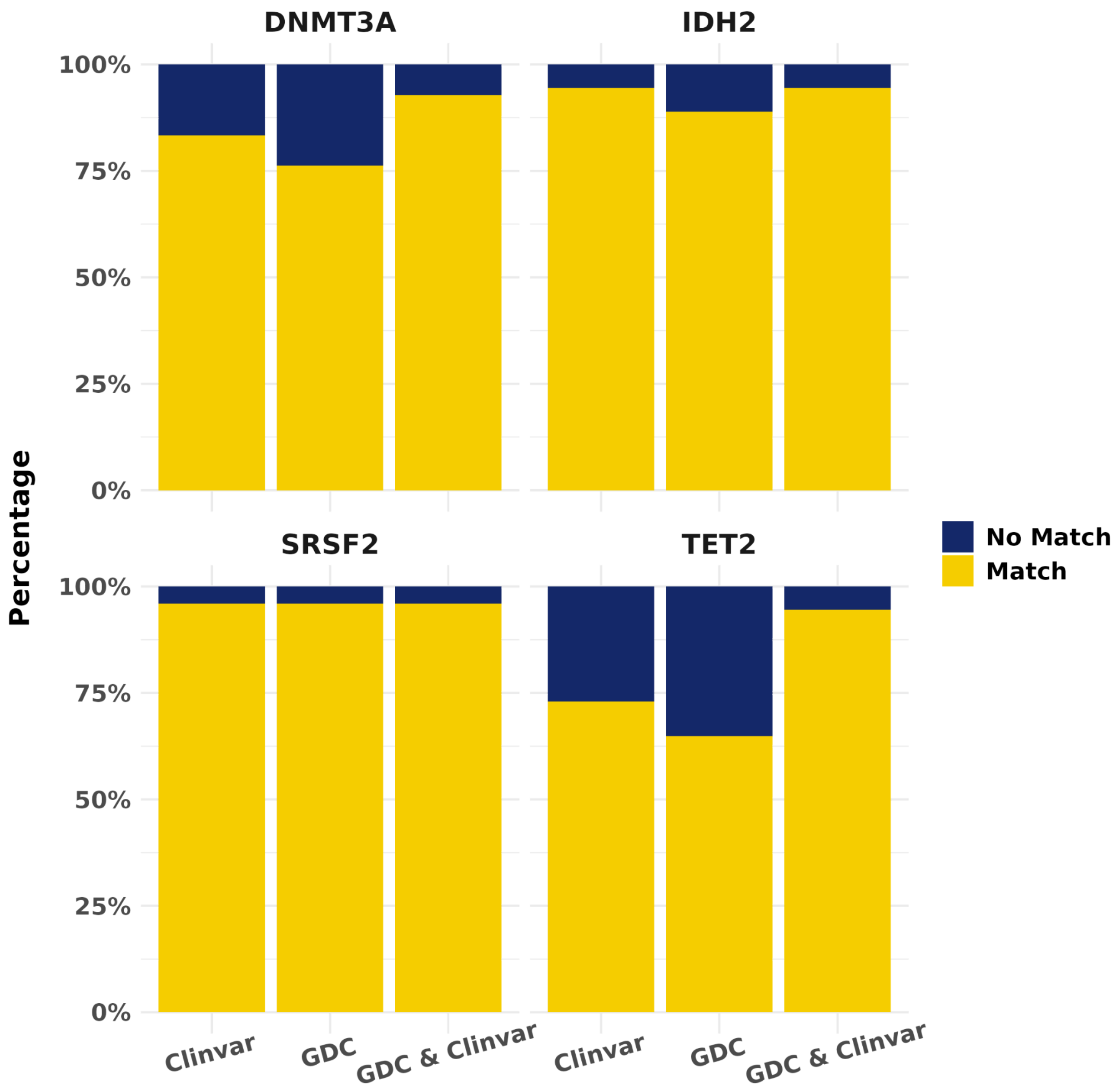


**Supplementary Figure 30. Cross-reference rare variants with the GDC and ClinVar.** For the genes identified from the gene set-based tests, rare variants (MAF < 1% and MAC ≥ 10) from the variant sets were cross-referenced with variants from the GDC and ClinVar databases. The proportion of overlapping rare variants is displayed as a percentage.

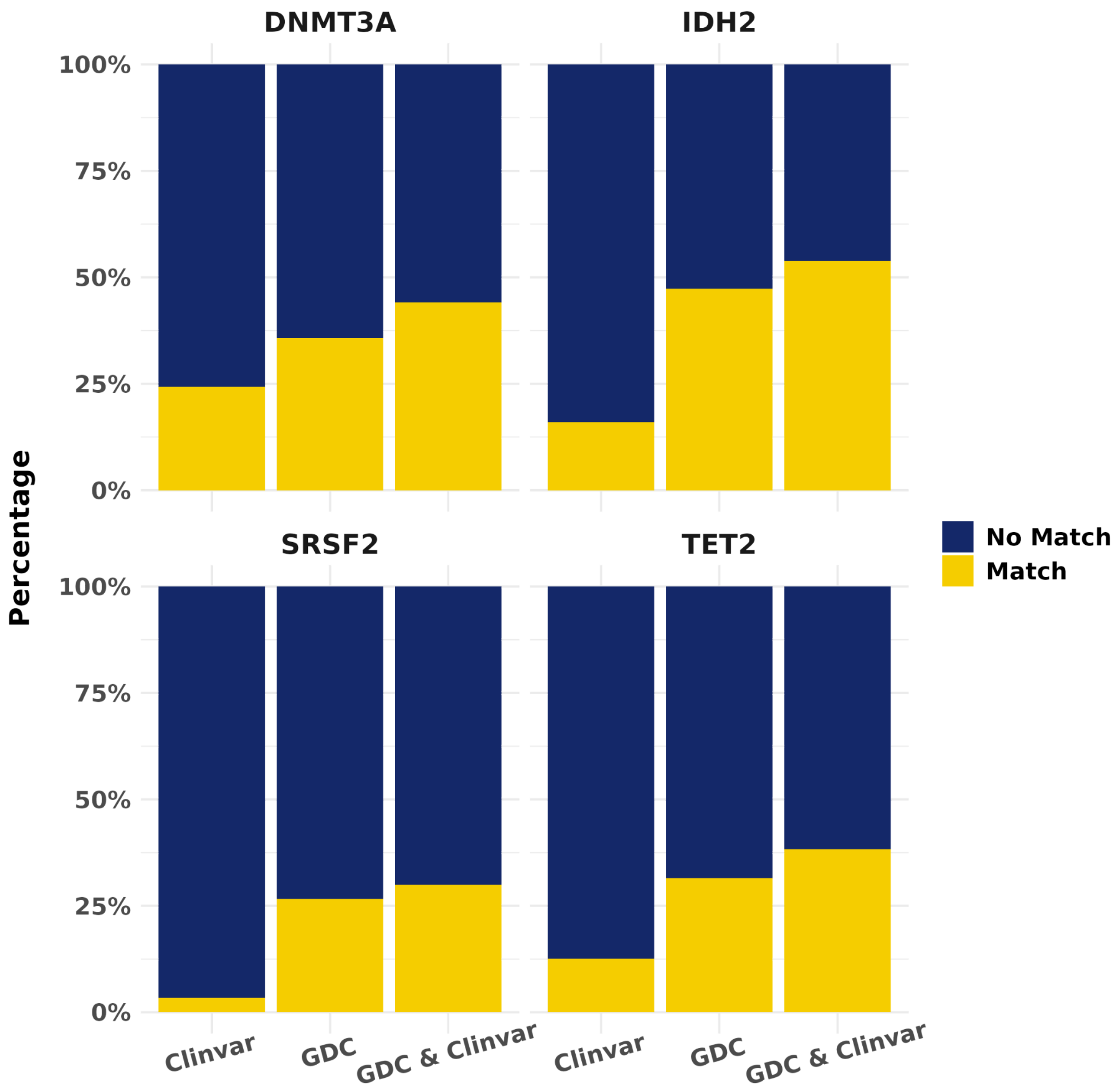


**Supplementary Figure 31. Cross-reference ultra-rare variants with the GDC and ClinVar.** For the genes identified from the gene set-based tests, ultra-rare variants (MAC < 10) from the variant sets were cross-referenced with variants from the GDC and ClinVar databases. The proportion of overlapping ultra-rare variants is displayed as a percentage.

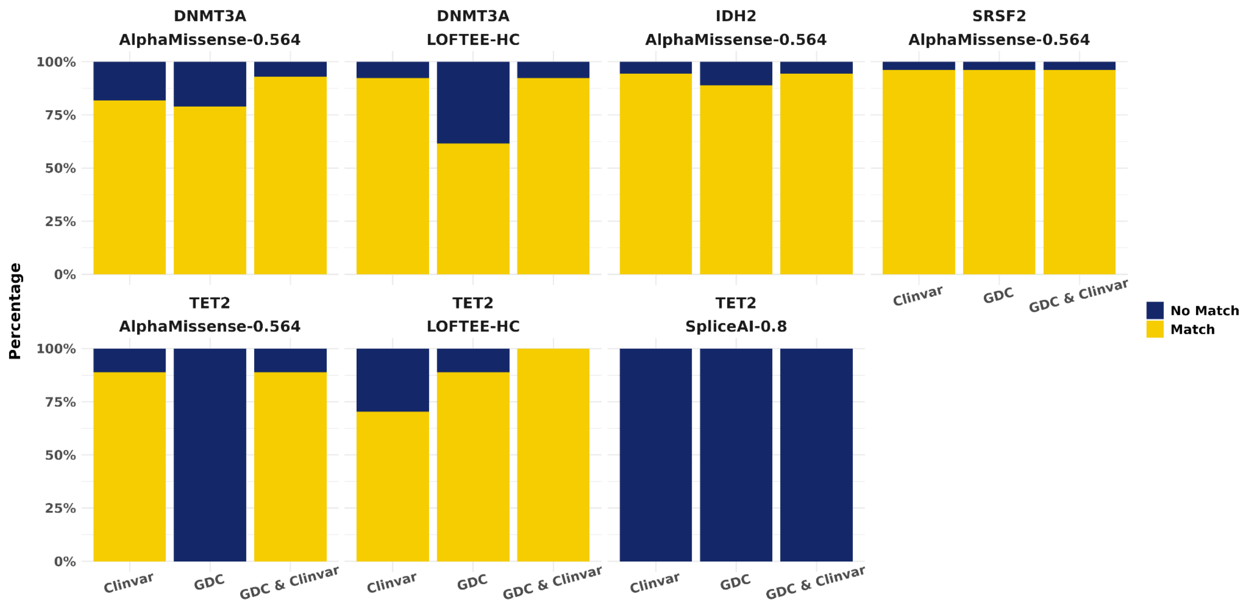


**Supplementary Figure 32. Cross-reference rare variants with the GDC and ClinVar, stratified by annotation tool.** For the genes identified from the gene set-based tests, rare variants (MAF < 1% and MAC ≥ 10) from the variant sets were cross-referenced with variants from the GDC and ClinVar databases. The proportion of overlapping rare variants is displayed as a percentage, stratified by the annotation tools used to designate variant function (LOFTEE, SpliceAI, or AlphaMissense).

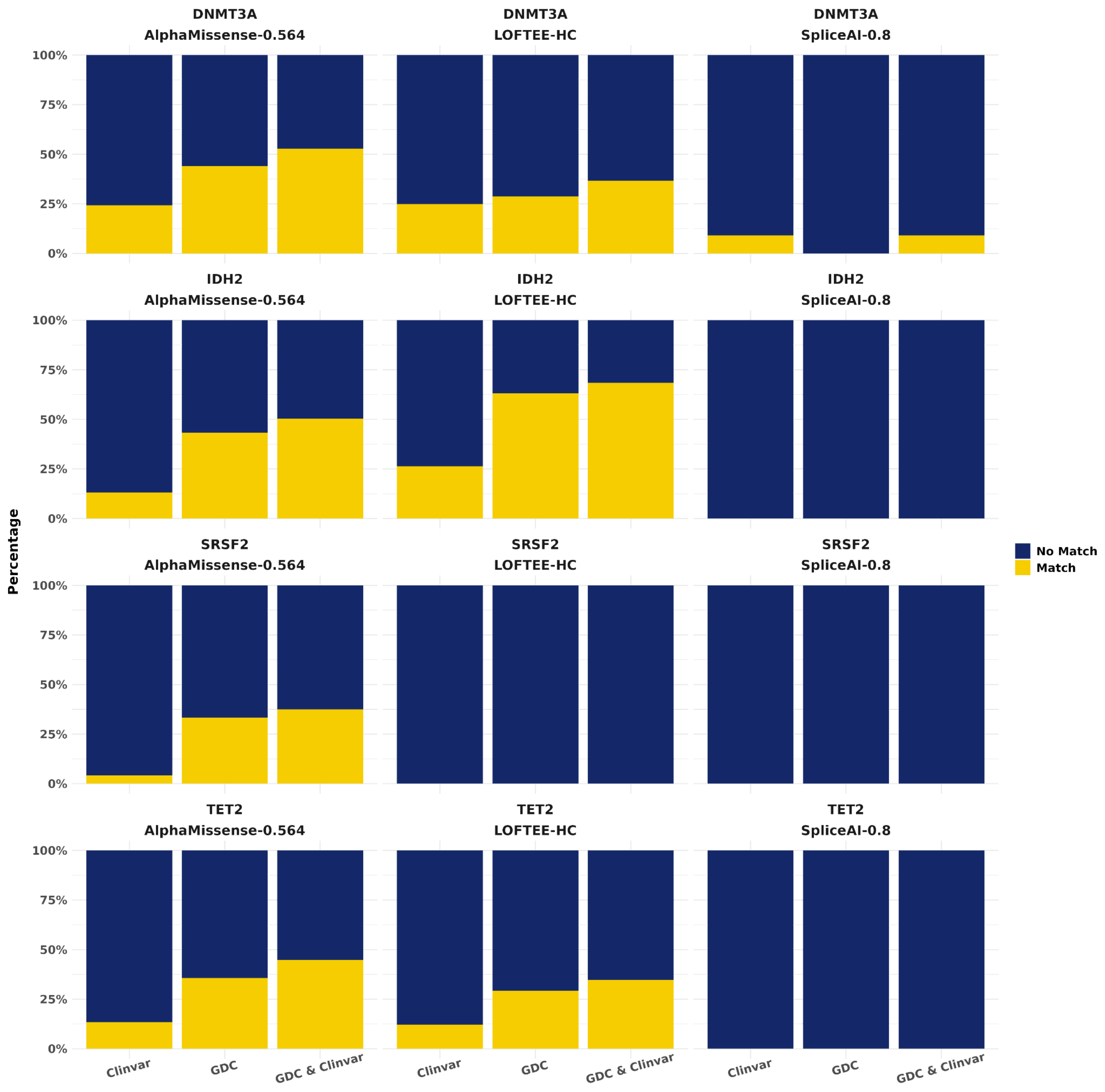


**Supplementary Figure 33. Cross-reference ultra-rare variants with the GDC and ClinVar, stratified by annotation tool.** For the genes identified from the gene set-based tests, ultra-rare variants (MAC < 10) from the variant sets were cross-referenced with variants from the GDC and ClinVar databases. The proportion of overlapping ultra-rare variants is displayed as a percentage, stratified by the annotation tools used to designate variant function (LOFTEE, SpliceAI, or AlphaMissense).

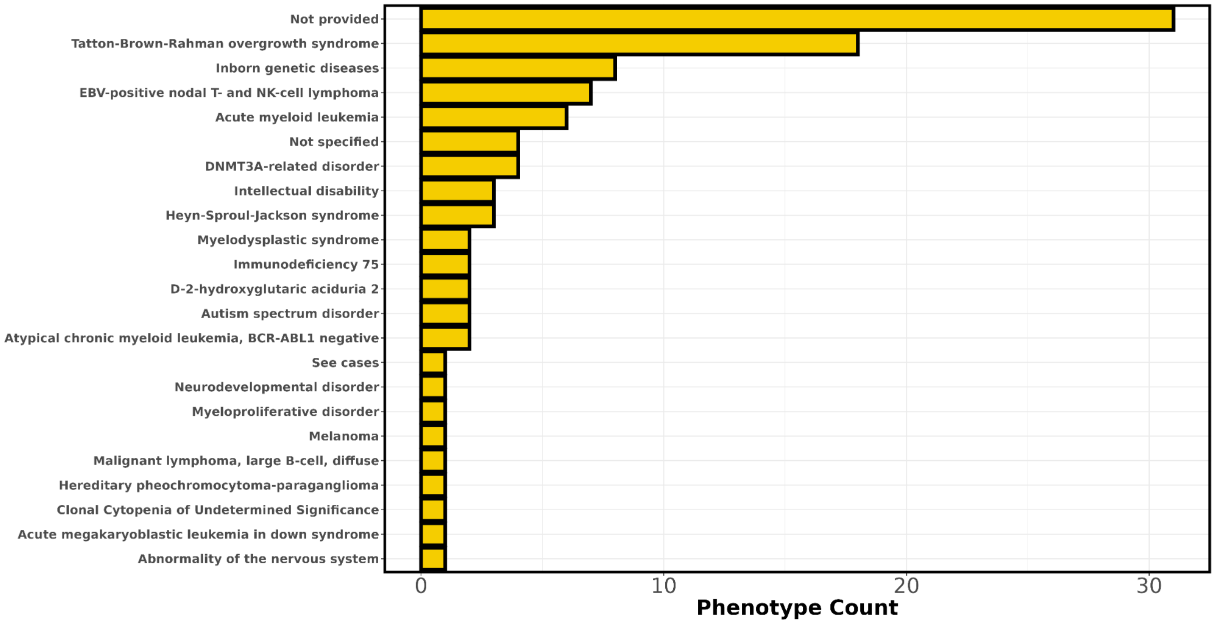


**Supplementary Figure 34. Cross-reference rare variants with associated disorders in ClinVar.** Rare variants (MAF < 1% and MAC ≥ 10) identified in significant genes from the set-based tests were cross-referenced with variants in ClinVar. For variants present in ClinVar, the counts of the most commonly documented phenotype associations are displayed.

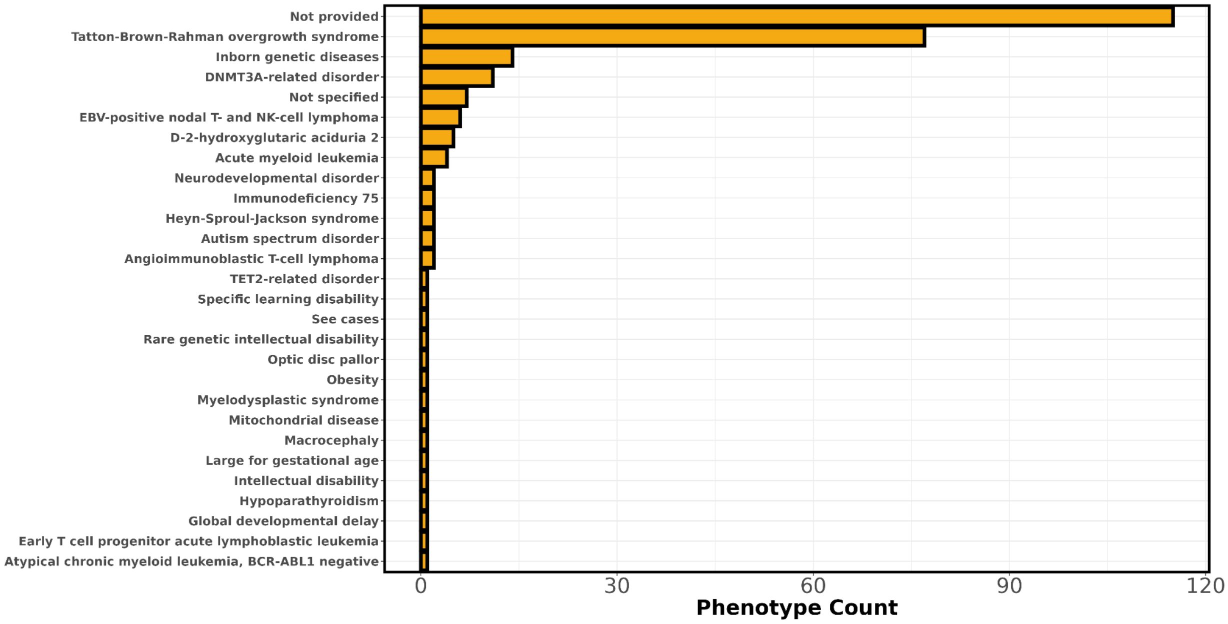


**Supplementary Figure 35. Cross-reference ultra-rare variants with associated disorders in ClinVar.** Ultra-rare variants (MAC < 10) identified in significant genes from the set-based tests were cross-referenced with variants in ClinVar. For variants present in ClinVar, the counts of the most commonly documented phenotype associations are displayed.

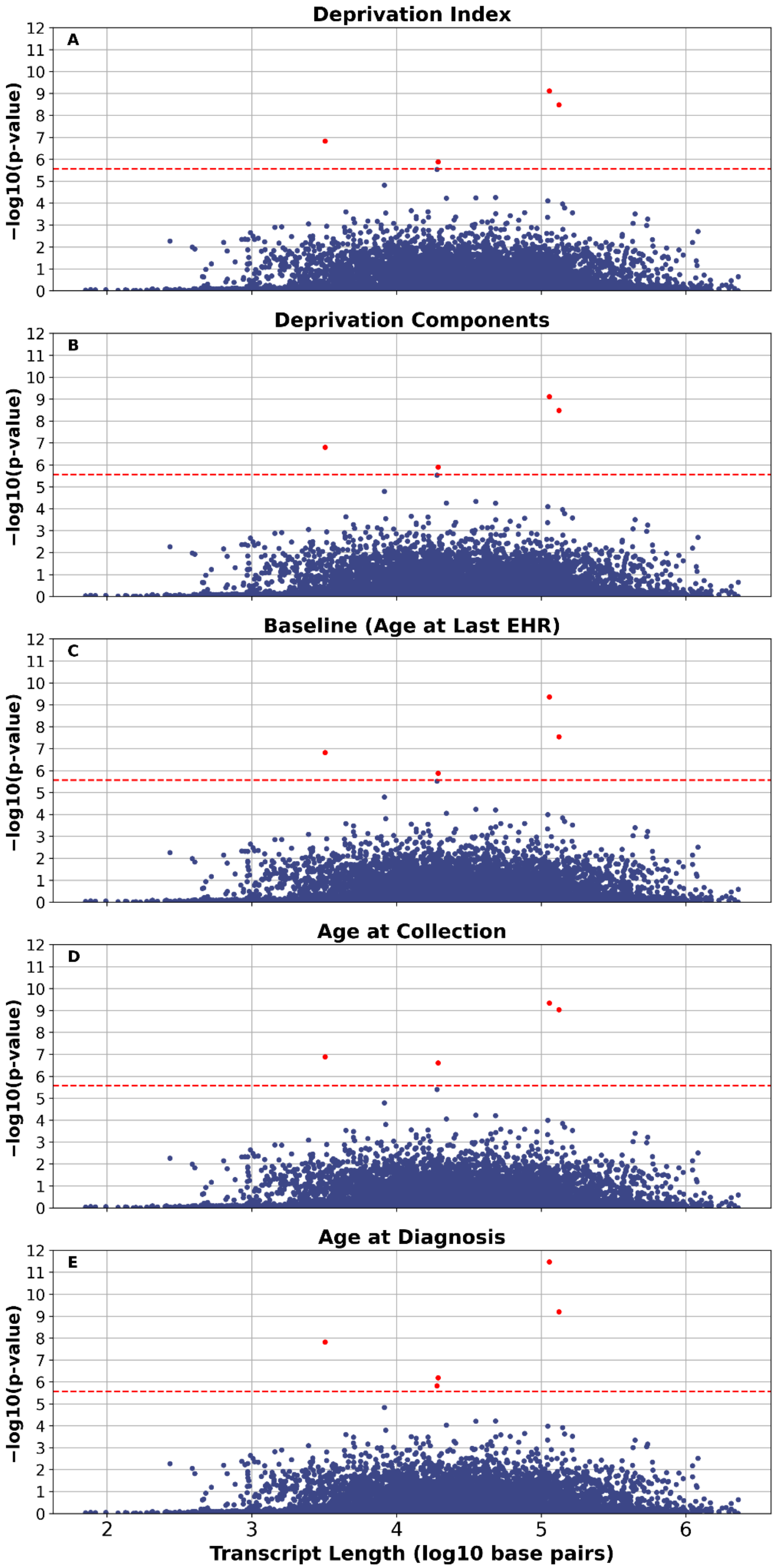

Deprivation Index
A
Deprivation Components
B
Baseline (Age at Last EHR)
C
Age at Collection
D
Age at Diagnosis
E
−log10(p-value)
Transcript Length (log10 base pairs)
0
1
2
3
4
5
6
7
8
9
10
11
12
2
3
4
5
6

**Supplementary Figure 36. Transcript length comparison with Cauchy p-values.** Cauchy SKAT-O p-values from the set-based tests against transcript length, expressed as log10 base-pair length. The "Deprivation Index" model (A) includes the deprivation index as an additional covariate, whereas the "Deprivation Components" model (B) includes the individual components of the deprivation index as covariates. The "Baseline" model (C) uses the standard set of covariates (age, sex, and principal components). The "Age at Collection" model (D) defines age based on age at biosample collection for whole-genome sequencing, while the "Age at Diagnosis" model (E) defines age using age at diagnosis for cases and age at the last EHR record for controls.
