## Supplementary Methods for "Prioritizing Genes and Rare Protein-Coding Variants in Acute Myeloid Leukemia via Whole Genome Sequencing Data"

**Table of Contents**


Overview of electronic health records in AoU

Deprivation index information

Microarray quality control

Whole genome sequencing quality control

Ancestry assignment

Exome variant quality control

ACAF variant quality control

Sensitivity analysis for deprivation index

Sensitivity analysis for age definition

Age at biospecimen collection

Age at biospecimen collection matching

Cross referencing

Global burden gene-sets: Somatic mutation gene-sets

Global burden gene-sets: Cox regression gene-sets

Global burden gene-sets: Differential gene expression gene-sets

Global burden gene-sets: Single-cell differently expressed gene-sets

Limitations

Supplementary Tables

- Gene set-based tests
- Leave-one-variant-out tests
- GAUSS results
- Global protein-coding variant burden tests

Single-variant tests

MAGMA results

Sensitivity analysis for deprivation index

Sensitivity analysis for age definitions

### Supplementary Tables

#### Gene set-based tests

**Supplementary Table 1:** Genomic inflation for gene set-based tests

**Supplementary Table 2:** P-values from set-based tests for AML

**Supplementary Table 3:** Cauchy p-values from set–based tests for AML

**Supplementary Table 4:** P-values from set-based tests for AML adjusted for the deprivation index

**Supplementary Table 5:** Cauchy p-values from set-based tests for AML adjusted for the deprivation index

**Supplementary Table 6:** P-values from synonymous set-based tests for AML

**Supplementary Table 7:** Cauchy p-values from synonymous set-based tests for AML

**Supplementary Table 8:** P-values from synonymous set-based tests for AML adjusted for the deprivation index

**Supplementary Table 9:** Cauchy p-values from synonymous set-based tests for AML adjusted for the deprivation index

#### Leave-one-variant-out tests

**Supplementary Table 10:** Cauchy p-values from leave-one-variant-out tests for AML

**Supplementary Table 11:** Cauchy p-values from leave-one-variant-out tests for AML adjusted for the deprivation index

#### GAUSS results

**Supplementary Table 12:** GAUSS gene-set association results for AML unadjusted for the deprivation index

#### Global protein-coding variant burden tests

**Supplementary Table 13:** Genomic data commons somatic mutation gene-sets

**Supplementary Table 14:** AML RNA-seq Cox regression gene-sets

**Supplementary Table 15:** AML RNA-seq differential gene expression gene-sets

**Supplementary Table 16:** AML single-cell RNA-seq differential gene expression gene-sets

**Supplementary Table 17:** Results of global protein-coding variant burden analysis

**Supplementary Table 18:** Results of global protein-coding variant burden analysis for unrelated participants

#### Single-variant tests

**Supplementary Table 19:** Common single-variant association testing in AML

**Supplementary Table 20:** Common single-variant association testing in AML adjusted for the deprivation index

### MAGMA results

**Supplementary Table 21:** MAGMA gene results from summary statistics unadjusted for the deprivation index

**Supplementary Table 22:** MAGMA gene results from summary statistics adjusted for the deprivation index

**Supplementary Table 23:** MAGMA gene-set results from summary statistics unadjusted for the deprivation index

**Supplementary Table 24:** MAGMA gene-set results from summary statistics adjusted for the deprivation index

### Sensitivity analysis for deprivation index

**Supplementary Table 25:** Logistic regression results for gene variant burden

**Supplementary Table 26:** Cox regression results for gene variant burden

**Supplementary Table 27:** P-values from set-based tests for AML adjusted for the deprivation components

**Supplementary Table 28:** Cauchy p-values from set-based tests for AML adjusted for the deprivation components

**Supplementary Table 29:** P-values from synonymous set-based tests for AML adjusted for the deprivation components

**Supplementary Table 30:** Cauchy p-values from synonymous set-based tests for AML adjusted for the deprivation components

### Sensitivity analysis for age definitions

**Supplementary Table 31:** P-values from set-based tests for AML adjusted for the age at collection

**Supplementary Table 32:** Cauchy p-values from set-based tests for AML adjusted for the age at collection

**Supplementary Table 33:** P-values from set-based tests for AML adjusted for the age at diagnosis

**Supplementary Table 34:** Cauchy p-values from set-based tests for AML adjusted for the age at diagnosis

**Supplementary Table 35:** P-values from synonymous set-based tests for AML adjusted for the age at collection

**Supplementary Table 36:** Cauchy p-values from synonymous set-based tests for AML adjusted for the age at collection

**Supplementary Table 37:** P-values from synonymous set-based tests for AML adjusted for the age at diagnosis

**Supplementary Table 38:** Cauchy p-values from synonymous set-based tests for AML adjusted for the age at diagnosis

#### Age at biospecimen collection tests

**Supplementary Table 39:** P-values from set-based tests for age at biospecimen collection

**Supplementary Table 40:** Cauchy p-values from set-based tests for AML for age at biospecimen collection

**Supplementary Table 41:** P-values from synonymous set-based tests for age at biospecimen collection

**Supplementary Table 42:** Cauchy p-values from synonymous set-based tests for AML for age at biospecimen collection

**Supplementary Table 43:** Rare single-variant association tests for age at biospecimen collection in top AML genes

### Age at biospecimen collection matching

**Supplementary Table 44:** Case-control matching summary statistics

**Supplementary Table 45:** Rare single-variant association tests for AML in biospecimen age matched cohorts

**Supplementary Table 46:** P-values from set-based tests in biospecimen age matched cohorts

**Supplementary Table 47:** Cauchy p-values from set-based tests in biospecimen age matched cohorts

### Cross-reference with GDC and Clinvar

**Supplementary Table 48:** Cross-referencing variants with the GDC and ClinVar

### Cohort Description

**Supplementary Table 49:** Cohort description

45. Finak G, McDavid A, Yajima M, Deng J, Gersuk V, Shalek AK, et al. MAST: a flexible statistical framework for assessing transcriptional changes and characterizing heterogeneity

in single-cell RNA sequencing data. Genome Biol. 2015 Dec;16(1):278. doi:10.1186/s13059-015-0844-5
